# Particle Based Model for Airborne Disease Transmission

**DOI:** 10.1101/2020.04.23.20076273

**Authors:** Michael B Dillon, Charles F Dillon

**Author notes:** **Auspices and Disclaimer**. This work was performed under the auspices of the U.S. Department of Energy by Lawrence Livermore National Laboratory under Contract DE-AC52-07NA27344. This document was prepared as an account of work sponsored by an agency of the United States government. Neither the United States government nor Lawrence Livermore National Security, LLC, nor any of their employees makes any warranty, expressed or implied, or assumes any legal liability or responsibility for the accuracy, completeness, or usefulness of any information, apparatus, product, or process disclosed, or represents that its use would not infringe privately owned rights. Reference herein to any specific commercial product, process, or service by trade name, trademark, manufacturer, or otherwise does not necessarily constitute or imply its endorsement, recommendation, or favoring by the United States government or Lawrence Livermore National Security, LLC. The views and opinions of authors expressed herein do not necessarily state or reflect those of the United States government or Lawrence Livermore National Security, LLC, and shall not be used for advertising or product endorsement purposes.

## Abstract

Prior literature documents cases of airborne infectious disease transmission at distances ranging from ≥ 2 m to inter-continental in scale. Physics- and biology- based models describe the key aspects of these airborne disease transmission events, but important gaps remain. This report extends current approaches by developing a new, single-particle based theory that (a) assesses the likelihood of rare airborne infections (where individuals inhale either one or no infectious particles) and (b) explicitly accounts for the variability in airborne exposures and population susceptibilities within a geographic region of interest. For these hazards, airborne particle fate and transport is independent of particulate concentration, and so results for complex releases can be determined from the results of many single-particle releases.

This work is intended to provide context for both (a) the initial stages of a disease outbreak and (b) larger scale (≥ 2 m) disease spread, including distant disease “sparks” (low probability, unexpected disease transmission events that infect remote, susceptible populations). The physics of airborne particulate dispersion inherently constrains airborne disease transmission. As such, this work suggests results that, *a priori*, may be applicable to many airborne diseases.

**Model Predictions:**

i. Modeling predictions of the single-particle transmission kernel suggest that outdoor airborne disease transmission events may occur episodically as the infection probabilities can vary over many orders of magnitude depending on the distance downwind; specific virus, prion, or microorganism; and meteorological conditions.
ii. Model results suggest that, under the right conditions, an indoor infected person could spread disease to a similar, or greater, number of people downwind than in the building they occupy. However, the downwind, per-person infection probability is predicted to be lower than the within-building, per-person infection probability. This finding is limited to airborne transmission considerations.
iii. This work suggests a new relative disease probability metric for airborne transmitted diseases. This metric, which is distinct from the traditional relative risk metric, is applicable when the rate at which the infectious agent losses infectivity in the atmosphere is ≲ 1 h^-1^.

## 1. Introduction

Particles of all types are routinely transported in the atmosphere, including infectious disease particles. The physics of the local airborne person-to-person inhalation pathway of infectious disease transmission was investigated by early Public Health investigators who were focused on human Tuberculosis and Measles outbreaks [1]–[3]. As documented in the **Supplemental Material A: Airborne Disease Transmission Literature Review**, airborne human, veterinary, and plant disease transmission and other bioaerosol transmission events naturally occur across a wide range of distance scales, from two meters up to and including inter-continental transmission. These multiscale transmission events include bacteria, viruses, and fungi.

The population disease burden caused by infectious agents that are transmitted through the air remains substantial as lower respiratory infections and Tuberculosis are the fourth and tenth, respectively, leading causes of death world-wide [4], [5]. We adopt here the classic definition of the airborne disease transmission pathway as the inhalation of ≤5 μm aerodynamic diameter (AD) infectious particles that have traveled airborne a distance of two or more meters.^1^ We note that the airborne pathway can be the primary means of disease transmission or a secondary pathway when more than one disease transmission pathway is active, including the droplet (> 5 μm AD particles) or contact pathways, e.g., [6], [7].

Physics- and biology- based atmospheric transport and dispersion models have been developed to predict airborne infectious disease transmissions, exposures, and the subsequent infection or disease incidence to various degrees of accuracy, e.g., [8]. However, important scientific gaps remain (discussed below). The aim of physics- and biology- based models is to determine (a) the extent of airborne disease spread (including identification of disease sources) and (b) disease outbreak control strategies. These remain key areas of research and this report extends current, physics- and biology- based modeling approaches. Specifically, we develop a new theory that provides methods and metrics to assess rare, but finite airborne infection incidence (single particle airborne disease transmission: dilute atmospheric exposures where an individual inhales either one infectious particle^2^ or none at all).

In this report, we highlight implications for two distinct categories of diseases. In the first case, the disease does not have significant individual to individual transmission (R_0_ < 1).^3^ One example is Q Fever in humans in which the source of infectious particles is outdoors (i.e., infected livestock and/or contaminated soil) and infections are not typically passed from one human to another. In the second case, the disease has notable individual to individual transmission (R_0_ > 1) and so there may be the potential for rapid disease spread. We note that both disease cases may have multiple disease transmission pathways, including direct contact, indirect contact, or ingestion.

## 2. Physics- and Biology- Based Airborne Transmission Modeling

Airborne transmission of infectious disease from an infected host to a susceptible individual occurs in three phases: (a) the release of infectious particles into the air; (b) particle transport to, and the exposure of, downwind individuals; and (c) the infection of susceptible exposed individuals (receptors), see **Figure 1**. Physics and biology based models mathematically describe each of these three phases. They estimate the number and/or distribution of subsequent infections using previously determined principles combined with key particle, environmental, infectious organism, and receptor properties, see **Table 1**. This paper focuses on improvements for modeling the second and third phases.

**Table 1.** **Key properties affecting the airborne disease transmission pathway**

| **Particle**<br>**Physical**<br>**Properties** | Particle number |
| --- | --- |
|  | Particle size (aerodynamic diameter) and infectivity* as a function of (a) time, (b) the infectious agent and (c) environmental conditions (e.g., temperature, humidity, solar radiation) |
| **Environmental**<br>**Properties** | Wind speed, Turbulence, Precipitation, Temperature, |
|  | Humidity, Solar radiation, Terrain/Land Use, and<br>Built Environment (for indoor exposures) |
| **Infectious**<br>**Agent**<br>**Properties** | Infectivity* as a function of (a) time and<br>(b) particle, environmental, and receptor properties |
|  | Pathogenicity (ability to cause disease) |
| **Receptor (Host)**<br>**Properties** | Breathing rate as a function of the receptor and environmental properties<br>(e.g., activity, temperature) |
|  | Number of receptors (population) |
|  | Factors affecting infection probability*<br>Age, Gender, Nutrition, Health Status, Immune status |

\* The probability that an inhaled particle causes infection for a given receptor is affected by parameters in several columns. Specifically, it depends on a combination of particle, infectious agent, and receptor properties - each of which depends, in part, upon environmental properties.

**Figure 1.**
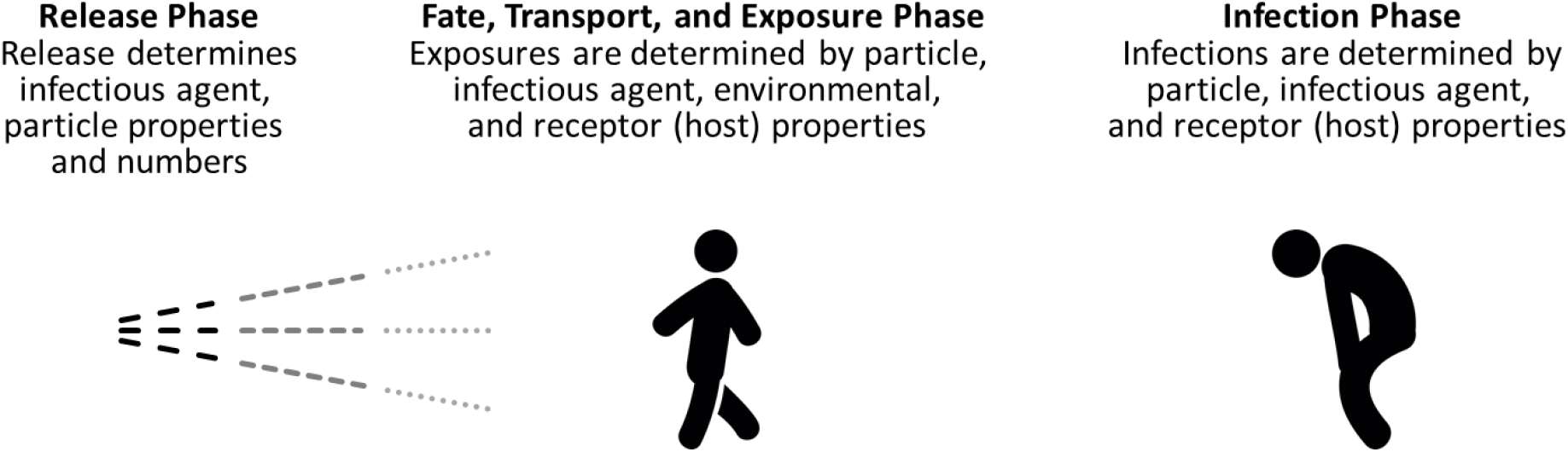
Conceptual model of the airborne transmission pathway

### 2.1. Dilute Particle Considerations

If airborne particles are added to a small mass of air, the resulting particle concentration is, in many cases, dilute and the material’s presence will not significantly affect the local atmosphere.^4^ Therefore the downwind transport and dilution (dispersion) of such material can be determined from environmental (meteorological + surface) considerations alone. Also, as a practical matter, dilute particles can be assumed to behave *independently* in the atmosphere, i.e., there is essentially no particle-to-particle interaction. Therefore, the downwind exposure associated with any release of dilute material is mathematically identical to the sum of the exposures associated with many independent “point” releases that occur at specific points in time and space. Thus, insights and model results derived for single particles released at a specific location and time can be combined to characterize the more complex cases, such as complex particle aerodynamic diameter distributions and/or time varying locations and release quantities.

### 2.2. Advantages of Spatial and Temporal Averaging

Physics-based models of atmospheric transport and dispersion simulate the effects of the physical processes which determine the air concentrations of infectious particles downwind of a release. All practical dispersion models resolve only a portion of the atmospheric motions and physical processes, typically the regional winds that transport particles downwind. The effects of the unresolved processes, such as turbulent eddies that dilute the airborne particulate cloud, are parameterized, often using empirical data. An important consequence is that model predictions can be much more accurate with predicting metrics that are spatially and/or temporally integrated (or averaged), see **Supplemental Material B: Key Atmospheric Transport and Dispersion Modeling Concepts**. This study uses the exposure integrated over a (a) a disc (circular area) and (b) along a circle arc both centered at the release point, see **Figure 2**. These exposure metrics also eliminate sensitivity to wind direction uncertainty, although we note that other uncertainties, such as wind speed, remain.^5^

**Figure 2.**
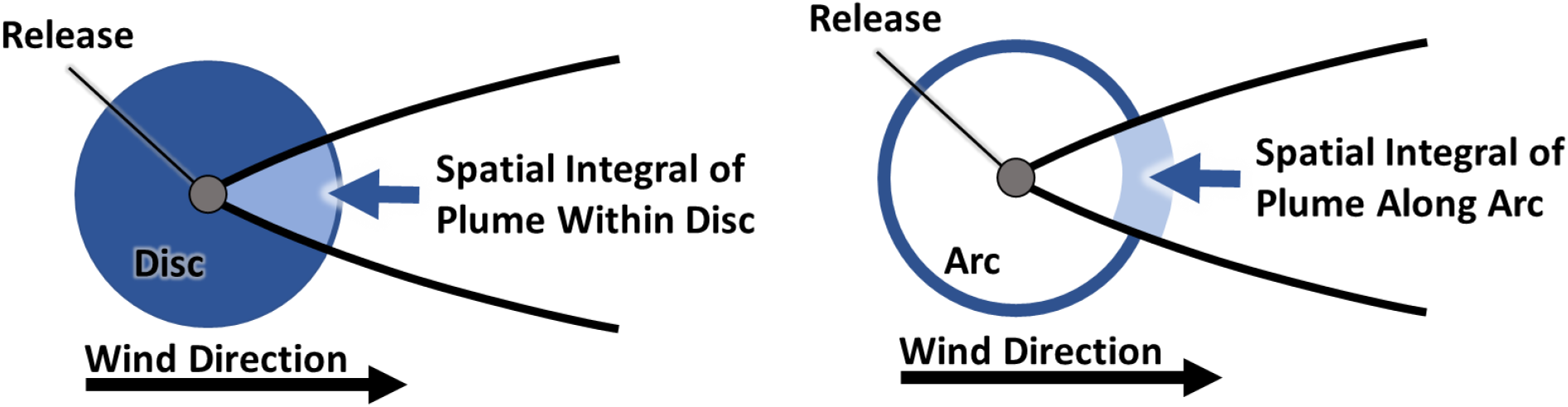
Illustration of airborne exposure (plume) integrated over a (left) a disc and (right) along a circle arc, both centered at the release point.

### 2.3. Discrete Nature of Airborne Infection

The research community understands that airborne, infectious agents are carried as or within discrete particles, either as individual viruses, prions, or microorganisms or as part of a larger particle which could contain more than one infectious agent. However, there has been limited attention, within the context of risk assessment, as to how the discrete, quantal nature of airborne particulate exposures affect downwind infection probabilities. This scientific gap is particularly notable for rare exposures - defined for the purposes of this paper as exposures where a typical individual inhales either one infectious particle or none at all, see **Supplemental Material C: General Theory**.

Historically, rare exposure infection probabilities have been estimated by (a) extrapolating population-mean dose-response models to “less-than-one” infectious agent exposures^6^ or (b) using “single hit” models. Population-mean dose-response models are often based on data from toxicology experiments that use multiple infectious agent exposures to extrapolate to single particle exposures. Single hit models assume that (a) the number of infectious agents inhaled by a given individual is a random number whose expected value equals the predicted exposure and (b) the likelihood of an individual inhaling an infectious organism is independent of the total number of infectious agents inhaled. Effectively each infectious agent inhaled is assumed to cause infection independently, although the overall infection probability can depend on other factors, including exposure to other viruses or microorganisms [3], [15]–[19]. Recent work has extended single hit models to consider infectious agents randomly aggregating while in the environment, see [20], [21] and refs therein. However with the exception of a single theoretical treatment of a complex exposure distribution [19], we did not identify a prior approach that explicitly addresses a real-world situation in which (a) a distribution of infectious particles types is aerosolized, (b) the number of infectious agents present in each particle may scale with particle volume (i.e., is not random), and (c) environmental fate (and hence downwind exposures) vary by particle type (again is not random).

Separately, we note that the current physics-based airborne exposure models have been validated against measurements of high concentrations of airborne particulates (and gases). As such they do not explicitly consider the quantal nature of exposures inherent to rare exposures to particulate matter, i.e., the downwind individuals are exposed to either one or no particles. The theory developed later in the paper addresses this historical gap, see the ***3. Theory*** section.

### 2.4. Building Protection

Ideally, airborne infectious particle risks would be assessed at the location where the exposure takes place. It is generally acknowledged that buildings afford protection from outdoor airborne infection. However most current assessments of downwind exposures to airborne infectious particles only consider outdoor exposures and typically do not consider the degree to which buildings protect their occupants from such hazards. For example, in a recent review of existing pathogenic, bio-aerosol dispersion modeling literature; only two studies were included that consider the degree to which indoor exposures may differ from outdoor exposures and no general theory was discussed [8]. This gap is notable as (a) individuals spend about 85% of a typical day indoors [22]; (b) outdoor airborne biological particles have been proven to infiltrate indoors and be inhaled by building occupants, e.g., [23], [24]; and (c) the scientific and public health communities have long recognized the importance of the contribution of indoor exposures during disease outbreaks involving outdoor, airborne plumes of infectious particles. For example, outdoor-origin fungal pathogens such as *Aspergillus*, including *fumigatus* and *flavus*, and *Histoplasma capsulatum* are known pose a particularly severe hazard to indoor, immune compromised individuals and hospital facilities are engineered to minimize such concerns [25]–[29].

The degree to which building occupants are protected from outdoor, airborne, infectious particles is determined by (a) the building air change rate (the rate at which outdoor air passes through air handling systems, open windows and doors, and/or cracks in the building envelope) and (b) indoor particle dynamics (e.g., deposition, resuspension and rate at which infectious agents lose infectivity), see **Supplemental Material D: Indoor Particle Dynamics**. We, and collaborators, have recently developed a proof-of-concept method, Regional Shelter Analysis (RSA), to (a) estimate the degree of protection US buildings provide against outdoor airborne particulate hazards and (b) estimate total population exposures [30], [31]. **Figure 3** illustrates the RSA method for calculating building protection-adjusted population impacts resulting from an estimate of the outdoor hazard. The example shown is for an external gamma radiation hazard. The process for inhalation exposures is similar. US building protection against particulate hazards varies strongly, i.e., by orders of magnitude, with particle size and building use (occupancy). For context, being inside a typical US residence is expected to reduce outdoor exposures by a factor of 5 for 1 μm AD particles, a size typical of many individual bacteria [30].

**Figure 3.**
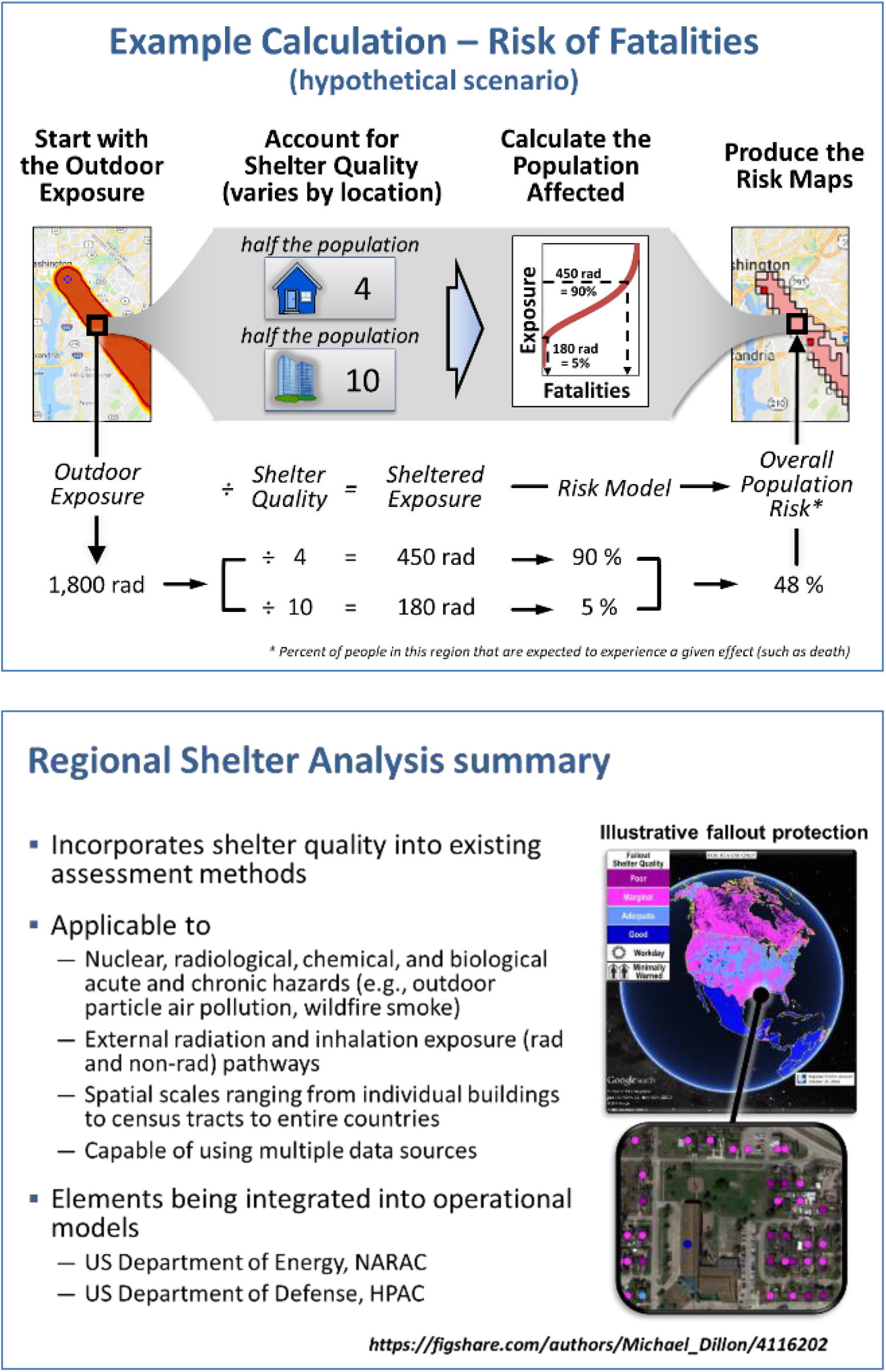
Regional Shelter Analysis illustrative calculation (top panel) and summary (bottom pane). This schematic illustration uses an outdoor (external) radiation exposure and shelter quality (building protection) estimates. Potential applications and uses are listed.

## 3. Theory

Theoretically, the total number of infections in a disease outbreak could be calculated by considering every exposed person. Computationally this approach is challenging to implement, due in part to the need to acquire high resolution input data such as individual exposures and responses to each exposure. To provide an alternative approach, we develop here infection probability prediction equations for (a) the number of airborne infections (and the absolute airborne infection probability) expected in a specific geographic region for both the general and the rare exposure cases (**Equations 1** and **2**) and (b) an equation that relates the relative probability of airborne infection across two regions (**Equation 3**). Note that these, and subsequent, equations can also be used directly without modification to estimate disease probability and other probabilities of interest. These equations are based on the atmospheric physics assumptions and considerations presented in the *2. Physics-and Biology- Based Airborne Transmission Modeling* section. **Supplemental Material C: General Theory** formally derives these equations and provides more detail, including theory that considers multiple particle types that have varying infectivity and environmental fates. **Supplemental Material D: Indoor Particle Dynamics** provides more detail on **Equations 5** to **7** which predict the degree to which buildings protect their occupants from outdoor airborne hazards; expected numbers of infections in the indoor environment from an indoor release; and the fraction of indoor, airborne, infectious particles that may exit a given building into the outdoor atmosphere.

### 3.1. Absolute (Mean) Infection Probabilities for Geographic Regions

Geographic regions, e.g., zip codes and census tracts, are often used when reporting epidemiological data and defining outbreak response zones, e.g., for quarantine and/or vaccination. In general, people within these geographic regions may have varying exposures and/or responses to a given exposure, e.g., individuals inside a building may receive a smaller exposure than those outside; immune compromised individuals may be more sensitive than a healthy person. To account for these considerations, we extend the previously developed Regional Shelter Analysis methodology [30], [31]. Specifically, we assign each group of people within a geographic region a linear scaling factor, termed *adjustment factor*, to account for both exposure and response variability.

For a given release, the total number of airborne infections in a region can be determined by **Equation 1. Equation 2** can be used when the (a) population density and adjustment factor are constant within a region, e.g., the geographic entity is a residential area, and (b) exposures are rare.

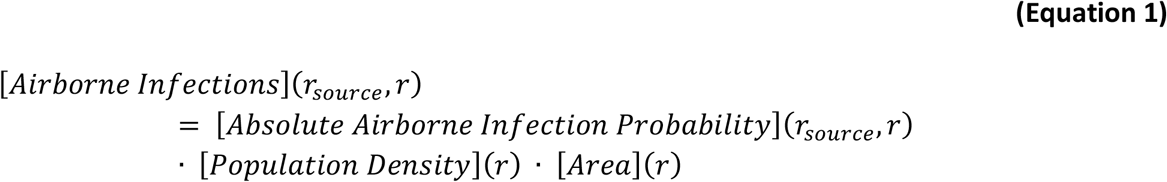

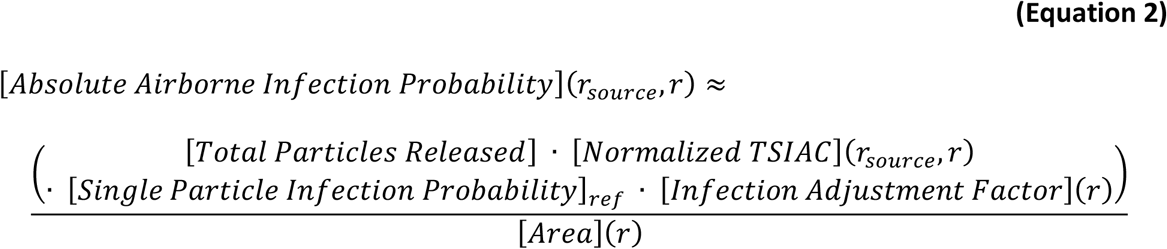

where

*r* = a specific geographic region (dimensionless),

[*Absolute Airborne Infection Probability*](*r*_*source*_, *r*) = mean probability that a random individual in region *r* becomes infected by inhaling an airborne particle emitted from region *r*_*source*_. (dimensionless),

[*Airborne Infections*](*r*_*source*_, *r*) = total number of people who become infected in region *r* by inhaling infectious airborne particles emitted from region *r*_*source*_. (people),

[*Area*](*r*) = area of region *r* (m^2^),

[*Infection Adjustment Factor*](*r*) = linear scaling factor for region *r* that accounts for the deviation of exposure and infection response from the reference exposure and infection response (dimensionless),

[*Normalized TSIAC*](*r*_*source*_, *r*) = particle air concentration integrated over region *r* and the passage of the airborne infectious plume assuming that a single particle has been released to the air from source region *r*_*source*_. TSIAC = time and space integrated air concentration. (s m^-1^),

[*Population Density*](*r*) = population density in region *r* (people m^-2^),

[*Single Particle Infection Probability*]_*ref*_ = reference probability that an individual will become infected after being exposed to a single particle. This term includes the individual’s breathing rate. (m^3^ s^-1^ particle^-1^), and

[*Total Particles Released*] = total number of particles released into the atmosphere (particles).

### 3.2. Relative Infection and Disease Probability

Although disease outbreak assessments are often based on absolute risks, key outbreak response parameters may not be known. In this setting, relative infection probability metrics can be useful. **Equation 3** can also be used to model either relative probabilities of infection or disease. When the source of airborne particles and, separately, the impacted regions are similar (e.g., *r* and *r*_*ref*_ are both residential areas and *r*_*new source*_ and *r*_*ref source*_ are both office areas), **Equation 3** provides the theoretically expected ratio of regional infection probabilities. *Note that* ***Equation 3*** *does not require any information on the release amount per person, the infectivity of individual particles, or the adjustment factor (variation of exposure and population sensitivity)*. When there is a single source region (i.e., *r*_*new source*_ = *r*_*ref source*_), **Equation 3** allows the infection probability of different downwind regions to be compared. When a clear case of region-to-region airborne disease transmission has been identified, **Equation 3** could be used to estimate the infection (or disease) rates for new sources and receptor regions. *We note that this relative infection/disease probability metric is distinct in form from the traditional “relative risk” metric which is defined as the ratio of infection (or disease) incidence between exposed and baseline (ideally non-exposed) populations*.

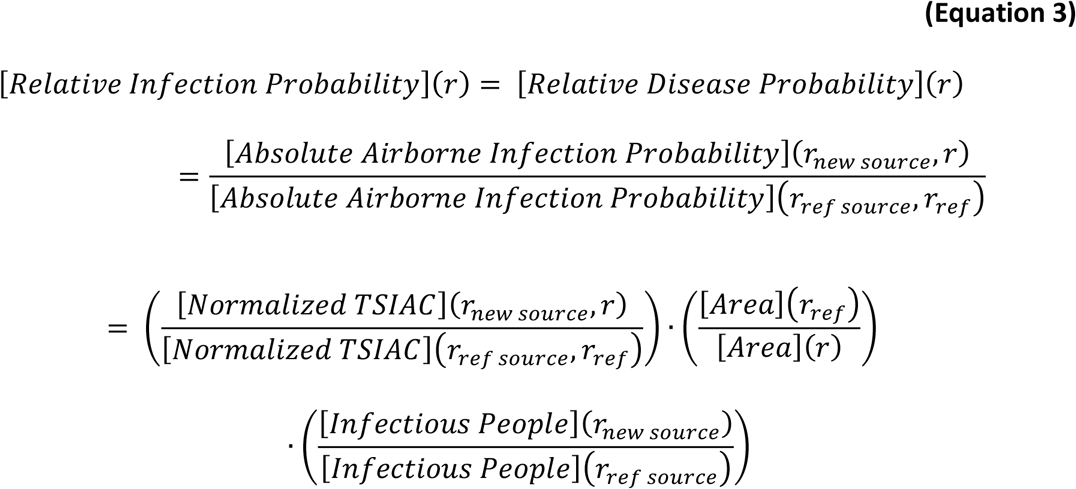

where

[*Infectious People*](*r*_*source*_) = number of people emitting infectious particles in source region *r*_*source*_. (people),

[*Refactive Disease Probability*](*r*) = ratio of the region *r* disease probability to reference region *r*_*ref*_ disease probability. (dimensionless), and

[*Refactive Infection Probability*](*r*) = ratio of the region *r* infection probability to reference region *r*_*ref*_ infection probability. (dimensionless).

The equations presented in the prior sections and derived in **Supplemental Material C: General Theory** focus on infection probability. However, not all infections result in disease. In addition, other metrics, such as the probability of needing medical resources or economic impact, may also be of interest. It is straightforward to adapt the prior equations to any metric in which the probability of an individual’s response to a single particle exposure can be calculated by multiplying (a) a reference response probability by (b) a linear scaling factor (i.e., a metric specific adjustment factor). While the scaling factor may take any value and may vary by individual, the value for a specific individual cannot change. The adjustment factors can vary by metric. When these conditions are met, the relative incidence, i.e., **Equation 3**, of multiple metrics of interest, such as infection and disease, are the same. The **Equation 3** disease spread estimates could also be used to assess importance of other disease transmission pathways for the new receptor region, see **Equations 1** and **4**.

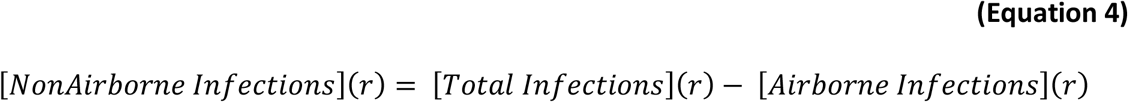

where

[*NonAirborne Infections*](*r*) = number of infected individuals in region *r* that were infected by the transmission pathways OTHER than airborne. (people), and

[*Total Infections*](*r*) = number of infected individuals in region *r*. (people).

### 3.3. Indoor Airborne Particle Dynamics

Infectious airborne particles can be emitted within a building. Once airborne, these particles have the potential, among other fates, to (a) be inhaled and infect people within the building or (b) exit the building, enter the outdoor atmosphere, and be transported downwind. To provide context for later discussion, we have extended prior work [30] and derived **Equations 5** to **7**, which estimate key parameters. ^7^ **Supplemental Material D: Indoor Particle Dynamics** derives these, and other, equations. These equations do not assume, but are compatible with, rare (single particle) exposures.

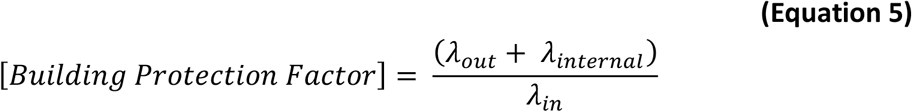

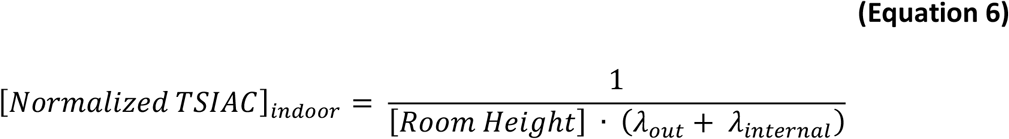

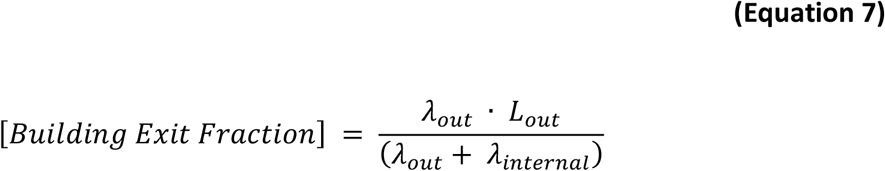

where

*λ*_*in*_ = the rate at which outdoor airborne particles enter the building – typically via infiltration or ventilation. Includes losses that occur during transport from outdoor to indoor. (h^−1^),

*λ*_*internal*_ = the rate at which indoor particles are lost within the building. This term includes both physical losses, such as deposition to indoor surfaces, and infectivity losses while the particle is airborne (h^−1^),

*λ*_*out*_ = the rate at which indoor particles exit the building. (h^−1^),

[*Building Exit Fraction*] = fraction of material released within a building that exits the building and enters the outdoor atmosphere. (no units),

[*Building Protection Factor*] = ratio of the outdoor to indoor exposure. Similar to sunscreen and personal protective respirator rating systems, higher protection factor values indicate lower exposures and thus increased protection. (protection factor),

*L*_*out*_(*particle size*) = the fraction of indoor particles lost while exiting the building. (dimensionless),

[*Normalized TSIAC*]_*indoor*_ = indoor time and space integrated air concentration assuming a single particle is released. (s m^-1^), and

[*Room Height*] = height of building living space. (m).

## 4. Results

This section applies the previously derived theory to estimate regional-level metric values. First, upper bound estimates are provided for both individual person level and regional level infection probabilities for locations downwind. These results assume that infectious particles are emitted from a single, non-moving source.^8^ Second, the relative infection and disease probabilities as a function of downwind distances are provided. The modeling examples provided here (a) assume 1 μm AD particles and no airborne loss of infectivity and (b) consider a wide range of common weather conditions.^9^ Other scenarios, including those with varying particle and environmental parameters, are briefly discussed for context. Third, the relative importance of within building and downwind (both indoors and outdoors) infections is illustrated.

In this section we also compare the modeled predictions for relative infection probabilities (**Equation 3**), with existing observational data from disease outbreaks which have a major airborne transmission pathway. This is not a formal theory validation but is a useful initial assessment of theory predictions compared to data from real-world events. As the primary data collected by the investigators were not available, existing data were abstracted from journal article tables and figures, see **Supplemental Material F: Outbreak Model-Measurement Comparison**. The scope and detail of the published observational data varied between studies which limited the comparisons. In general, the outbreak disease case ascertainment method was notified clinical cases (passive surveillance) and so a fraction of diagnosed cases and any undiagnosed cases were missed. Detailed sub-regional population demographics were not presented, and so adjusted age-specific disease incidence rates could not be estimated. Disc- specific disease incidence rates were not available in some studies. Also, true background disease incidence rates were not reported. Multiple transmission pathways are common in infectious diseases. Most of these studies evaluated that possibility, but one study did not [37].

All studies showed a clear pattern of high disease incidence rates close to an exposure source with a rapid decrease in disease incidence with increasing distance thereafter. **Figure 4** demonstrates this behavior using the Q Fever data. This attack rates shown in **Figure 4** likely underestimate the actual values, see the *5*.*2. Background Disease Incidence* section below.

**Figure 4.**
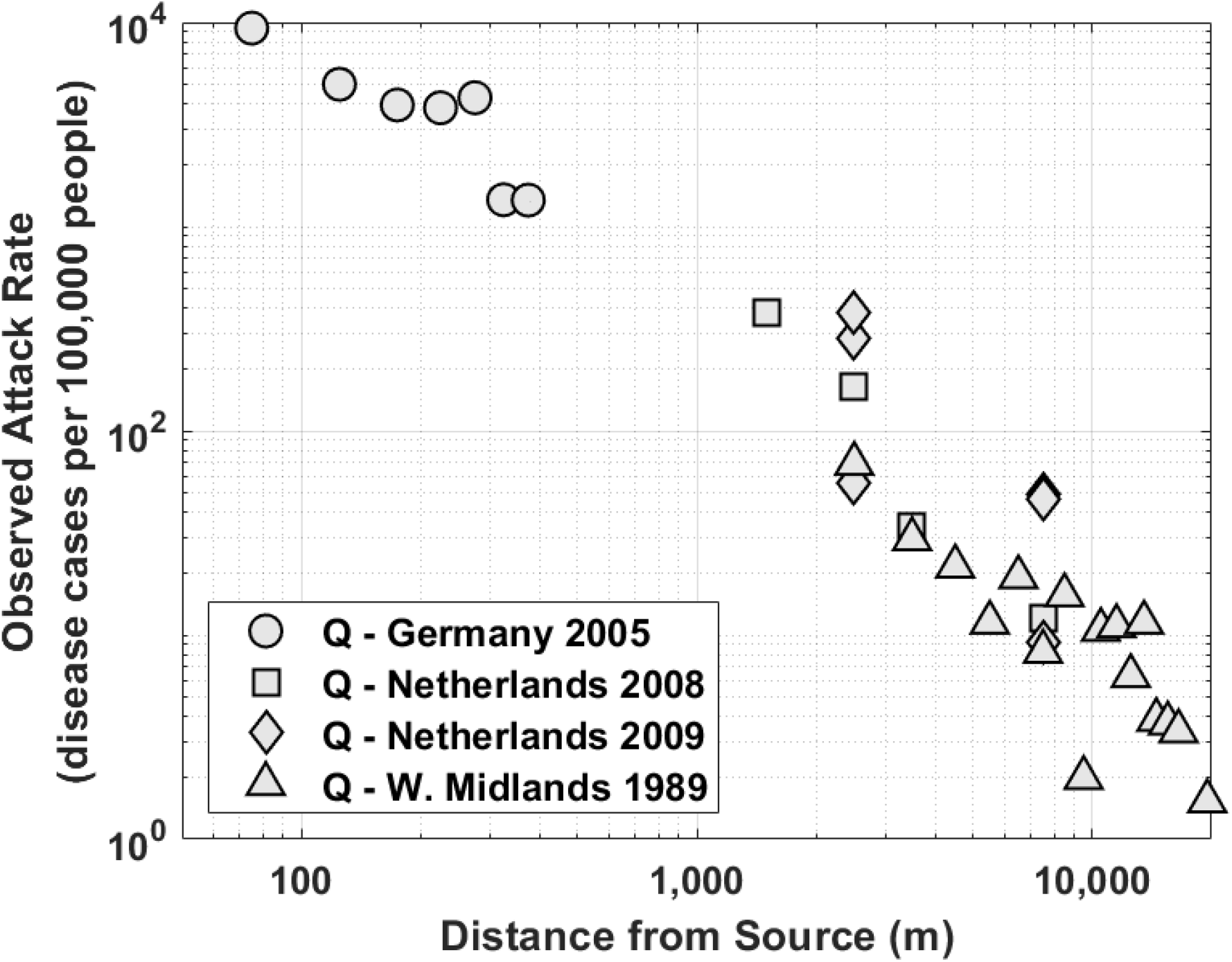
Observed Q Fever attack rate as a function of distance as published in airborne infectious disease outbreak studies. The distance downwind corresponds to the radial distance from the source to the center of a circular band (torus) centered on the source. The band width varies with each point and ranges from 50 m to 5 km. The legend shows the study location, outbreak year, and infectious agent (Q = Q Fever (Coxiella burnetti)).

### 4.1. Upper Bound on Absolute Infection Probability vs. Distance

The top panels in **Figure 5(a-b)** show the predicted upper bound on the absolute airborne infection probability for each particle released to the outdoor atmosphere as integrated (a) over a series of discs and (b) along a series of circle arcs (see **Figure 2**) at distances between 50 m and 20 km from the release point. The bottom panels show the corresponding number of infections in an urban area (assuming 0.01 people m^-2^) for each airborne particle released. For context, about 30% of the New York State, US 2000 Census Tracts have population densities ≥ 0.01 people m^-2^.

**Figure 5.**
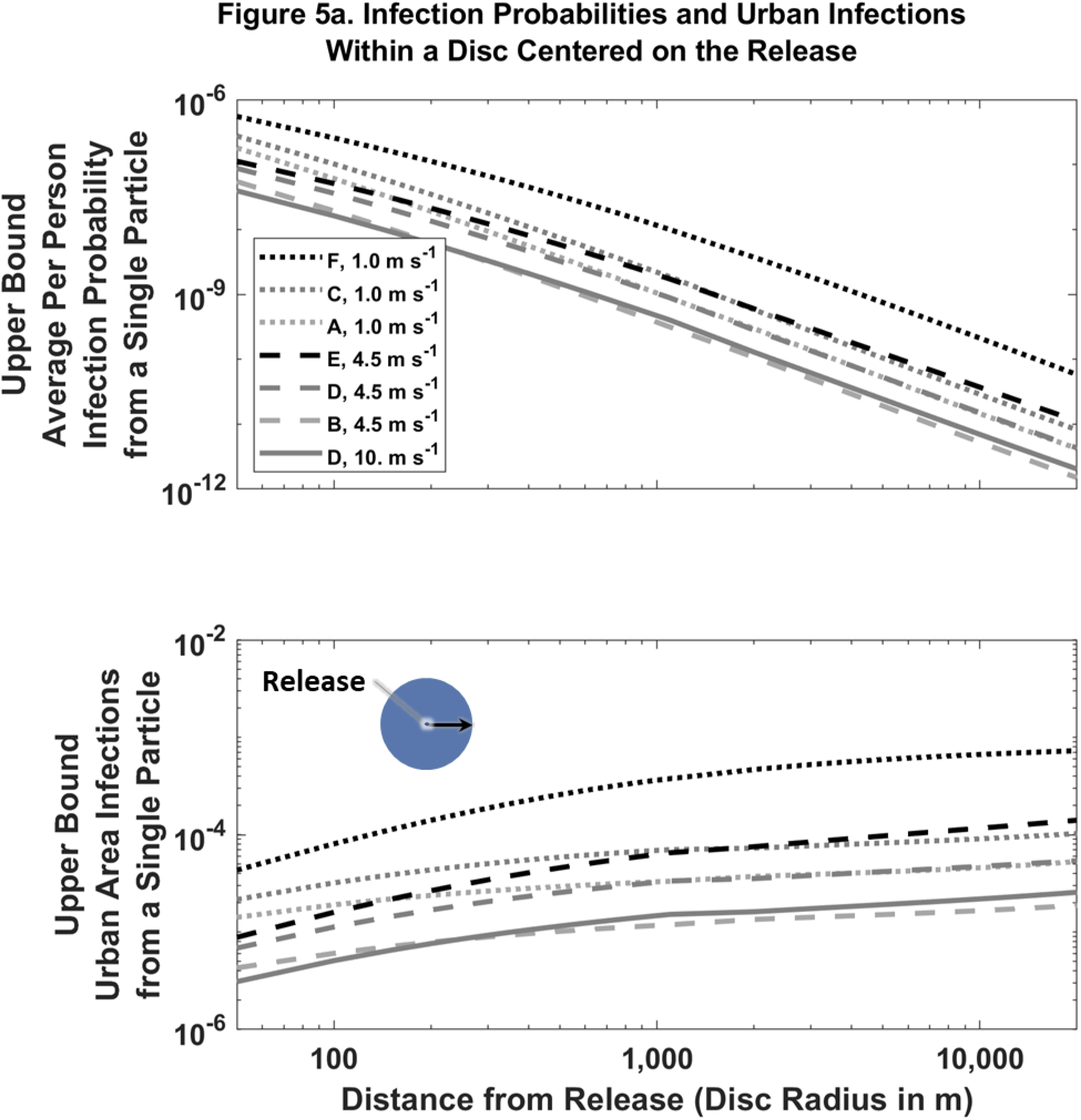
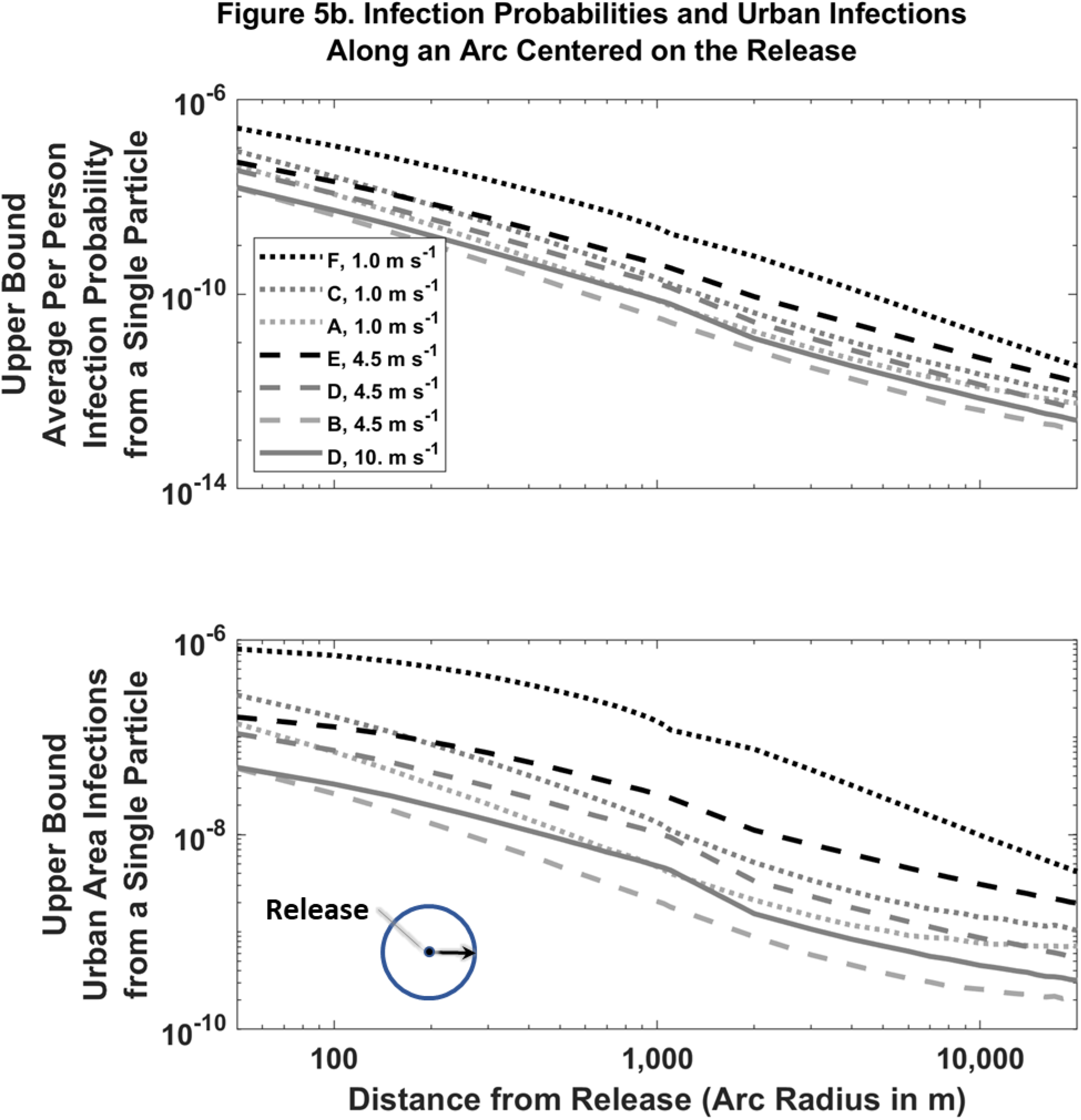
Predicted infection probabilities and infections by distance, wind speed and atmospheric stability for a single airborne particle with no airborne loss of infectivity. Legend box indicates Pasquill-Gifford-Turner atmospheric stability class (A to F) and the 10 m agl wind speed. Individual person infection probability (top panels) is dimensionless. Urban area infections (bottom panels) assume a uniform population density of 0.01 people m^-2^ and has dimensions of people (disc) or people m^-1^ (arc).

The results shown in **Figure 5** were derived from **Equations 1** and **2** and **Table E2** from **Supplemental Material E: Outdoor Normalized Time and Space Integrated Air Concentrations**. We present here the results for one important class of possible weather scenarios, where constant light, moderate, and strong winds (1, 4.5, and 10 m s^-1^, respectively) would transport an airborne particle 1 km downwind in about 17, 4, and 2 min, respectively.^10^ Thus these results are reasonable when losses are not significant on the timescale of minutes to hours.^11^

The absolute infection probability varies over several orders of magnitude and decreases rapidly with distance. Broadly speaking, the infection probability increases with (i) decreasing wind speed and (ii) increasing atmospheric stability (increasing stability corresponds to decreasing rates at which material dilutes within the broader atmosphere). The actual infection probability will be smaller if not all inhaled particles cause infections and/or the individual is indoors or otherwise protected. In the latter case, downwind infection probability estimates can be determined by dividing the values shown in **Figure 5** by a building protection factor, e.g., [30]. We note that the expected number of infections linearly scales with both the total number of particles released and the population density.

When the airborne loss rate is significant on the timescale of minutes, e.g., 10 h^-1^, the modeled relative infection incidence increases with wind speed at distances greater than 1 km, see **Figure 6**.

**Figure 6.**
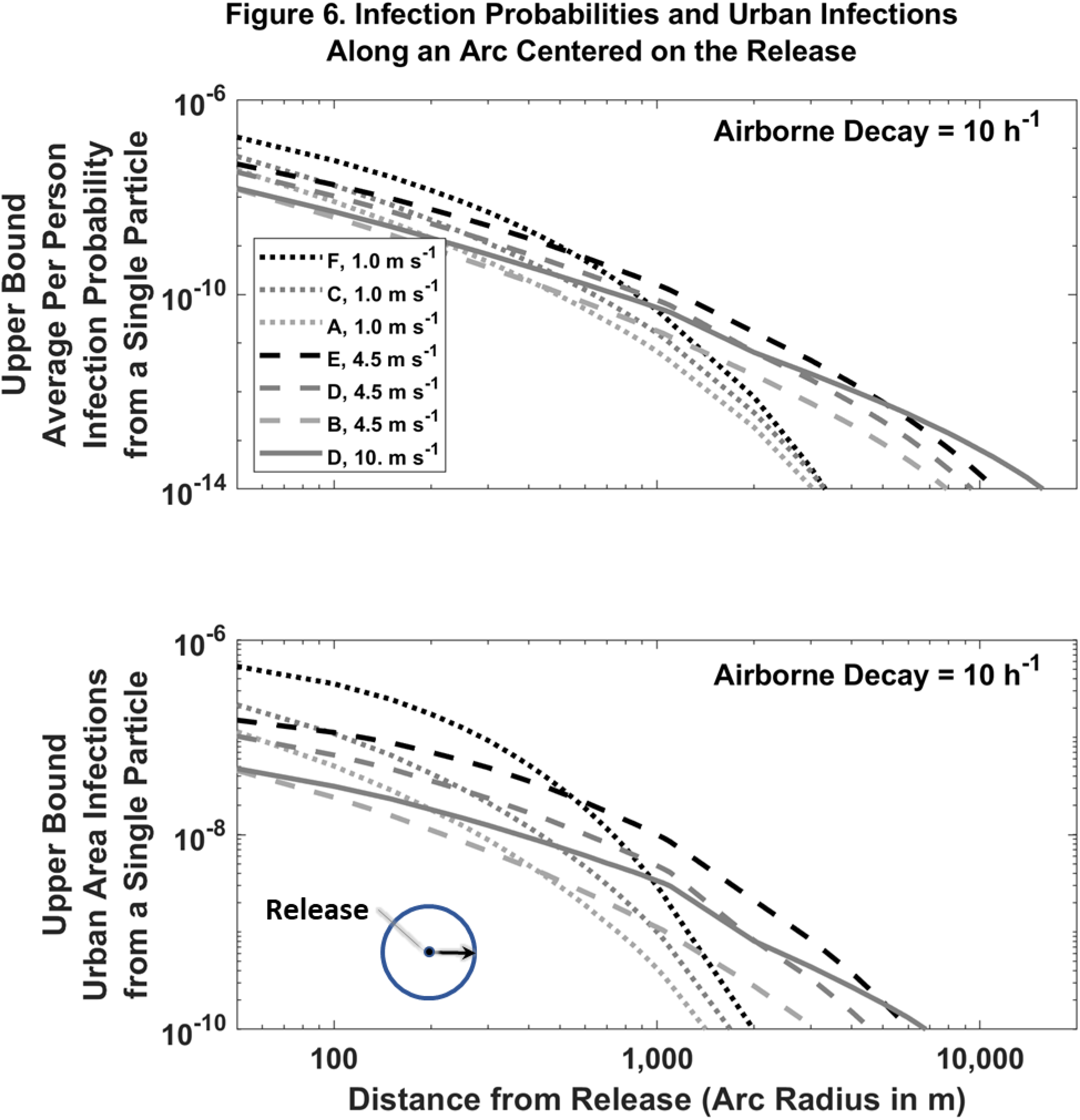
Predicted infection probabilities and infections by distance, wind speed and atmospheric stability for a single airborne particle with a 10 h^-1^ airborne loss of infectivity. Legend box indicates Pasquill-Gifford-Turner atmospheric stability class (A to F) and the 10 m agl wind speed. Individual person infection probability (top panels) is dimensionless. Urban area infections (bottom panels) assume a uniform population density of 0.01 people m^-2^ and has dimensions of people (disc) or people m^-1^ (arc).

### 4.2. Relative Infection and Disease Probability vs. Distance

**Figure 7a** shows the corresponding relative infection (and disease) probabilities as calculated by **Equation 3** and the upper bound absolute infection probability curves shown in **Figure 5** as integrated over (top panel) a disc and (bottom panel) along a circle arc, both being centered at the common release point (*r*_*new source*_ = *r*_*ref source*_). While the relative infection (and disease) probabilities again decrease rapidly with distance, there is minimal variation with wind speed or atmospheric stability over the range of atmospheric conditions considered.^12^ Hence the relative infection probability metric may be particularly valuable when there is incomplete or entirely lacking information on meteorological conditions, but (a) atmospheric conditions are known to not be materially changing during the time it takes for the infectious particles to travel from the source to the receptor and (b) the particle infectivity is not lost rapidly in the atmosphere. *This relationship is expected to hold even when the exposed population has varying sensitivity to the infecting particle and/or are located inside buildings affording different degrees of protection as long as the distributions of both the infection sensitivity and building protection are similar in both exposure regions*.

**Figure 7.**
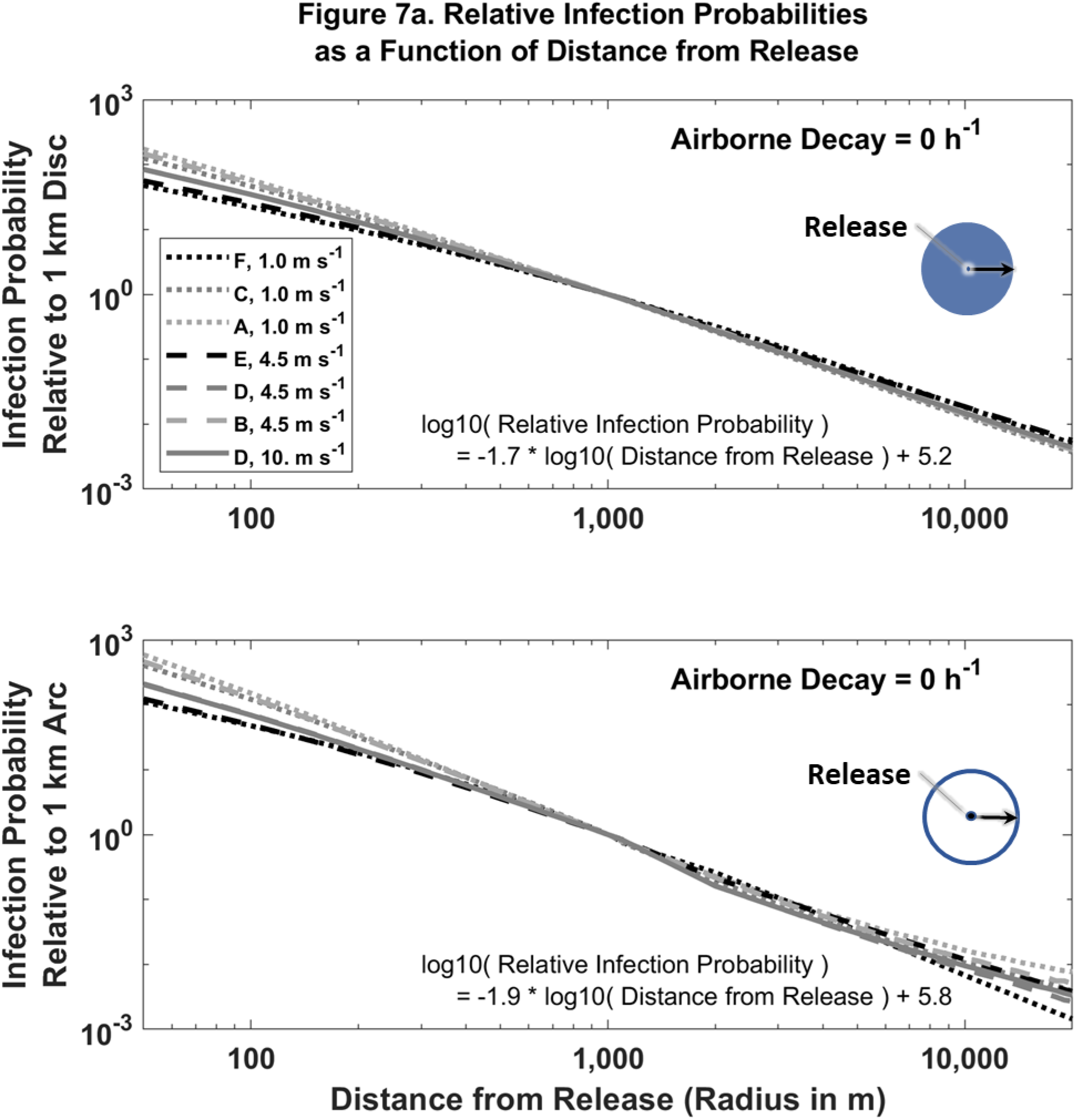
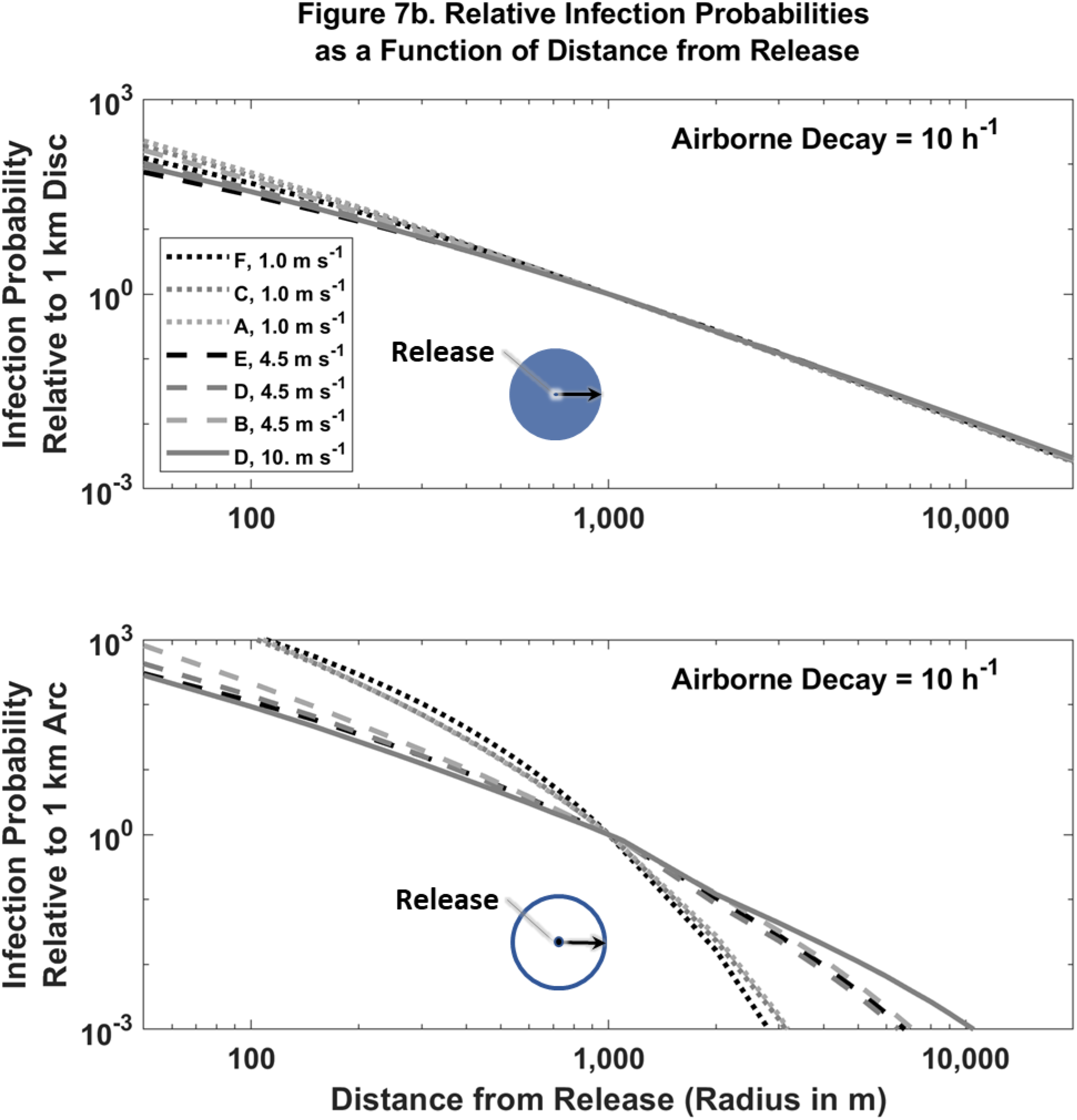
Predicted relative infection probabilities by distance, wind speed and atmospheric stability for a single airborne particle with (a) 0 h^-1^ and (b) 10 h^-1^ airborne infectivity loss rates. Legend box indicates Pasquill-Gifford-Turner atmospheric stability class (A to F) and the 10 m agl wind speed. Relative infection probability is dimensionless.

When the airborne loss rate is significant on the timescale of minutes, e.g., 10 h^-1^, the modeled relative infection incidence for circle arcs increases with wind speed, see **Figure 7b**. Notably, the slope of log_10_(distance from source) vs. log_10_(relative infection rate) is smaller than the no (0 h^-1^) airborne loss rate case. Due to the rapid decrease in airborne infection probability with distance, the corresponding plots for relative infection incidence for a disc are nearly identical to that shown in **Figure 7a**.

### 4.3. Model Prediction – Outbreak Data Comparison

In this section, we compare previously published, major airborne outbreak data against relative probability values calculated using **Equation 3**. First we use data from Q Fever (Coxiella burnetti) [37], [40]–[42], Legionnaire’s disease (Legionella pneumophilia) [43], [44], and Valley Fever (Coccidioides immitis) [45], [46] outbreaks. We note that these human pathogens have reproductive number (R_0_) values less than 1 (Case 1 diseases) and so they have little or no potential for person-to-person transmission, simplifying the airborne disease modeling.^13^ Subsequently we use results derived from veterinary disease outbreak studies of Highly Pathogenic Avian influenza (HPAI) [47]–[49], Foot and Mouth Disease (FMD) [50], [51], and Classical Swine Fever (CSF) [52]. These diseases have reproductive numbers (R_0_) greater than 1 (Case 2 diseases) and so have the potential for secondary epidemic spread (they also are known to have multiple disease transmission pathways). The modeled values were determined using **Equation 3** and the neutral (D) stability, moderate wind (4.5 m s^-1^ at 10 m agl) modeling results presented in **Supplemental Material E: Outdoor Normalized Time and Space Integrated Air Concentrations**. Other than the source location, definition of the base and reference areas, and the assumption of a single source (*r*_*new source*_ = *r*_*ref source*_), no disease or outbreak specific information was used in the modelling.

**Figure 8** compares the predicted model relative disease probabilities to the observed relative incidence of clinical disease reported for Q Fever, Legionnaire’s disease, and Valley Fever outbreaks. The modeled and measured values are reasonably well correlated, r^2^ = 0.86, and close to the 1:1 line.^14^ The base and reference areas vary by comparison point and are specified in **Supplemental Material F: Outbreak Model-Measurement Comparison**. Some data points from these studies were not used either due to (a) the low number of observed disease cases^15^ or (b) because they were adjacent to the source and did not exhibit the expected decrease in disease rate with distance. The specific data points used and omitted are indicated in the **Supplemental Material F: Outbreak Model-Measurement Comparison**.

**Figure 8.**
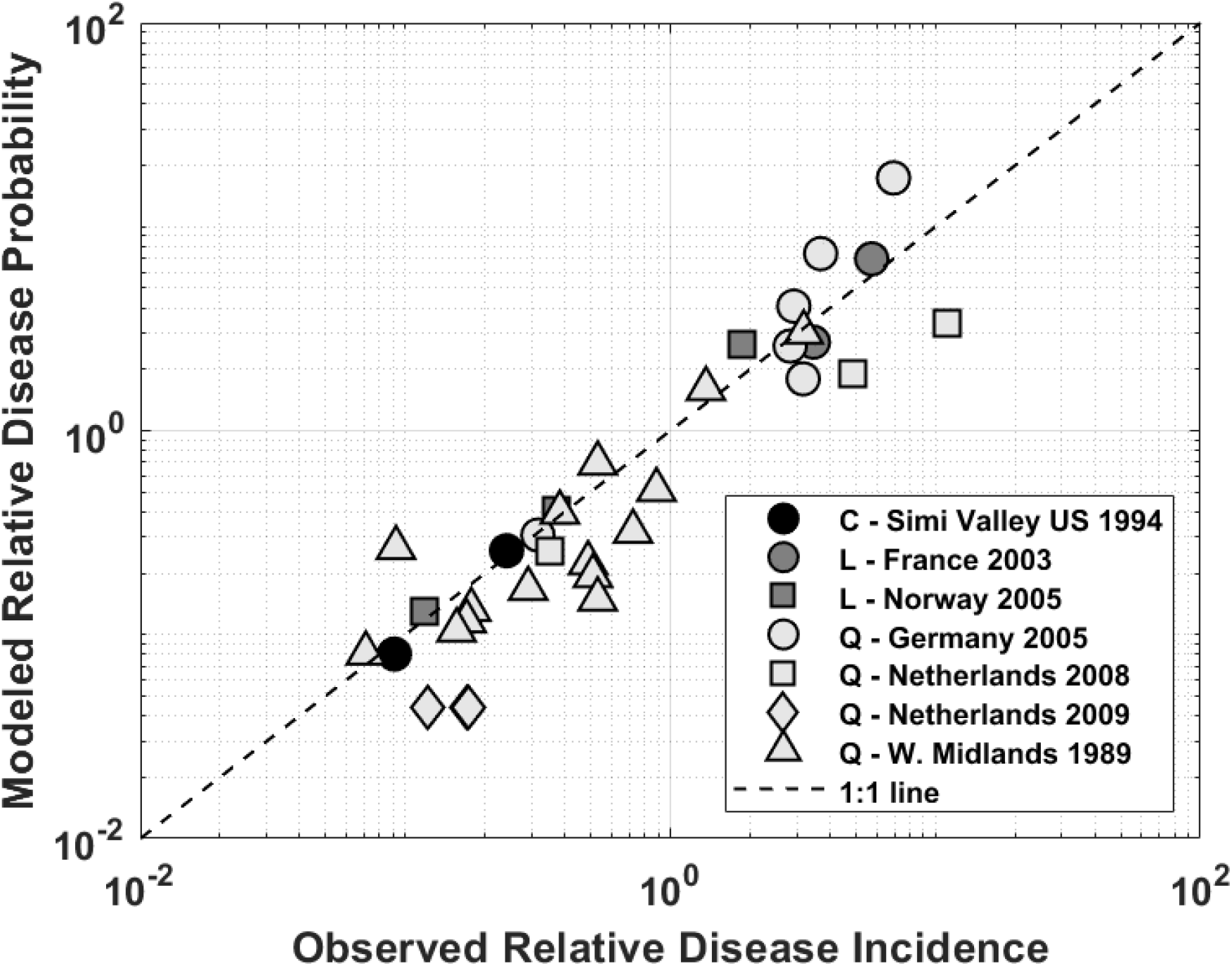
Modeled Relative Disease Probability Compared to Observed Relative Disease Incidence as published in airborne infectious disease outbreak studies. The dashed line is the 1:1 line which indicates perfect agreement. The legend box shows the study location, outbreak year, and infectious agent (Q = Q Fever (Coxiella burnetti); L = Legionnaire’s Disease (Legionella); C = Valley Fever (Coccioidomycosis)).

**Table 2** demonstrates that our prediction^16^ of the slope of log_10_(distance from source) vs. log_10_(relative infection rate) ≤ −2 is consistent with both (a) the previously discussed studies and (b) as previously reported for Highly Pathogenic Avian influenza and Foot and Mouth Disease outbreaks. We note that Classical Swine Fever and Valley Fever do not have sufficient data to formally assess this statement, but the reported values are ±25% of the predicted values.

**Table 2.**
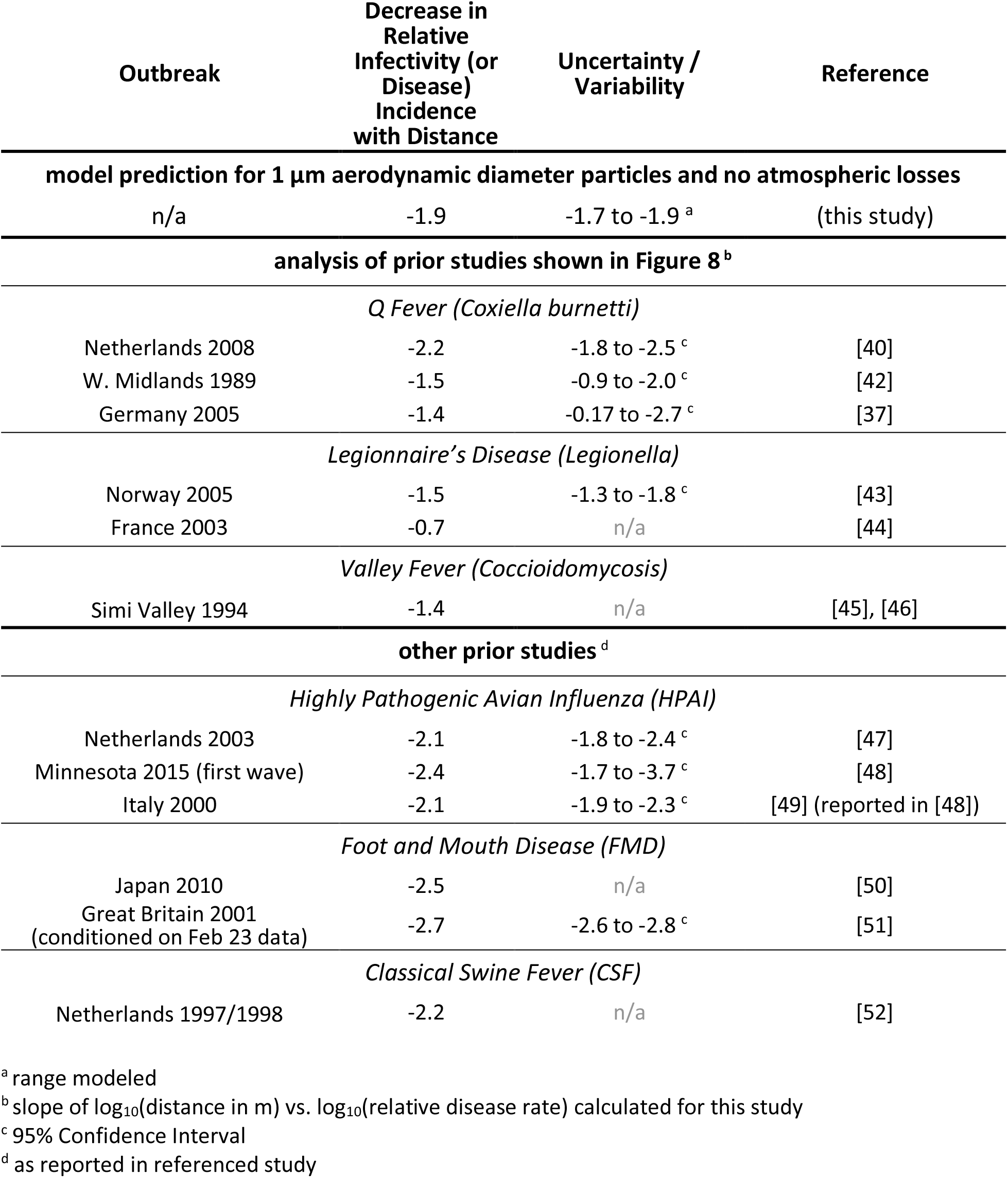
Summary of decrease in relative infectivity (or disease) with distance for selected studies.

For the HPAI, FMD, and CSF cases, the infection rate as a function of distance was determined by the original authors from a set of base equations whose parameter values were determined from the timing of the discovery of each specific disease case, the locations of known infected farms, assumptions about disease incubation periods, the time from infection until detection, certain farm attributes, and (for some studies) varying sensitivity to a given infectious exposure. In contrast to the current analysis, the analyses presented in those studies did not model the physics of disease transmission mechanisms, nor do they assume airborne transmission. In these outbreaks, the infection probability (transmission kernel) was demonstrated to be relatively constant over an initial, short-range geographic extent and then decrease with distance. The latter decrease in infection probability (disease incidence rate) with distance is equivalent to the slope in equation shown in lower panels of both **Figure 7(a-b)**.^17^

The observed FMD disease rate decreases notably faster than that demonstrated in our baseline results, which assume no atmospheric loss of infectivity. The FMD results are, however, more consistent with modeling that includes infectivity losses that are significant on the timescale of the plume transport and we note that FMD virus is in fact known to lose infectivity in the atmosphere [53].

### 4.4. Relative Magnitude of Within Building and Downwind Infections

Physical science considerations suggest that some fraction of airborne indoor particles will exit buildings. For example, we predict that, on average, about 35% of the airborne, 0 hr^-1^ infectivity loss rate, 1 μm particles in US single family homes will be released to the outdoor atmosphere, see **Supplemental Material D: Indoor Particle Dynamics** and [30].^18^ Indeed under some conditions, infections due to airborne transmission could result in similar numbers of people being infected downwind than in the same house as an infected individual – see the example provided in **Supplemental Material G: Infection Estimates**. When the regional population density is notably greater than 0.01 people m^-2^, more people may be infected downwind than within the source building. We note that (a) not all diseases are capable of longer-distance downwind transmission, see *5*.*3. Key Infectious Agent/Disease Characteristics* section, (b) other disease transmission pathways may substantially increase the probability of being infected while sharing the same house as an infected individual, (c) downwind individuals may be indoors, and (d) even for the airborne transmission pathway, the “per-person” infection probability is higher within a given building than it is downwind.

Building operation changes could alter the absolute and relative probability of source building and downwind infections. For example, actions that increase the indoor/outdoor air change rates (*λ*_*out*_, *λ*_*in*_), such as increasing the ventilation, would (a) decrease indoor exposures within the source building, (b) increase the number of infectious particles emitted to the greater atmosphere, and (c) decrease the protection downwind buildings provide their occupants. In contrast, actions that increase the within building loss rates (*λ*_*internal*_), potentially including replacement of existing furnace filters with higher efficiency filters, have the potential to reduce exposures in both the source and downwind buildings.

## 5. Discussion

### 5.1. Characterizing Airborne Disease Spread

In a disease outbreak situation, it may be necessary to establish whether airborne disease transmission is occurring. Laboratory studies to identify the suspect pathogen and the known pathogen characteristics (see the *5*.*3. Key Infectious Agent/Disease Characteristics* section above) are valuable in this regard. Epidemiological factors may indicate a spatio-temporal signal consistent with airborne transmission, specifically (a) new infections that have no contact with a known infectious individual, (b) correlation between new infections and key weather conditions, and (c) the presence of disease “sparks” outside the regions of known infection. This work suggests an additional method, where the geographic distribution of observed disease cases agrees with that predicted by **Equation 3**, i.e., log10(distance from source) vs. log10(disease rate) is linear with a slope ≤ −2. Comparison to prior outbreak data suggests that this airborne transmission signal may be readily apparent, especially in cases where there is a single emission source. However, we note that it could be (a) mimicked by other infection transmission pathways if they are also correlated with distance from the source, (b) obscured by other disease transmission pathways, and (c) challenging to identify when there are multiple, geographically separate emission source regions. However, other disease transmission pathway(s) should be considered when the **Equation 3** predicted signal is absent.

The theory and modeling presented in this paper define the single particle airborne disease transmission kernel. This kernel corresponds to the set of lower probability, longer-distance airborne transmission events. These could contribute to large scale disease spread by “sparking” unexpected disease outbreaks distant from an emission source. The impacts of these longer-distance transmissions can be high for diseases that can cause many secondary infections (i.e., Case 2 diseases). In the plant biology and biosafety literature, it is well understood that these low probability, long distance (100’s of km) spread events do occur and are a primary mechanism of long-term species colonization of new environments and plant pathogen spread [54]–[57]. Indeed, unexpected long-distance disease sparks are a particularly well-documented problem in Foot and Mouth Disease of ruminants, where prior investigators have noted “Airborne spread is the most important mechanism of uncontrollable [disease] spread and although this is not a common event, when it occurs the speed and extent of spread can be spectacular” [58].

### 5.2. Background Disease Incidence

Due to the predicted finite, but characteristically low, individual-person infection probability rates in the general population, airborne diseases that present similarly to other common, non-airborne diseases may be prone to being under diagnosed and under reported. While an individual event may not affect many people, the cumulative impact of routine, small-scale events could pose a significant health burden. More accurately defining the airborne disease burden may be important since with appropriately targeted programs, there is a potential to reduce the currently existing US population airborne disease burden.

As an illustration, Q Fever is a disease with a well-established airborne transmission pathway and disease control measures exist [59]. Q Fever is often considered a rare disease; however, it is known that Q Fever cases are significantly underreported even though it is a US nationally notifiable disease with mandatory reporting in most states [60], [61]. US nationally representative data from 2003 to 2004 suggest that the overall adult population prevalence of acute, recent, and chronic Coxiella infections is 6 million people, 3% of the general US population [62]. Further analysis of this data indicates the prevalence of Acute Q Fever (Coxiella burnetti) infections may be approximately 2 million people, see **Supplemental Material H: U.S. Coxiella burnetti Infection and Disease Estimates**.^19^ For comparison, during this time period there were 70 Q Fever cases in the national reporting system [64], [65] and in 2017 there were 153 acute and 40 chronic Q Fever cases reported in the US [66]. Disease underreporting rates are similar for other major airborne disease mentioned here. For example, among US hospitalized persons with pneumonia, Legionnaire’s Disease is ten times more common than currently being diagnosed [67].

Accurately estimating background disease rates is also an important step in investigating epidemic outbreaks as these are needed to produce accurate absolute outbreak incidence estimates and relative risks. We note that true population-based background incidence and prevalence rates are unavailable for most diseases. ^20^ This literature gap is a practical matter as analysts often arbitrarily choose the lowest observed incidence rates at a location adjacent to the main body of outbreak cases as a background reference rate. However, as seen above, infections at these distances might be due to long-distance airborne transmission as this work suggests that once an infectious particle is airborne, there is a non-zero probability of infections downwind. Misclassification of background disease rates could result in inaccurate (a) estimation of the true spatial extent of an outbreak and (b) dose-response modeling [69]. There is also risk of undiagnosed cases and secondary disease spread.

### 5.3. Key Infectious Agent/Disease Characteristics

Only select infectious agents can transmit disease via inhalation of a single particle, particularly over longer (>5 m) distances. Based on **Equation 2**, the key infectious agent/disease characteristics that determine the likelihood of airborne transmission are (a) the number of infectious particles emitted, (b) the degree of infectivity of individual particles, and (c) the loss of particle infectivity while airborne. **Table 3** summarizes these characteristics.

**Table 3.**
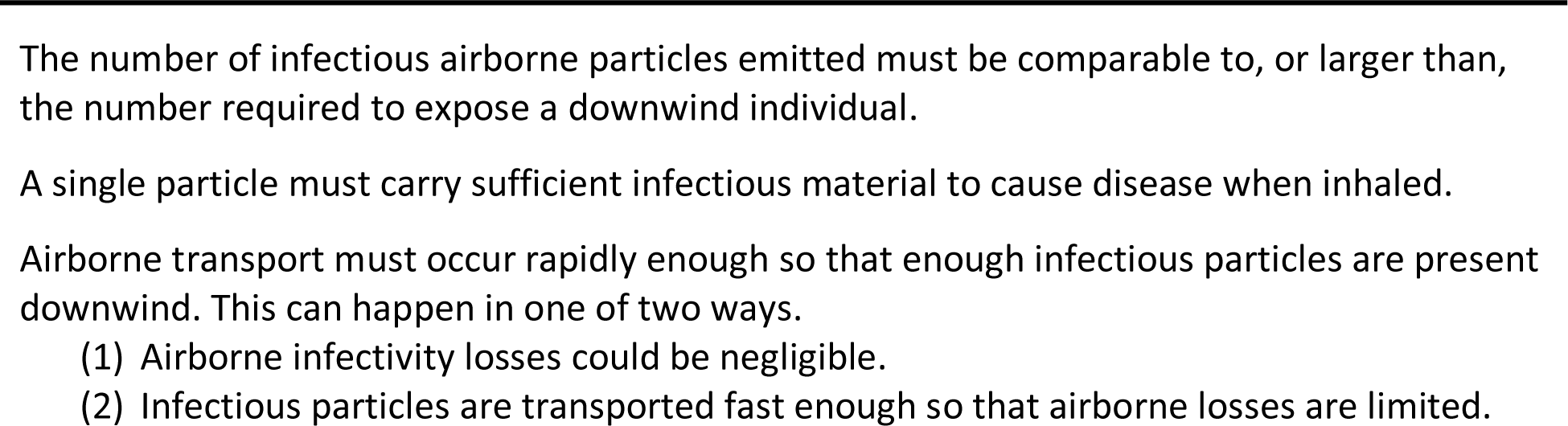
Summary of Factors Relevant to Single Particle Airborne Disease Transmissio

#### 5.3.1. **Number of Infectious Particles Emitted:** [*Total Particles Realeased*]

With respect to Case 2 (R_0_ > 1) human diseases, human-origin particles can comprise a significant fraction of indoor airborne particles, e.g., [70], [71], and ill individuals are known to emit infectious particles through coughing, sneezing, breathing, and/or shedding of infected skin or mucosal cells [71]–[74]. While it is challenging to develop reliable estimates of the number of infectious particles emitted by an ill individual, working estimates of total human respirable (1 to 5 μm AD) particle emissions are available from published studies. For context, up to 10^6^ airborne particles can be emitted per sneeze (although there is some debate on the fraction of these particles that are respirable) [75], [76]; 10^3^ to 10^5^ respirable diameter particles are emitted both in a cough and for short (15 to 30 minute) periods of breathing and talking [33], [73], [77]–[80]; and 10^7^ skin cells are shed per day (of which a fraction become airborne respirable particles) [53], [81]. We note that sick individuals may emit more particles than healthy individuals and some individuals can emit an order of magnitude or more particles than the average person, e.g., super-spreaders [77], [82]. For context, we estimate 10^4^ to 10^6^ environmentally stable infectious particles emitted indoors are required to infect one downwind person in an urban area (0.01 people m^-2^). ^21^

#### 5.3.2. Single Particle Infectivity: [Single Particle Infection Probability]

Individual particles need to carry enough infectious material to cause disease. For some disease-causing infectious agents, this is a given as they possess a very low infectious inhalation dose. One example is Coxiella burnetii, which causes Q Fever, where a single bacterium can potentially cause infection in humans [15], [83].^22^ For other infectious agents, multiple individual agents are required for infection. Theoretically, the required number of infectious agents may be transported on the surface of, or within, larger carrier particles.^23^ From physical considerations alone, approximately 100 1 μm AD infectious agents (a common bacteria size) could fit within a single 5 μm AD particle.^24^ For smaller agents, such as viruses, the number of agents in a carrier particle could be much larger.

#### 5.3.4. Loss of Particle Infectivity While Airborne: Affects [Normalized TSIAC]

Some infectious agents – including important human pathogens and economically important plant disease pathogens – are environmentally hardy and are known to readily transmit disease over long distances - causing significant disease outbreaks [57], [89]–[95]. However, other agents, including many human and veterinary pathogens, are more fragile and their potential for long-distance airborne disease transmission is more controversial. For some of these pathogens, the rate at which infectivity is lost in the atmosphere can be high and depends, in part, on the specific virus or microorganism and strain, microbial state (e.g., vegetative cell vs. spore), environmental conditions (e.g., temperature, humidity, insolation), and particle composition [96]–[98]. For context, reported airborne loss rates for a collection of pathogens (Category A select agents) range from undetectable to up to about 5 h^-1^ when expressed as a first order (base e) loss rate [98]. Observed loss rates are not always first order, as it is common to observe an initial “die-off” followed by a slower loss rate for a relatively hardy subpopulation, and so an alternate metric is also informative. These same studies reported that time required for 90% of the original material to be lost is at least 15 min [98].

The mere existence of notable atmospheric infectivity loss rates may not, in and of itself, prevent agents from transmitting disease via the atmosphere. Indeed, many of the examples provided in the medium and long range subsections of the **Supplemental Material A: Airborne Disease Transmission Literature Review** are known to have significant airborne loss rates. However, our modeling results indicate that even when infectivity is lost rapidly, 10 h^-1^, infection probabilities are minimally impacted several hundred meters downwind, see **Figure 7b**. Longer distance airborne transmission of infectious agents with notable atmospheric loss rates is also facilitated by the following:

> First, environmental conditions (e.g., insolation, temperature and humidity) are not fixed and so airborne disease transmission may simply be limited to when conditions are favorable for agent atmospheric survival. As one example, viruses or microorganisms that are sensitive to solar (UV) radiation would not be affected at night. ^25^ Similarly, UV exposure varies significantly by latitude, season, time of day, cloud cover, air pollution, airborne dust, indoors vs. outdoors, etc.. Similar considerations hold true for other common environmental variables including humidity and temperature.
>
> Second, even if there is a significant airborne infectivity loss rate, disease transmission might occur if the outdoor atmospheric transport is rapid relative to the loss rate. For many common weather conditions, downwind transport of airborne particles may indeed be rapid - with kilometer scale transport occurring in 15 minutes or less.^26^ When airborne infectivity loss rates are significant, infection rates are predicted to be higher at higher wind speeds, see **Figure 6**. For context when airborne infectivity loss rates are negligible, infection rates are predicted to be higher during the night (stable atmospheric conditions) and at lower wind speeds, see **Figure 5**.

### 5.4. Potential Future Efforts

This paper develops new theory for airborne disease transmission; models potential physical exposures; provides a selective literature review of airborne disease transmission; and provides an initial model comparison to published epidemiological data of the predicted relative infection/disease prevalence as a function of downwind distance for a single source region. However, the key variables and modeling parameters presented here are necessarily general and/or limited to archetypical cases.

More detailed analyses could provide more accurate predictions for specific scenarios and – coupled with laboratory, epidemiological, and/or field measurements – could also be used to validate the work presented in this paper. These detailed analyses could consider:

a. specific agents, particle properties, environmental conditions, and population demographics;^27^
b. mobile populations;
c. particle distributions (including sizes greater than 5 μm AD);
d. improved exposure modeling particularly for (i) outdoor exposures < 50 m and > 20 km from the source and (ii) indoor exposures where which the indoor air is not well mixed, e.g., < few meters from the source; and
e. other disease transmission pathways, such as droplets, fomites, and direct/indirect contact including the case in which outdoor airborne particles travel indoors, deposit on indoor surfaces, and pose an on-going contact or resuspension hazard.

We note that special, at-risk, populations, such as (a) children < 5 years of age, (b) the elderly, or (c) immune compromised persons, may have higher infection disease risks relative to the general population, e.g., the examples of influenza risks or Aspergillus infections in immune compromised people.

After appropriate scientific peer review and validation, applications of this research may help enhance airborne disease outbreak management and chronically occurring airborne exposure management. Potential future research areas are briefly highlighted here, although we note that such work should be conducted with due consideration of (a) the potential impact on, or interaction with, other disease transmission pathways^28^ and (b) other factors, such as food, water, medical needs, resource limitations, communication capabilities, expected population compliance, and impending hazards (e.g. fire).

> The theory developed here could inform future updates to remediation clearance and biosafety criteria, i.e., how clean is clean enough. As one example, the interim B. anthracis clearance strategy suggests a best-practice clearance goal of no detectable viable spores on any environmental sample [100]. This goal was selected, in part, because the dose (exposure) response (infectivity or disease) relationship for B. anthracis is not sufficiently understood to adequately assess disease risk. The theory presented in this paper, coupled with estimates of how much surface contamination could become airborne, could bound the downwind risk – even in the absence of any data on a dose-response relationship.
>
> Suspectable populations could be protected when high-risk environmental conditions are forecasted [101]–[103]. This response paradigm is in common use for other airborne hazards where evacuation^29^ and shelter (see next paragraph) are routinely used protective actions [31].
>
> Sheltering has the potential to reduce airborne disease spread, both within a single building as well as further downwind. Sheltering actions may include changes to population locations (e.g., shelter in place) as well as changes to building stock and operations (e.g., weatherization programs to reduce indoor/outdoor air change rates; improved particulate filtration). The use of mass (“cleaner-air”) shelters, such as those currently used to reduce population level outdoor wildfire smoke (air quality) exposures, should be carefully considered in infectious disease settings as some diseases can rapidly spread when many people are in close quarters.

## Data Availability

Data provided in supplemental materials.

## 6. Acknowledgements

The authors are grateful to their family for their support and enduring patience. The authors also thank Ron Baskett and John Nasstrom of the Lawrence Livermore National Laboratory for their considerable assistance in developing the Supplemental Material B: Key Atmospheric Transport and Dispersion Modeling Concepts section. Furthermore, the authors acknowledge the assistance of Brooke Buddemeier, Steve Homann, Brenda Pobanz, Ellen Raber, Antoun Tarabay, and Ken Turteltaub of the Lawrence Livermore National Laboratory; Richard Sextro of Lawrence Berkeley National Laboratory; David Brown of Argonne National Laboratory; Dev Jani of the US Department of Homeland Security, Countering Weapons of Mass Destruction Office; Frederick Miller of the National Institute of Health; William Rhodes of Mele Associates, Inc.; Benjamin Arnold of the University of California, San Francisco; Joseph Chang of the RAND Corporation; Michael Mastrangelo of the University of Texas Medical Branch; and Mark Nicas of the University of California, Berkeley during the development of this manuscript. Financial support was provided by the US Department of Homeland Security for related early efforts and the Lawrence Livermore National Laboratory for facilitating the review of this material. Finally, the authors would like to thank Freepik for the icons, designed by Freepik from Flaticon (www.flaticon.com), used, in part, in the development of Figures 1 and D1.

This document was prepared as an account of work sponsored by an agency of the United States government. Neither the United States government nor Lawrence Livermore National Security, LLC, nor any of their employees makes any warranty, expressed or implied, or assumes any legal liability or responsibility for the accuracy, completeness, or usefulness of any information, apparatus, product, or process disclosed, or represents that its use would not infringe privately owned rights. Reference herein to any specific commercial product, process, or service by trade name, trademark, manufacturer, or otherwise does not necessarily constitute or imply its endorsement, recommendation, or favoring by the United States government or Lawrence Livermore National Security, LLC. The views and opinions of authors expressed herein do not necessarily state or reflect those of the United States government or Lawrence Livermore National Security, LLC, and shall not be used for advertising or product endorsement purposes.

Lawrence Livermore National Laboratory is operated by Lawrence Livermore National Security, LLC, for the U.S. Department of Energy, National Nuclear Security Administration under Contract DE-AC52-07NA27344.

## 7. Attestations

### Ethics statement

No humans or animals were used this in work

### Data accessibility

No primary data were used in this analysis. The referenced supplemental materials have been provided.

### Author’s contributions

Michael Dillon (MBD) was responsible for the study concept and design, theory development, modeling, and data analysis.

Charles Dillon (CFD) was responsible for the epidemiologic content (including identification of previously reported outbreak data) and, jointly with MBD, developed the Supplemental Material A: Airborne Disease Transmission Literature Review Discussion and Supplemental Material H: U.S. Coxiella burnetti Infection and Disease Estimates sections.

Ron Baskett (RB) and MBD developed the Supplemental Material B: Key Atmospheric Transport and Dispersion Modeling Concepts (i.e., RB was a co-author on this section). John Nasstrom provided significant feedback and suggestions.

MBD and CFD both participated in literature searches, manuscript drafting, and revision through its many versions.

All authors give approval for release.

### Funding statement

MBD was supported, in part, by the US Department of Homeland Security for early efforts.

CFD was self-supported.

## Supplemental Material A: Airborne Disease Transmission Literature Review

Air is not a sterile medium, as initially demonstrated in the early 19^th^ century experiments of Louis Pasteur. Bacteria and fungi are ubiquitous in the atmosphere and reach concentrations of about 10^4^ and 10^3^ cells m^-3^ in air, respectively [104]–[107]. These facts are well understood and elucidated in the field of Aerobiology which has documented the life cycles, including transport and dispersion, of naturally occurring airborne viruses, microorganisms, and bioaerosols [108]– [111]. However, there appears to be much confusion on the potential for airborne disease transmission, particularly at longer spatial scales due likely in part to the dispersed nature of this literature.

To provide the reader context for the main manuscript, this section briefly presents a set of well-documented airborne disease transmission examples on spatial scales ranging from a meter to thousands of kilometers. The specific cases discussed here are from the human disease as well as veterinary and agricultural biosafety^30^ literatures and have multiple studies showing airborne disease transmission. This synopsis is not intended to be comprehensive and we do not cite all relevant literature. We note that [8] is a useful literature review of atmospheric dispersion modeling for infectious diseases, with a focus on human and veterinary studies.

### Near Range (< 5 m)

Near distance airborne disease spread (≤ 5 m) is common and occurs when infected individuals generate large quantities of infectious droplet (and droplet nuclei) particles when coughing or sneezing [33], [112]. Tuberculosis and the Measles virus (Rubeola) have long been known to transmit over this distance [1], [2]. Bordatella pertussis (Whooping Cough), Varicella Zoster virus (Chickenpox), Mumps virus, Rubella virus (German Measles) and Neisseria meningitides (bacterial meningitis) are additional examples [113], [114]. This near range airborne disease spread is known to contribute to the overall disease burden as lower respiratory infections and Tuberculosis are the 4^th^ and 10^th^ leading causes of death world-wide [5]. Finally, two percent of US adults (6.5 million) are hospitalized each year for treatment of community acquired pneumonias caused in part by the near-range airborne transmission of common bacteria and viruses including Influenza [115].^31^

### Short Range (5 m to 50 m)

Airborne disease spread is also known to occur over short distances (5 m to 50 m) [34]. The spread of human pathogens within buildings has been particularly well documented in hospital environments [97], [116]–[118]. Furthermore, outdoor airborne particles are known to infiltrate indoors and cause disease in building occupants.^32^ Building air filtration systems and other measures are in active use to reduce the incidence of airborne animal diseases, e.g., Newcastle Disease virus, and porcine reproductive and respiratory syndrome (PRRS virus) [120]–[123]. Research also indicates that air filtration is beneficial to control MRSA in veterinary settings [124]. Furthermore, high-risk patient areas in hospitals are designed with physical and ventilation barriers to minimize infections in immune compromised patients. Hospital design features include, but are not limited to, permanently sealed hospital room windows and HEPA air filtration [116], [125], [126]. Ultraviolet germicidal irradiation is also routinely used to reduce airborne disease risk in hospital and other facilities, especially for Tuberculosis control [127].

The Newcastle Disease (ND) virus, an avian paramyxovirus, is one well-known example of an airborne disease that can transmit over short distances. ND virus is a commercially important pathogen in poultry production and virulent strains can be devastating, with flock mortality approaching 100% [128], [129]. ND virus has multiple routes of transmission, including airborne. ND vaccines have been a mainstay of ND control, but a number of pandemics have occurred and the disease remains endemic in many countries [129]. With regards to the plausibility of airborne transmission, we note some ND live virus vaccines are delivered via fine aerosolized powders [130]. Furthermore, experimental field studies have documented short- range (60 m) airborne dispersion based on positive viral cultures of air samples [131], [132]. Recent experimental work has reconfirmed an airborne transmission route for ND virus [129]. Furthermore, outfitting buildings with negative air ionization and dilute viricidal chlorine aerosols may be useful in attenuating airborne disease transmission [121], [122].

In plant biology and biosafety studies, short-range (<50 m) airborne particle and pathogen transmissions are thought to be the most frequent scenarios. For example, initial median windborne (anemochorous) plant seed dispersals are typically short (<10 m) but the 95%th percentile for airborne seed dispersion occurs over longer distances and varies significantly by species [133]–[135]. In plant pathology studies of Wheat Stripe Rust (*Puccinia striiformis f. sp. Tritici*) and the wind-dispersed banana plant fungus *Mycosphaerella fijiensis*) [91], [136], single- field experimental studies are used to model initial local plot/field-level airborne pathogen dispersal and clearly demonstrate short-range airborne infection transmissions. These studies have been also used to develop source (emission) estimates for larger scale long-distance disease spread and propagated epidemics.

### Medium Range (50 m to 500 m)

Medium range airborne dispersions of infectious pathogens are well-documented in the plant biosafety, veterinary and human disease Epidemiology literature. These papers include both observational and experimental field studies coupled with physics-based airborne transport and dispersion modeling. Human data for downwind infections are primarily from epidemiology studies of unexpected disease outbreaks, supported by toxicology, clinical studies and environmental studies using background disease rate data.

Epidemiologic disease outbreak studies provide human data for medium range (≤ 500 m) airborne transmission of disease. Well-documented examples include ongoing community-level outbreaks of Legionnaire’s Disease (Legionella *pneumophilia*) from building cooling towers [137]–[145]; Q Fever (Coxiella *burnetii*) transmission from livestock farms to their surrounding communities [37]; as well as Histoplasmosis (Histoplasma *capsulatum*) and Aspergillosis *fumigatus* and *flavus* dispersions from construction work or sites where contaminated soil is disturbed [25]–[29], [146].

On this spatial scale, best-practices and regulatory standards aim to reduce the risk of airborne transport of infectious particles. Guidelines exist to control occupational and environmental construction-associated dust during building renovations. In endemic regions these guidelines are codified into law to reduce infections [147], [148]. Furthermore, there is a long standing, yet still evolving, literature that supports existing regulatory standards aimed at protecting workers and nearby communities from airborne pathogen dispersal from environmental sites such as composting facilities, sewage processing and waste-water aerosols, agricultural gray water aerosols, livestock feed yards and land applications of manure [17], [149]–[155]. As one example, a protective ring of up to 250 m is commonly specified under the assumption that existing air monitors detect little to no airborne material beyond that point [152], [153].

### Long Range (500 m to 500 km)

Long range airborne infectious pathogen transmission is also well-documented in plant biology, the plant and veterinary biosafety literature, as well as in human disease outbreak epidemiology studies. Long-distance atmospheric transmission mechanisms, termed LDD in this literature, have been shown to play a crucial ecological role in plant species invasion, migration and survival as well as plant pathogen dispersal [54], [57], [156]–[159]. This field is well advanced in its understanding of the connection between airborne pathogen transport and dispersion and disease epidemics [58], [160]–[163].

Biosafety experimental field studies also clearly demonstrate kilometer-range dispersion of plant pathogens. For example, fungal plant pathogens are currently an increasing threat to world food security [164]. A wind-dispersed banana plant fungus (*Mycosphaerella fijiensis*) field experiment documented 1 km airborne dispersal in one generation [91]. Studies such as these and others, e.g., [90], together with the above cited plant biology and biosafety literature, demonstrate that the infection probability first decreases quickly with distance which is followed by regimen of kilometer-range LDD events (termed a long “dispersion tail”).

In the United States, long-distance airborne spread of economically significant plant disease across the landscape is an ongoing concern. Predictable seasonal airborne pathogen incursion pathways across the continent are well-identified and routinely monitored to protect crop yields. These continental-scale incursions typically proceed in a stepwise series of shorter (long- distance) airborne dispersions. Chief examples are the seasonal airborne south to north US dispersion incursion pathways across the Midwest Great Plains for wheat stem rust (*Puccinia graminis f. sp. tritici*); the pandemic spread of tobacco blue mold spores (*Peronospora tabacina*) across the Eastern US; and the seasonal US airborne invasion of soybean rust (*Phakopsora pachyrhizi Sydow*) [89], [93], [159], [165], [166].

In the Veterinary literature there are many examples of probable kilometer-range airborne infection transmission. For example, Newcastle Disease virus, Equine Influenza (A/H3N8), Highly Pathogenic Avian Influenza A(H7N7) are important ongoing diseases and each has evidence for long-range airborne virus transmission [132], [167]–[170]. The best described long distance airborne transmitted disease in animals is Foot and Mouth Disease virus, an economically significant disease of veterinary livestock. Long-distance FMDV aerosols are suspected to have contributed to a number of costly, regional-scale disease outbreaks in Europe, including airborne transmission from continental Europe to the United Kingdom [171], [172]. This disease has motivated the significant development and testing of scientific models for long distance infectious aerosol dispersions with the aim of limiting epidemic spread [58], [160]–[162], [173]– [176].

In the human epidemiology literature, many well documented examples exist for airborne disease transmissions over distances greater than 1 kilometer downwind. Coxiella *burnetii*, an endemic disease of ruminants and livestock, is also the cause of Q Fever in humans [59]. Long- distance airborne transmission disease outbreaks from animal farms to human populations have occurred in many European countries [42], [83], [177]–[179]. Notably the recent regional- scale Q Fever epidemic in 2007-2010 in the Netherlands was caused by infectious aerosols emitted from small animal farms [40], [41], [180]–[182]. The epidemic resulted in 4,000 clinical cases and 2,700 hospitalizations [180]. A more recent 2018 follow up of this outbreak showed that among the 519 chronic Q Fever cases identified, 86 patients had died [69]. Legionella pneumophilia dispersions from building cooling towers are also an ongoing source of kilometer- range community Legionnaire’s Disease outbreaks despite the introduction of preventive legal regulations for cooling equipment maintenance [142], [183]. Significant airborne Legionnaire’s disease outbreaks have been reported in many countries including the US, France, Norway, Sweden, and Spain [43], [44], [137]–[141], [143]–[145], [184], [185].

The fungal pathogens Histoplasma *capsulatum* and Coccidioides *immitis* and *posadasii* cause significant human disease due to inhalation (Histoplasmosis and Coccidioidomycosis [or Valley Fever], respectively). Both are endemic in the US: Histoplasmosis in the Eastern and Midwestern states, Coccidioidomycosis in the American West and Southwest [29], [186]. Based on observational epidemiologic studies, three city-wide airborne outbreaks of Histoplasmosis have occurred, two at a community level [187]–[189] and the third being a series of 3 large outbreaks that occurred in urban Indianapolis, IN [190]–[192].

Coccidioidomycosis occurs after inhalation of fungal spores which are widely distributed in southwestern US soils [193], [194]. Forty percent of exposed persons will have clinical symptoms, ranging from an influenza-like illness to disseminated disease and chronic meningitis. Symptomatic disseminated disease requires aggressive treatment and has increased rates of hospitalization and mortality [195], [196]. Desert dust cloud dispersions containing Coccidioides spores are an important ongoing cause of disease in endemic areas [197]. In addition long-distance airborne dust cloud Coccidioides dispersal events triggered by natural disasters have caused significant regional Coccidioidomycosis outbreaks in the US state of California [198]. Kilometer-scale airborne transmission occurred in the Los Angeles area following the 1994 Northridge earthquake, where strong aftershocks generated landslides on the slopes of the Santa Susana Mountains creating large, contaminated dust clouds [45], [46], [199]. These clouds were blown by ambient winds into the urban Simi Valley and Ventura County areas – causing a Coccidioidomycosis outbreak (203 total cases; 55 hospitalizations; 3 fatalities).

### Continental and Global Range (> 500 km)

Continental-scale airborne dispersion events, especially plant seed dispersions, have been well studied and influence the spread of invasive species, metapopulation dynamics, and plant diversity [134], [156], [157], [200]. Continental-scale transport of common environmental bacteria species, either on normal atmospheric air currents or in association with dust cloud dispersions, has also been well-demonstrated [55], [105], [201], [202]. As one example, bacterial communities from the Saharan desert are known to travel airborne to high European Alpine lakes [203], [204]. Pathogenic bacteria have also been observed in the ambient atmosphere, including plant, animal, and human pathogens [205]–[207]. Furthermore, airborne transmission of Neisseria *meningitidis*, a major cause of Meningitis world-wide, is currently under investigation in the endemic Sahel region of North Africa as outbreaks occur most often in dry months with frequent dust storms [202], [206].

Airborne continental-scale disease spread often proceeds as a series of sequential long range airborne transmission events over the landscape (saltatory transmission). However, individual continental-scale airborne disease transmission events, i.e., a single airborne plume transporting pathogens more than 500 km, are also documented in the literature [54], [89], [208], [209]. Most but not all of the existing examples are from agricultural biosafety studies where these events are termed “single-step” LDD pathogen invasions [54]. These types of events are thought to be rare and often associated with extreme weather events or natural disasters [89], [198]. However, routinely occurring, “single-step” LDD events could be more frequent, although the possibility has not been systematically investigated. For example, a sentinel study LDD of airborne pathogenic plant fungi (Erysiplic *graminis*, f.sp. *hordei* [barley mildew] and Erysiplic *graminis*, f.sp *tritici* [wheat mildew]) demonstrated transmission over a distance of 650 km across the North Sea from Great Britain to Scandinavia [209]. Samples were obtained using disease-free receptor plant populations compared to unexposed control plants and a multi-year series of samples were obtained in the highest expected transmission regions.

A major weather-related “single-step” LDD event was the 2,000 km airborne dispersion of Asian soybean rust (Phakopsora *pachyrhizi*) across the Caribbean from northwestern South America to the US during Hurricane Ivan [89]. This 2004 event marked the invasion of Asian soybean rust into the North American continent. The event was anticipated as the spread of Asian soybean rust from Brazil northward in South America was being monitored and Brazil had lost a significant fraction of its soybean production to this pathogen. Among other measures (and prior to the event itself), predictive atmospheric dispersion modeling for potential transport to the US during tropical cyclone seasons were conducted and the US Department of Agriculture deployed disease forecasting systems and field-tested a detailed response plan for use in the event the soybean rust was identified [89]. Soybean rust was detected infesting soybean fields in Louisiana (as predicted) within two weeks after Hurricane Ivan had passed. Subsequently Asian soybean rust has remained endemic in many southern states, especially in the initial epidemic outbreak area [210].

A clear human-disease example of single-plume continental-scale airborne disease transmission is the 600 km dispersion of Coccidioides *immitis* spores in California which resulted in widespread Coccidioidomycosis outbreaks [211]. In this 1977 event, a 160 km h^-1^ windstorm scoured 15 cm of Coccidioides *immitis* contaminated topsoil from Kern County, located in the southernmost basin of California’s great central San Joaquin Valley, carrying a resulting dust cloud to an altitude of 1,500 meters (Image, reference [212]). The dust was transported northward and dispersed over a 87,000 km^2^ area [191], [208], [211], [213] - burying freeways and shutting down interstate transportation. There were 3 immediate storm-related fatalities and 3 firefighters died in a forest fire spread by the strong winds. Sacramento, a low endemicity area 500 km to the north, experienced a large Coccidioidomycosis outbreak (115 cases and 6 fatalities reported vs. a background incidence of 0 to 6 cases per year). Overall fifteen California counties northward in the dust cloud dispersion area reported a ten-fold increase in Coccidioidomycosis cases and 9 counties reported lesser increases [211]. This 1977 Coccidioides *intimus* dispersion, with a total of more than 379 new cases, serves as a historical benchmark for the potential magnitude of Coccidioidomycosis cases from a significant dust storm [208]. Integrated Coccidioidomycosis case surveillance and dust storm forecasting are currently standard in US endemic areas [214].

## Supplemental Material B: Key Atmospheric Transport and Dispersion Modeling Concepts

Atmospheric physics, as well as atmospheric transport and dispersion models of airborne hazards, are well-established and extensive fields. This supplemental material aims to briefly introduce the reader to key concepts needed for this report. We note that while the theory developed in this study is applicable at a wide range of spatial and temporal scales, the epidemiological datasets that are compared to theoretical predictions relate to exposures that occur near the earth’s surface (in the atmospheric boundary layer), from 0.05 km to a few tens of km downwind of an airborne release, and less than a few hours from the time of initial release. The interested reader is referred to [215]–[222] for further details of atmospheric air flow physics, dispersion, and modeling including those present at other spatial and temporal scales.

### Mean Air Flow in the Atmosphere

The mean (time-averaged) air flows are driven by a spatial gradient in atmospheric pressure. When the horizontal surface pressure gradient between two locations is a few millibar over a hundred kilometers (tenths of mb km^-1^), as occurs near the center of a high-pressure weather system; the atmosphere is relatively calm and horizontal surface mean wind speeds are light, typically less than 2 m s^-1^. Larger surface pressure gradients, several to tens of mb km^-1^, result in moderate surface winds (3 to 7 m s^-1^) which are sufficient to stir leaves and twigs. Finally, larger pressure gradients, which can occur during storms, produce strong winds (> 8 m s^-1^). The spatial and temporal distribution of wind, i.e., the “wind field,” may be quite complex as local topography can steer the direction of local winds. For example, winds can be channeled along river and mountain valleys and within urban streets or be blocked by hills or mountains.

### Atmospheric Turbulence

In addition to the regional (mean) air flow described in the previous paragraph, smaller scale motions (turbulent eddies) are typically present in the atmosphere. These turbulent motions are due to two major causes. *Mechanically generated* turbulent motions are generated by drag resulting from the wind moving over and around physical objects on the earth’s surface. The largest mechanically generated eddy size is proportional to the height above ground and obstacle size. *Buoyancy generated* turbulent motions are generated, for example, when the solar heating of the earth’s surface causes warmer (buoyant) air to rise and be replaced by colder air. Buoyancy forces can also decrease turbulent motions, for example, when the surface is cooler than the air. The size of the largest buoyancy generated eddies depends on the atmospheric conditions and can be up to approximately 4,000 m.

Once generated, atmospheric eddies interact with each other and the earth’s surface. These interactions result in the original eddies breaking into smaller eddies. The original eddy’s (turbulent kinetic) energy is transferred to the smaller eddies. This process continues, generating successively smaller eddies until the millimeter size eddies dissipate into heat. This breakdown process results in a well-characterized spectrum of eddy sizes for a given set of atmospheric and surface conditions. One important consequence is that turbulent motions in the atmosphere are correlated over shorter distances (and times) and become uncorrelated at larger distances (and times). Commonly, turbulent motions are correlated on a few minutes timescale.

For many common atmospheric conditions, atmospheric eddies are at least an order of magnitude smaller (spatially and temporally) than the scales of motion present in the regional (mean) air flow. This separation provides a natural division of air motions that transport (mean winds) and dilute (turbulent eddies) contaminated air [222]. This separation results in a gap in the atmospheric energy spectra associated with timescales on the order of an hour.

### Atmospheric Dispersion Physics

If a small amount of airborne material is added to a mass of air (called an air parcel), its presence will not significantly affect the regional wind, eddies, or other atmospheric properties.^33^ Thus the *transport and dilution of these contaminated air parcels can be determined from atmospheric (meteorological) and surface considerations alone and does not depend on properties of the contaminant*. Furthermore, the *airborne exposure to such dilute materials can be mathematically represented as the sum of the exposure to many simpler releases* at specific locations and short time periods (superposition principle). Consequently, insights (and model results) derived for the transport and dilution of single particles released from a single location and time directly contribute to the characterization of more complex cases.

During transport by ambient winds, dispersion occurs when the turbulent eddies, described in the prior subsection, mix surrounding air with the contaminated air parcel which reduces the contamination concentration in the original air parcel and increases the contamination in neighboring air parcels. **Figure B1** shows photographs of this process using point-source smoke aerosol released at a constant rate into a constant airflow. The upper two panels are instantaneous snapshots which clearly demonstrate high moment-to-moment (stochastic) variations in plume concentrations that result from the effect of turbulent eddies. The bottom panel shows a time-lapse photograph of a smoke plume taken over several minutes. This panel shows two important turbulent dispersion regimes: First, near the release, the time-average plume crosswind spread (“width”) depends linearly on distance (and time) from the release because the turbulent motions dispersing the plume are correlated with each other. Second, and farther downwind, the plume spread becomes proportional to the square root of distance (time) from the release because the turbulent motions that disperse the contamination are no longer correlated with the motions present at the time of pollutant release to the atmosphere.

The plume crosswind spread increases with increasing averaging time since larger turbulent motions affect the plume spread at larger averaging times, see **Figure B2**. In some cases, the growth rate of the plume width can dramatically slow after the plume outgrows the size of typical mechanically or buoyancy generated eddies. However, other processes can result in the plume width continuing to grow, for example, due to differential advection – where different parts of the plume are blown in different directions and at different speeds by variations in the mean wind. Differential advection can occur under several conditions including, but not limited to, (a) variation of the mean wind with height, (b) regional scale, horizontal, mean wind variability, and (b) variation of the mean wind with time, e.g., diurnal wind cycles. Differential advection effects are not shown in **Figures B1** and **B2**. *We note that in the absence of additional loss mechanisms (such as radioactive decay or deposition to the earth’s surface), an airborne plume will continue to transport (and dilute) far downwind – indeed plumes of airborne material have been observed on global scales*.

**Figure B1.**
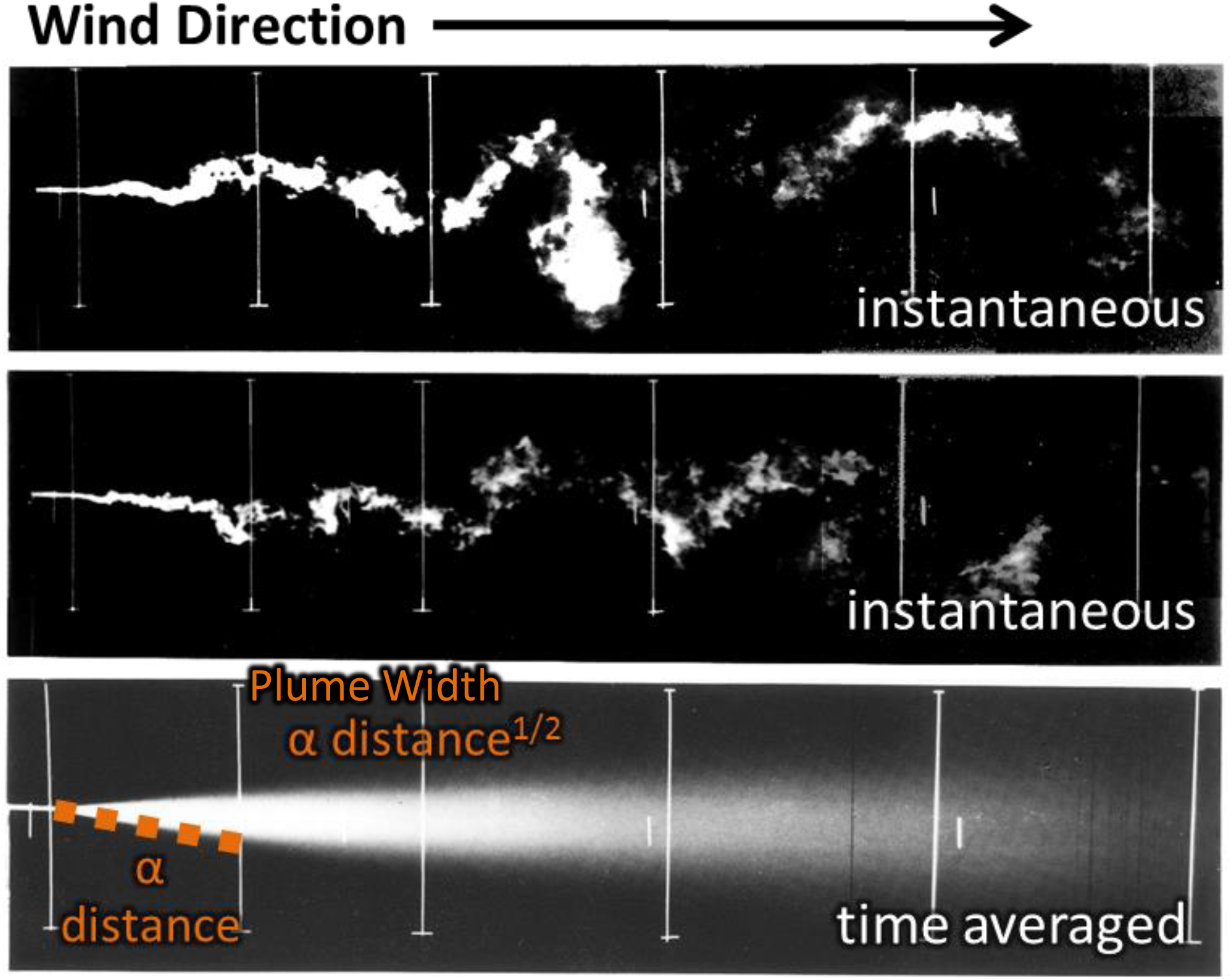
Laboratory photographs of plume dispersion (air flows from left to right). The top two frames are show a snapshot of an individual plume at two different times which highlight the effects of the individual eddies. The bottom frame shows a time averaged concentration over several minutes and more clearly highlight the changes in the growth rate of the plume width with time and downwind distance. (Adapted from image provided by Snyder, EPA)

**Figure B2.**
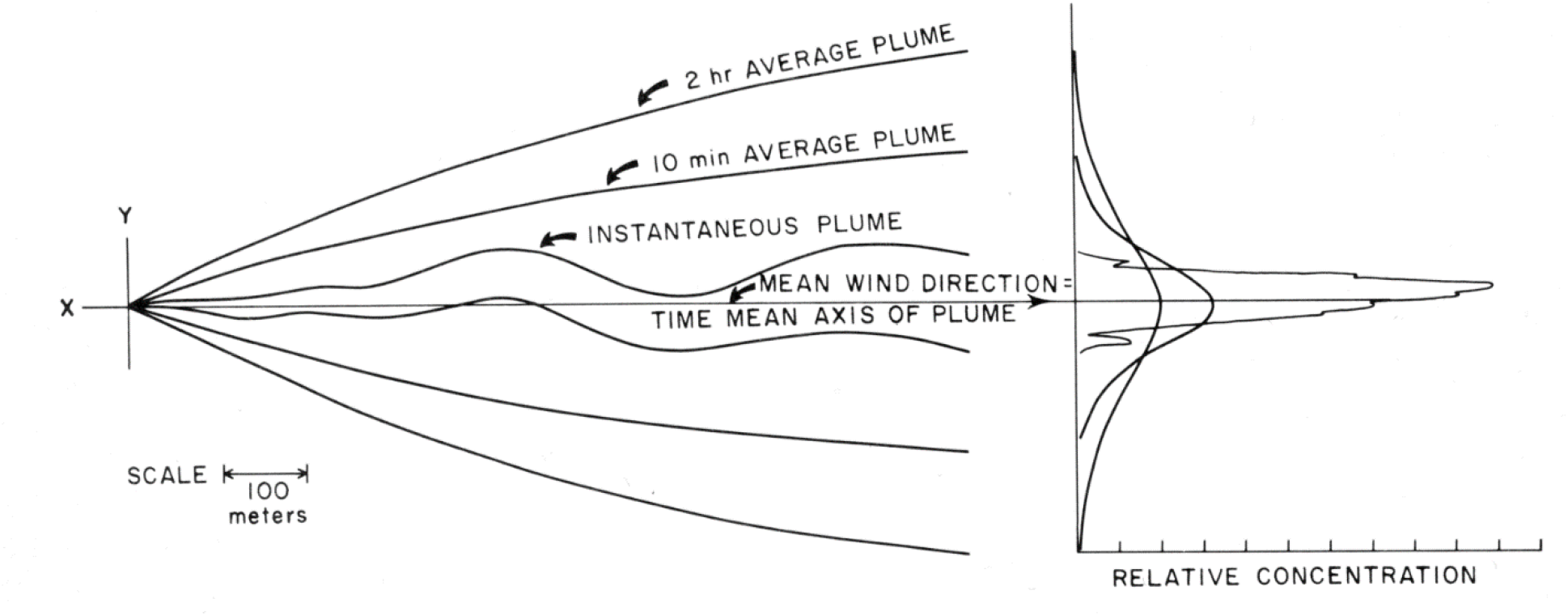
Illustration of the increase in plume width with averaging time taken from Slade [223]. The left panel shows the nominal plume outline for one of three exemplar averaging time (wind blows along the x-axis). The right panel shows the crosswind (y axis) relative plume concentration at a single, nominal downwind point for the three averaging times considered.

### Atmospheric Dispersion Models

Atmospheric transport and dispersion models simulate the previously discussed physical processes of mean wind transport and turbulent diffusion. Direct Numerical Simulation modeling, which explicitly considers all physical processes, can simulate all relevant air motions and contaminant concentrations, e.g., [224], [225]. However, due to their high computational burden and extensive, and often unavailable, input data requirements, these models are not practical outside a research environment. As such, *all practical dispersion models resolve only a portion of the atmospheric motions and physical processes while the effects of the unresolved processes are parameterized, often using statistical descriptions of turbulent motions and fits to empirical data*. As a consequence, these model results are inherently statistical in nature and, depending on the model type, predict (a) the ensemble-average (expected value) result (vast majority of models), (b) the distribution of results (concentration fluctuation models), or (c) a single realization from a distribution of possible flows (Computational Fluid Dynamics models using Large Eddy Simulation). Due to their ubiquity and relevance to the theory developed in this paper, we focus here on the ensemble average models. By analogy, the actual plume at any given time is one of the top two panels in **Figure B1** while the ensemble average dispersion model is predicting the time-averaged bottom panel (in steady state conditions).

There are several common types of ensemble average models, including the classic Gaussian plume model, in which downwind concentrations are calculated analytically; Gaussian puff models, in which one or more expanding “puffs” of contaminant are transported downwind; and Lagrangian particle models, in which downwind concentrations are inferred from the Monte Carlo simulations of the trajectories of computational marker particles.^34^ These model types differ in their ability to resolve key spatial and temporal features as well as to describe complex scenarios. The more detailed model types typically have enhanced computational and input data requirements relative to the simpler models. All of these models will produce nearly identical results for basic scenarios and simpler flows (e.g., constant wind speed and direction) in which in the simpler models are valid (and have been validated against experimental data). Gaussian plume models are analytic solutions to transport and diffusion equations for simple conditions are often used as part of the verification process for the more complex numerical models. *We note that the scenarios used in this study are closely related to a set of classic, straightforward scenarios and so the modeling results presented in this report are expected to be very similar to those that would be produced by different models*.

Atmospheric transport and dispersion models are generally considered to be accurate when they can reproduce individual measurements paired in time and space to within a factor of ten (or better) for a variety of different terrains, downwind distances, and environmental conditions [226], [227]. For example, one modeling system (which is used in this study) is the LLNL ADAPT/LODI diagnostic wind field and Lagrangian particle models [228], [229]. The LLNL ADAPT/LODI models have been validated against a wide range of source terms, environmental conditions, and downwind distances (0.05 to 1,500 km) [230]–[233]. About 50% of the model- measurement comparisons are (a) within a factor of 2 to 5 for simpler releases (source terms) and environmental conditions and (b) within a factor of 10 for more complex conditions. Most of these validation studies use gas tracers; however one experiment used particulate matter.

*When model and measurement data are compared in a way that reduces the importance of stochastic variability, model accuracy can markedly improve*. One case is the Quartile-Quartile (Q-Q) plot which is used, in part, to validate operational transport and dispersion models, e.g., [230], [234]. This plot compares *distributions* of measured and modeled concentration data and does not have the requirement that the model/measurement data is paired in time or space. As one example, when comparing model predictions and measurements from a single tracer study that took place in Copenhagen, 38% of the point to point comparisons are within a factor of 2 but the Q-Q plot demonstrates much better agreement, see **Figure B3** [230]. Another useful metric is integration within respect to time and/or space. For the Copenhagen study, all the model predictions agreed within a factor of 2 of the observed concentrations when integrated along arcs equidistant from the release site (crosswind integrated air concentrations), see **Figure B4**.

**Figure B3.**
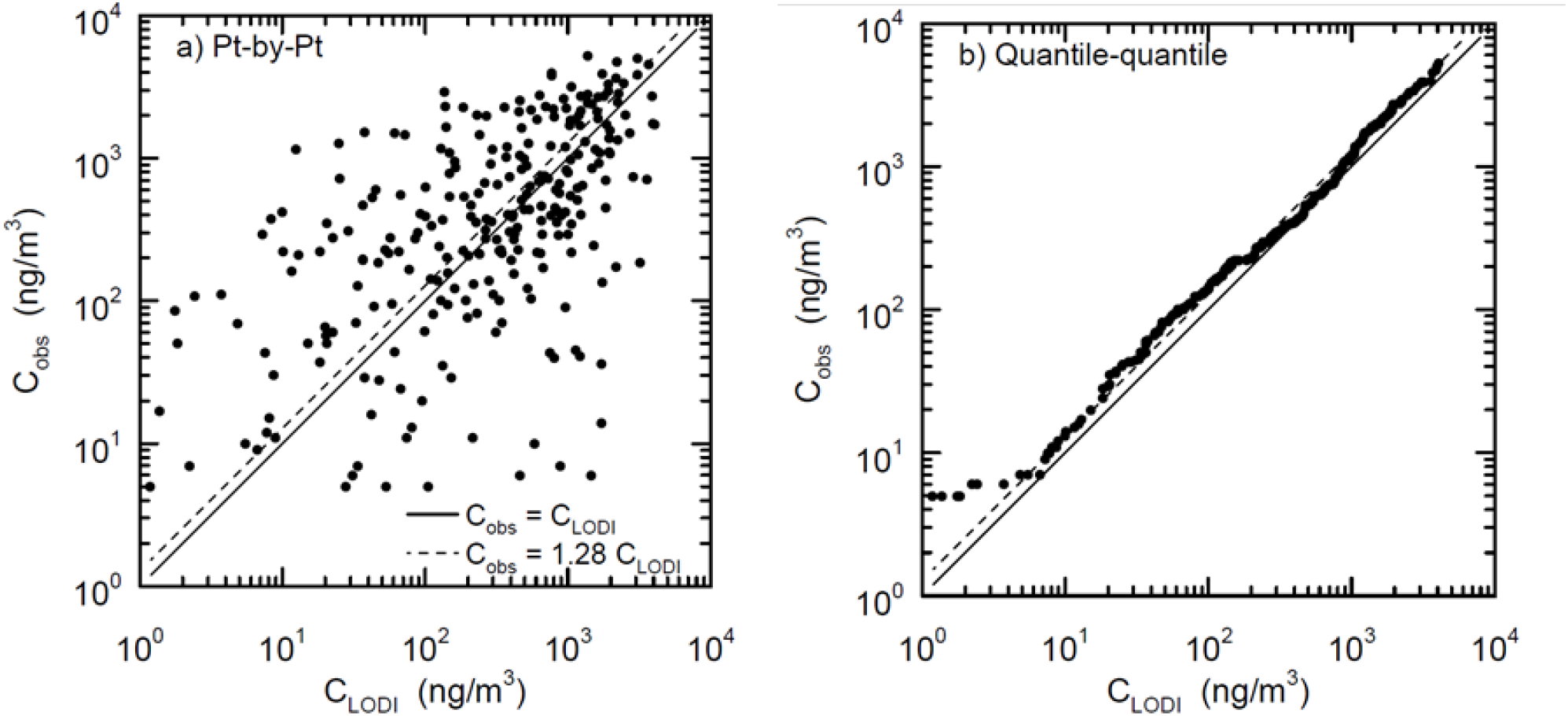
Observed versus predicted ground-level concentrations in the Copenhagen experiment: a) point-by-point comparison, and b) quantile-quantile comparison; only nonzero concentrations included. Dashed line corresponds to geometric mean (0.78) of LODI predicted concentration (C_LODI_) to Observed Concentrations (C_obs_). Image adapted from [230].

**Figure B4.**
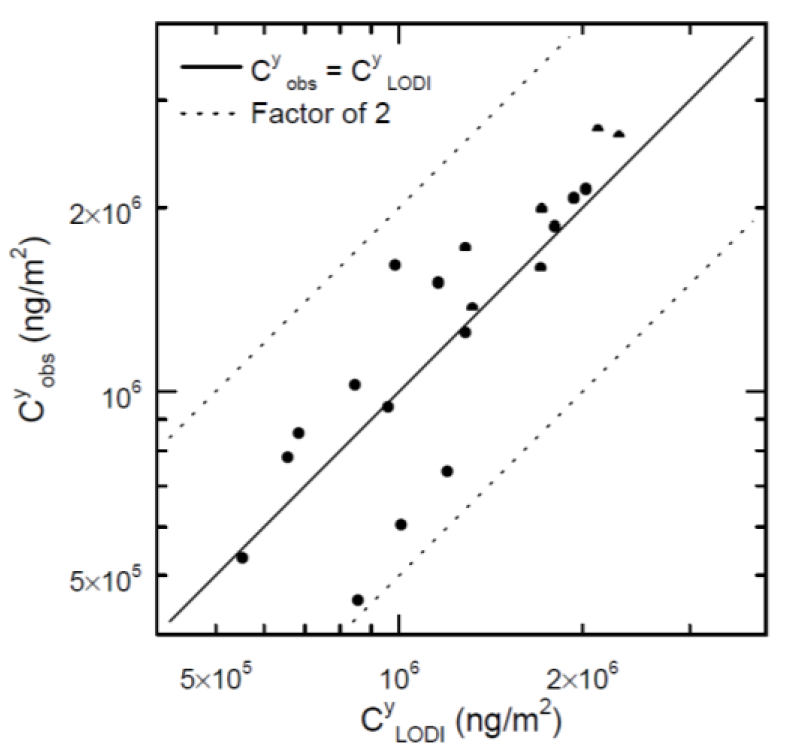
Comparison between observed 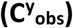 **and predicted** 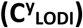 crosswind-integrated concentration at the surface for all Copenhagen experiments except day 7; dotted lines correspond to factor of 2 over- and under- prediction. Image adapted from [230].

## Supplemental Material C: General Theory

In this supplemental material, we derive key theoretical concepts that are implemented in the main paper. Theoretically, the total number of infections can be calculated by individually considering every exposed person. However, this approach can be challenging to implement due to computational considerations, including the need to acquire high resolution input data, and such approaches potentially obscure useful interpretations. Therefore to facilitate the use of **Equations 1, 2**, and **3** presented in the main paper, we derive here a series of self-consistent equations for exposure and infection probability applicable to individuals (indicated by the index *p*), (sub)groups of individuals with the same exposure but varying response to exposure (index *s*), groups of individuals with varying exposures and response to a given exposure (index *g*), and geographic regions containing a group of people (index *r*).

### Section Variables Definitions

*r* = a specific geographic region

*r*_*ref*_ = a geographic region used as a reference in a relative incidence analysis

*r*_*source*_ = geographic region where infectious airborne particles are emitted

*p* = an individual person that could be infected (i.e., a susceptible individual)

*TP* = total number of exposed, susceptible individuals (people)

*s* = a specific (sub)group of individuals with the same exposure but varying response to that exposure

*TS* = total number individuals in (sub)group *s*

*g* = a specific group of individuals with varying exposures and responses to a given exposure

*TSG* = total number of subgroups in group *g*

particle type = specifies particle size and infectivity as a function of time and environmental properties

*b* = a specific particle type

*TB* = total number of particle types

[*Adjustment Factor*](*g, b*) = scaling factor for group *g* that accounts for the deviation of *b*- type particle exposure and response from that of the reference exposure and response (no units)

[*Adjustment Factor*](*r, b*) = scaling factor for region *r* that accounts for the deviation of *b*- type particle exposure and response from that of the reference exposure and response (no units)

[*Area*](*r*) = area of region *r* (m^2^)

[*Effective Source Term*](*r, r*_*source*_, *b*) = mathematical construct used to scale infection probability from one region to another for *b*-type particles. (m^3^ s^-1^ people^-1^)

[*Exposure*](*p*) = number and type of infectious airborne particles in the breathing volume (respiratory second volume)^35^ of individual *p* (particles s m^-3^)

[*Exposure*](*p, b*) = the number of *b*-type infectious airborne particles in the breathing volume (respiratory second volume) of individual *p* (particles s m^-3^)

[*Exposure*](*s, b*) = number of *b*-type infectious airborne particles in each individual’s breathing volume (respiratory second volume) for individuals in (sub)group *s* (particles s m^-3^)

[*Exposure*]_*ref*_(*g, b*) = reference number of *b*-type airborne particles in the breathing volume (respiratory second volume) of an individual in group *g* (particles s m^-3^)

[*Exposure*]_*ref*_(*r, b*) = reference number of *b*-type airborne particles in the breathing volume (respiratory second volume) of an individual in region *r* (particles s m^-3^)

[*Exposure Adjustment Factor*](*g, s, b*) = scaling factor that for group *g*, subgroup *s* accounts for the deviation *b*-type particle exposure from the reference exposure (no units)

[*Exposure Adjustment Factor*](*r, s, b*) = scaling factor that for region *r*, subgroup *s* accounts for the deviation *b*-type particle exposure from the reference exposure (no units)

*Health Effect Model*(*p*, [*Exposure*](*p*)) = mathematical model describing the probability that an individual *p* will be infected given the individual’s specific exposure (no units)

[*Infection Probability*](*p*) = probability that an individual *p* becomes infected (no units)

[*Infection Probability*](*s*) = mean probability that a random individual in (sub)group *s* becomes infected. All individuals in (sub)group *s* have the same exposure. (no units)

[*Infection Probability*](*g*) = mean probability that a random individual in group *g* becomes infected (no units)

[*Infection Probability*](*r*) = mean probability that a random individual in region *r* becomes infected (no units)

[*Infection Probability*](*r, b*) = mean probability that a random individual in region *r* becomes infected by a *b*-type particle (no units)

[*Infections*] = total number of people infected (people)

[*Infections*](*r, b*) = total number of people infected by *b*-type particles in region *r* (people)

[*Infectious People*](*r*_*source*_) = total number of people capable of emitting infectious particles in source region *r*_*source*_ (people)

[*Metric of Interest Probability*](*p*) = probability that an individual *p* exhibits the metric of interest (no units)

[*Normalized TSIAC*](*r, b*) = *b*-type particle air concentration integrated over region *r* and the passage of the airborne infectious plume assuming a single particle was released at the source (s m^-1^)

[*Normalized TSIAC*](*r, r*_*source*_, *b*) = *b*-type particle air concentration integrated over region *r* and the passage of the airborne infectious plume assuming a single particle was released from source region *r*_*source*_ (s m^-1^)

[*Population Density*](*r*) = population density in region *r* (people m^-2^)

[*Population Probability*](*g, s*) = probability that an individual in group *g* is in subgroup *s* (no units)

[*Refactive Infection Probability*](*r, b*) = ratio of the region *r* infection probability to reference region infection probability due to exposure to *b*-type particles (no units)

[*Refease Probability*](*b*) = probability that a particle released into the environment is a *b*- type particle (dimensionless)

[*Response Adjustment Factor*](*g, s, b*) = scaling factor that for group *g*, subgroup *s* accounts for the deviation of *b*-type particle response from the reference response (no units)

[*Single Particle Infection Probability*](*p, b*) = the probability that individual *p* will become infected after being exposed to a single *b*-type particle. This term includes the probability that particle(s) will be inhaled and deposit in the respiratory system. (m^3^ s^-1^ particle^-1^)

[*Single Particle Infection Probability*](*s, b*) = the mean probability that a random individual in (sub)group *s* will become infected after being exposed to a single *b*-type particle. This term includes the probability that particle(s) will be inhaled and deposit in the respiratory system. (m^3^ s^-1^ particle^-1^)

[*Single Particle Infection Probability*]_*ref*_(*g, b*) = reference probability that an individual in group *g* will become infected after being exposed to a single *b*-type particle. This term includes the probability that particle(s) will be inhaled and deposit in the respiratory system. (m^3^ s^-1^ particle^-1^)

[*Single Particle Infection Probability*]_*ref*_(*r, b*) = reference probability that an individual in region *r* will become infected after being exposed to a single *b*-type particle. This term includes the probability that particle(s) will be inhaled and deposit in the respiratory system. (m^3^ s^-1^ particle^-1^)

[*Single Particle Metric of Interest Probability*](*p, b*) = the probability that individual *p* will exhibit the metric of interest after being exposed to a single *b*-type particle. This term includes the probability that particle(s) will be inhaled and deposit in the respiratory system. (m^3^ s^-1^ particle^-1^)

[*Source Adjustment Factor*](*r*_*source*_, *b*) = scaling factor that accounts for the deviation of *b*- type particles emitted from the *r*_*source*_ region from that of a reference source region (no units)

[*Subgroup Adjustment Factor*](*g, s, b*) = scaling factor that accounts for the deviation of group *g*, subgroup *s, b*-type particle exposure and response from that of the reference exposure and response (no units)

[*Total Particles Released*] = total number of particles released into the atmosphere (particles)

### Conceptual Model

The general environmentally-mediated infection process can be mathematically represented by **Equations C1 and C2**.

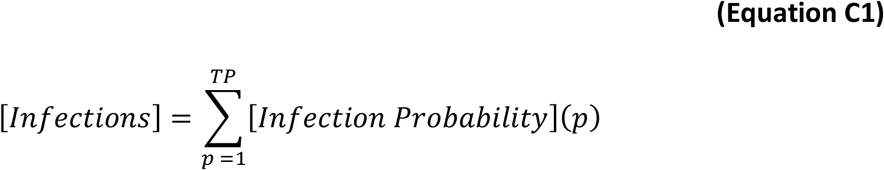

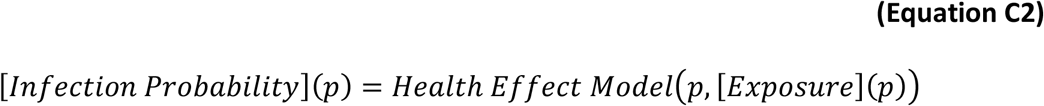

### Rare Exposures

If there are *TB* different types of particles AND any individual inhales either one or no particles, then the probability of an individual *p* becoming infected is shown in **Equation C3**.^36^

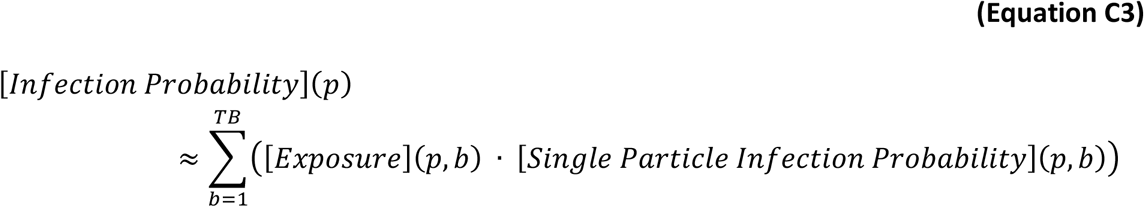

### Subgroups

Consider a (sub)group *s* comprised of *TS* individuals with same exposure, but varying response to that exposure,^37^ then **Equations C4** and **C5** provides the mean infection probability for (sub)group *s*.

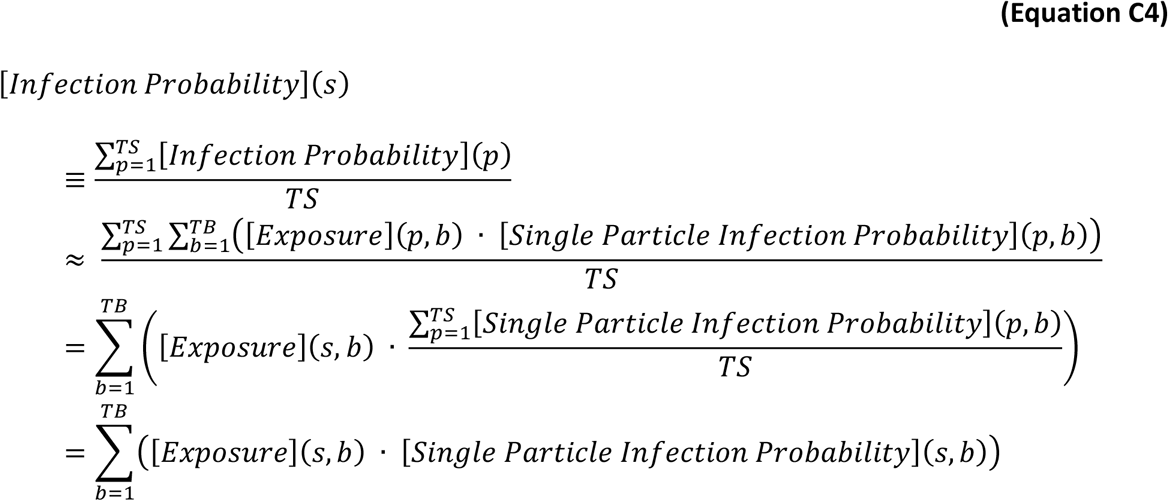

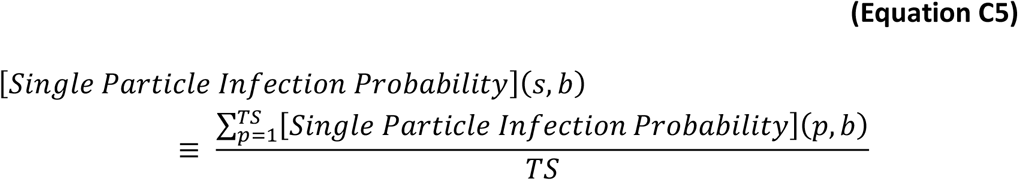

### Groups

When exposures within a group of people are not constant,^38^ each subgroup can be considered a separate, constant exposure group and the overall group’s (mean) infection probability is equal to the population weighted average of the individual subgroup infection probabilities, see **Equation C6**.

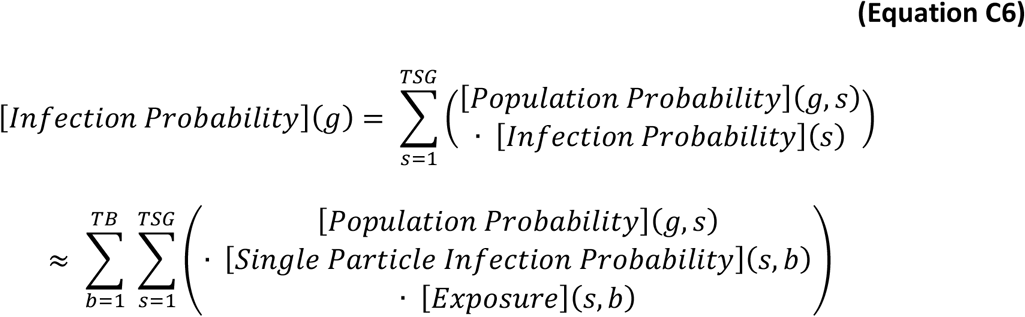

We assume that the individual exposures and health effects can be defined as a ratio to a reference exposure and response, respectively. While these ratios can take any value and vary by individual, the individual-specific value cannot change. With these assumptions, **Equation C6** can be re-written as **Equation C7** to **C11**.

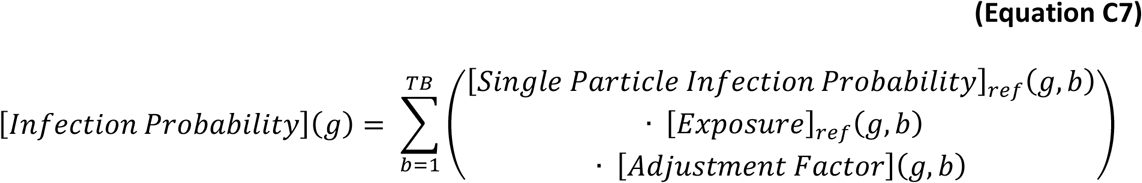

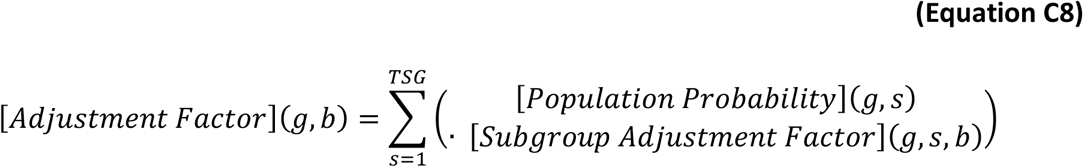

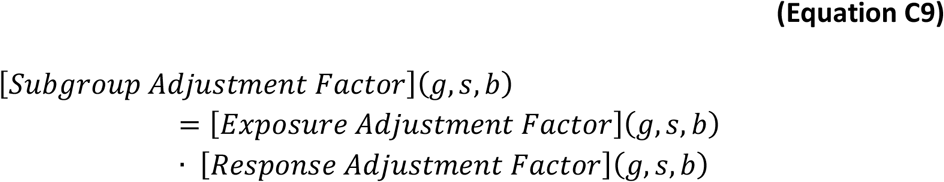

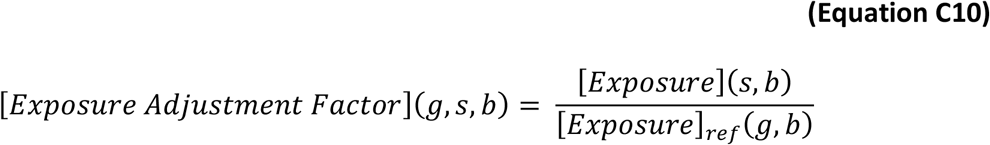

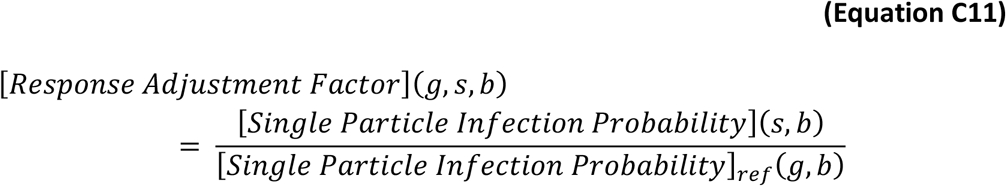

### Absolute Infection Probability for Geographic Regions

Geographic regions, e.g., zip codes and census tracts, are often used when reporting epidemiological data and defining outbreak response zones, e.g., quarantine and/or vaccination. For this case, we define region *r* as a type of group and **Equation C7** can be rewritten as **Equation C12** where the reference exposure is defined by **Equation C13**.

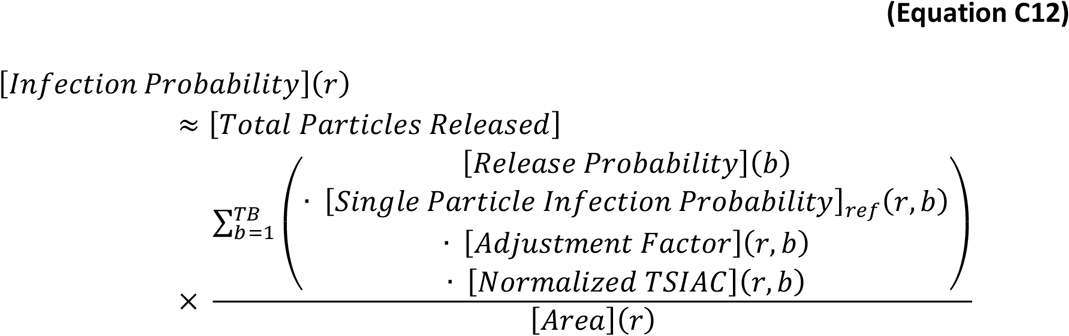

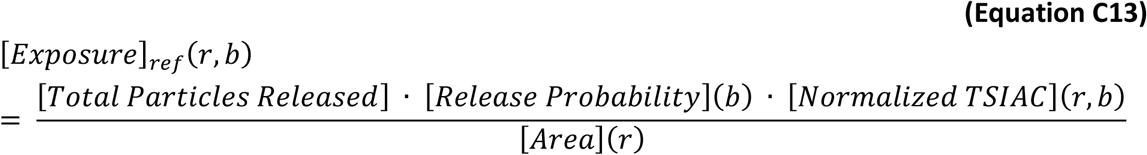

When considering outdoor plumes of infectious particles, we adapt the Regional Shelter Analysis (RSA) methodology to assign each location (subgroup *s*) a “protection factor” which is defined as the ratio of the outdoor to indoor exposures. As shown in [30], [31]; building protection factors depend only on the building operating conditions, environmental parameters, and the particle type. Thus, for a given particle type, the assigned protection factors are identical to the inverse of the [*Exposure Adjustment Factor*](*r, s, b*) where [*Exposure*]_*ref*_(*r, b*) is the outdoor time-integrated air concentration of *b*-type particles during the passage of the airborne infectious plume over region *r*.

### Relative Infection Probability for Geographic Regions

Given a plume of airborne infectious particles from a single source, then **Equation C12** implies **Equation C14** when a single particle type dominates the exposures. **Equation C14** does not depend on the release or the infectivity of individual particles. When the two regions are similar (e.g., both *r* and *r*_*ref*_ are residential areas with similar demographics) the adjustment factor ratio is unity. We demonstrate the utility of **Equation C14** in the *4. Results* and *5. Discussion* sections.

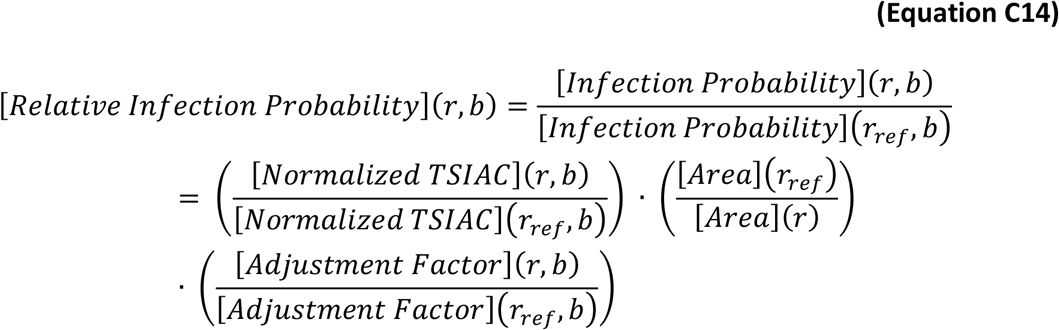

### Extrapolating Regional Infection Probabilities

When a clear case of region-to-region airborne disease transmission has been identified and a single particle type dominates the exposures, **Equation C16**, derived from **Equations C15(a-b)**, estimates an effective source term for each infectious person in the source region.

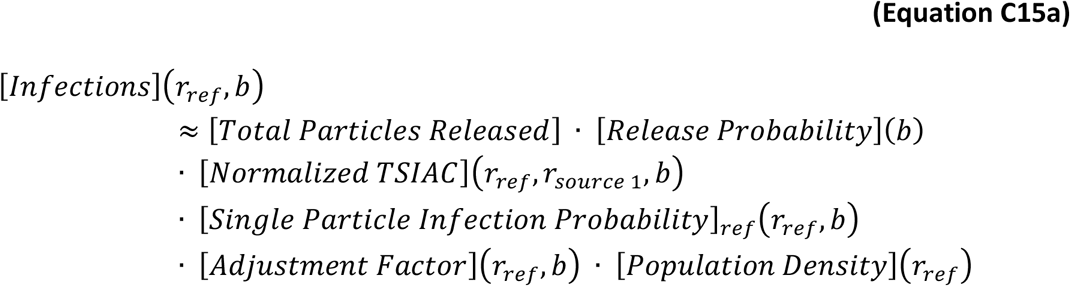

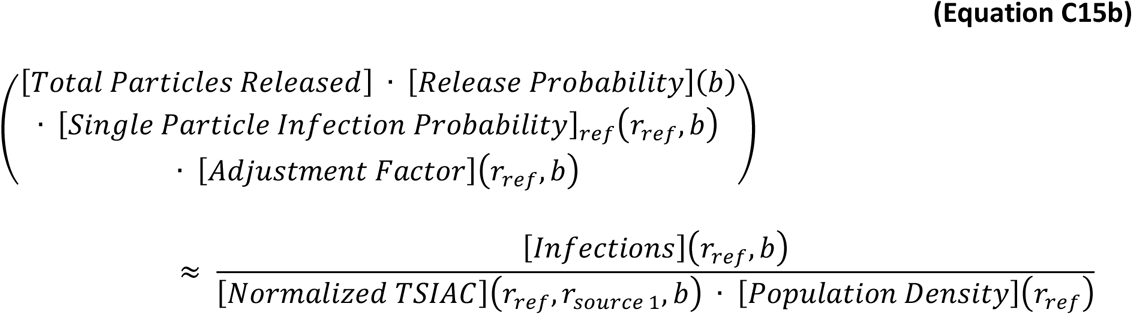

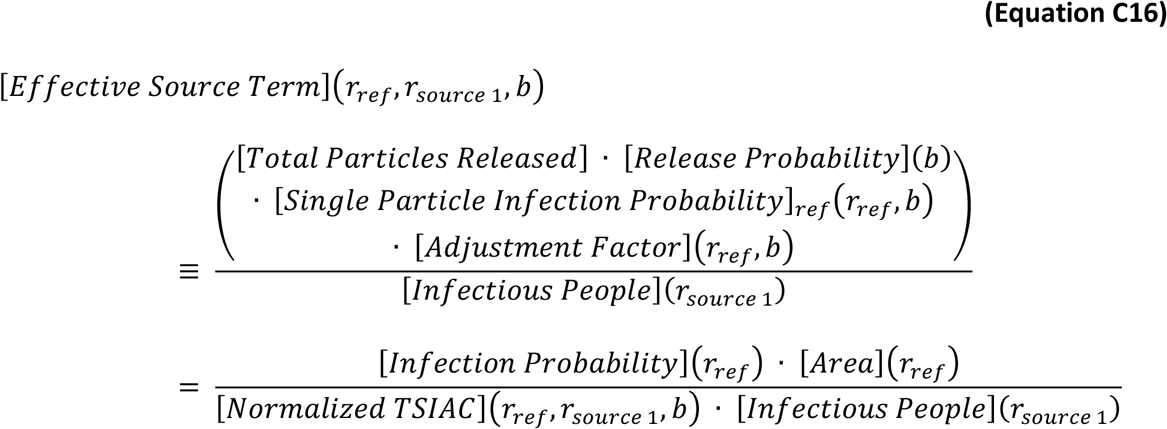

Given the effective source term (**Equation C16**), the infection rates in other regions and times can then be estimated with **Equation C17a**. Alternately, **Equation C17b** can be used without the need to calculate the intermediate “effective source” quantity. The new term, the source adjustment factor, is introduced which adjusts for the number of airborne infectious particles emitted, e.g., a reduction in cough-emitted infectious particles due to use of respiratory masks. As with **Equation C14**, the reference and source regions disease adjustment factor may also be similar, i.e., their ratio may be unity, if buildings and demographics are similar in both source and target regions.

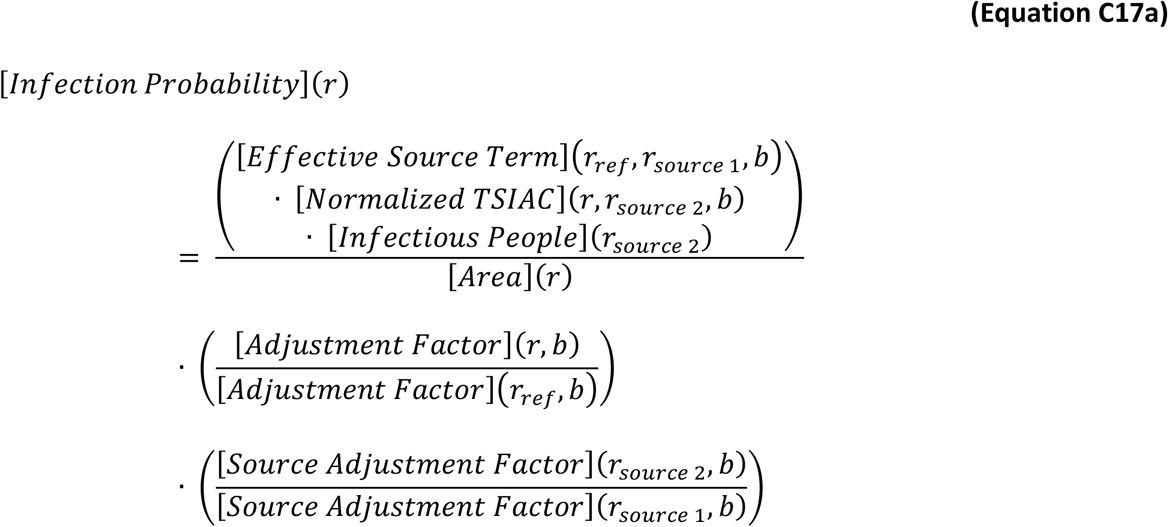

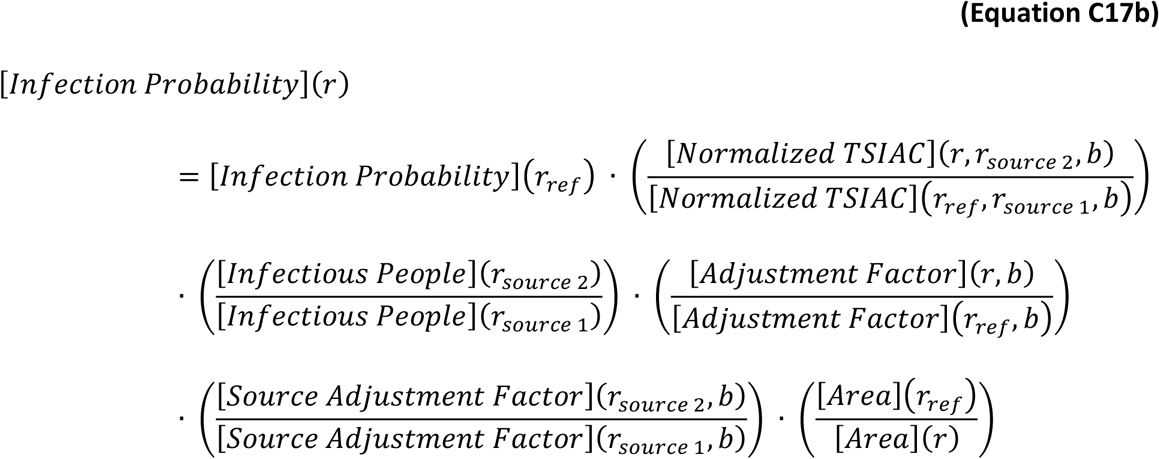

### Relationship to Disease and Other Metrics

The Supplemental Material equations presented up to this point focus on infection probability. However, not all infections result in disease. In addition, other metrics, such as probability of needing medical resources, may also be of interest. When the probability of the metric of interest, e.g., disease, can be linearly related to exposure, see **Equation C18**, then it is straightforward to adapt the prior equations for the metric of interest by replacing [*Infection Probabilty*](*p*) with [*Metric of Interest Probability*](*p*). We note than while [*Single Particle Metric of Interest Probability*](*p, b*) can take any value and vary by individual and/or particle type, the value for a specific individual and particle type cannot change. We note that the adjustment factors can vary by metric.

*The relative incidence of multiple metrics of interest, such as infection and disease are the same, since* ***Equation C14*** *is unchanged by this substitution*.

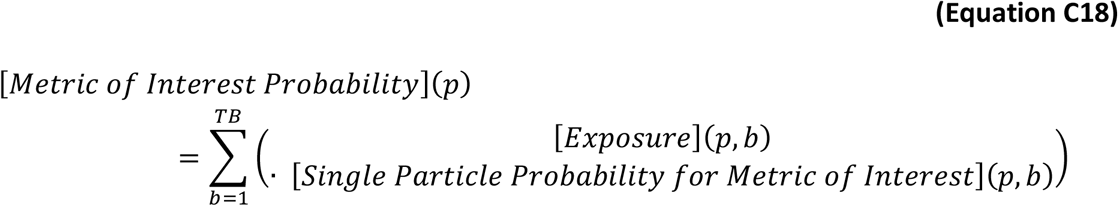

## Supplemental Material D: Indoor Particle Dynamics

This supplemental material derives equations to estimate three related, but distinct building properties: (1) the degree to which indoor individuals are exposed to outdoor-origin airborne particulates, (2) the degree to which indoor individuals are exposed to indoor-origin airborne particulates, and (3) the faction of indoor-origin particulates that exit the building and enter the outdoor atmosphere. The first section, building protection, is adapted (with minor edits) from a prior publication [30] which also provides detailed discussion on the relevant indoor particle dynamics and representative estimates for both (a) parameter values (which are also used in the other sections) and (b) protection estimates for US buildings and Census Tracts. The other two subsections, which extend the building protection material, are new.

While we do not discuss prior approaches in detail in this report, we note the Wells-Riley model which estimated the indoor infection risk to susceptible populations who share the same building as potentially infectious individuals. The Wells-Riley model also provided quantitative guidance on the indoor loss rates and ventilation rates required to reduce disease spread [3], [236]. Later extensions of this model have addressed many of its original limitations. However, all versions retain the concept of a “quantum of infection” whose definition combines many key physical properties, including the probability of infection when there is exposure to infectious particles [237].

### Section Variables Definitions

*λ*_*dep*_(*particle size*) = the particle size dependent indoor deposition loss rate (h^−1^)

*λ*_*decay*_ = the “generic” first-order airborne decay (loss) rate (h^−1^)

*λ*_*in*_ = the rate at which outdoor airborne particles enters the building – typically via infiltration or ventilation. Includes losses that occur during transport from outdoor to indoor (h^−1^)

*λ*_*inf*_ = the air infiltration rate at which air enters a building, i.e., the air change rate (h^−1^)

*λ*_*internal*_ = the rate at which indoor airborne particles are lost within the building – typically by deposition to surfaces or by filtration (h^−1^)

*λ*_*out*_ = the rate at which indoor airborne particles exit the building – typically via exfiltration or ventilation (h^−1^)

*λ*_*T*_ = the total building ventilation rate = sum of the infiltration and mechanical ventilation rates (h^−1^)

[*Building Exit Fraction*] = fraction of indoor airborne material that exits the building and enters the outdoor atmosphere. (no units)

[*Building Floor Area*] = floor area of the building (m^2^)

[*Building Volume*] = volume of the building (m^3^)

[*Normalized TSIAC*]_*indoor*_ = indoor time and space integrated air concentration assuming a unit amount of material is released (s m^-1^)

[*Room Height*] = height of building occupied space (m)

[*Total Particles Released*] = total number of particles released into the indoor air (particles)

*C*_*Indoor*_(*t*) = the indoor particle air concentration at time t (g m^-3^)

*C*_*Outdoor*_(*t*) = the outdoor particle air concentration at time t (g m^-3^)

*L*_*inf*_(*particle size*) = the particle size dependent efficiency by which particles can penetrate the building shell (dimensionless)

*L*_*out*_(*particle size*) = the particle size dependent efficiency by which particles in indoor air exit the building through cracks and other penetrations in the building shell (dimensionless)

*F*_*filter*_(*particle size*) = the particle size dependent filtration efficiency (dimensionless)

*F*_*oa*_ = the fraction of outdoor air passing through the HVAC supply fan (dimensionless)

*F*_*r,fan*_ = the fraction of time the forced air furnace recirculation fan is on, i.e., the fan’s duty cycle (dimensionless)

*r*_*fan*_ = the rate at which a building volume of air recirculates through the furnace systems when the fan is on (h^−1^)

*v*_*fan*_ = the rate at which a ventilation or HVAC supply fan delivers air to the building when the fan is on and combines the outside and recirculation air rates (building volume h^−1^)

*t* = time (h)

*τ* = an integration variable (h)

### Building Protection Against Outdoor-Origin Particles

Indoor individuals can be exposed to particles of outdoor origin when these contaminants enter buildings through mechanical ventilation, e.g., heating, ventilation and air conditioning (HVAC) systems, natural ventilation (e.g., open windows), and/or infiltration (e.g., exterior wall cracks). Particles may also be transported into buildings via deposition on outdoor surfaces or fomites that are subsequently tracked indoors by individuals, or otherwise transported, into the building and then resuspended into the indoor air. These transport pathways are illustrated in the top panel of **Figure D1**. Once indoors, airborne particles can be removed from the indoor air through (a) air leaving the building through mechanical or natural ventilation and exfiltration; active filtration within ventilation systems (if present); (c) deposition on indoor surfaces (some particles may resuspend); and (d) other processes, including radioactive decay, chemical reactions, stand-alone indoor air filtration systems, and the loss of infectivity of airborne infectious agents, among others. The latter three loss terms are illustrated in the bottom panel of **Figure D1**.

Modeling indoor contaminant concentrations requires choosing among a variety of models with increasing complexity, ranging from simple single-compartment models to multizone models to highly detailed computational fluid dynamics models. While increasingly detailed and complex models may reduce modeling conservatism and uncertainty, the number and fidelity of the input parameters also increases (see [238] for a general discussion). Detailed parameters are not generally available for many buildings of interest therefore we make two key modeling assumptions: (a) that indoor volumes can be represented as a single compartment, which can be used to describe the time evolution of indoor contaminant concentrations, and (b) that concentrations within that single compartment are spatially uniform [36], [119], [239], [240].^39^

**Figure D1.**
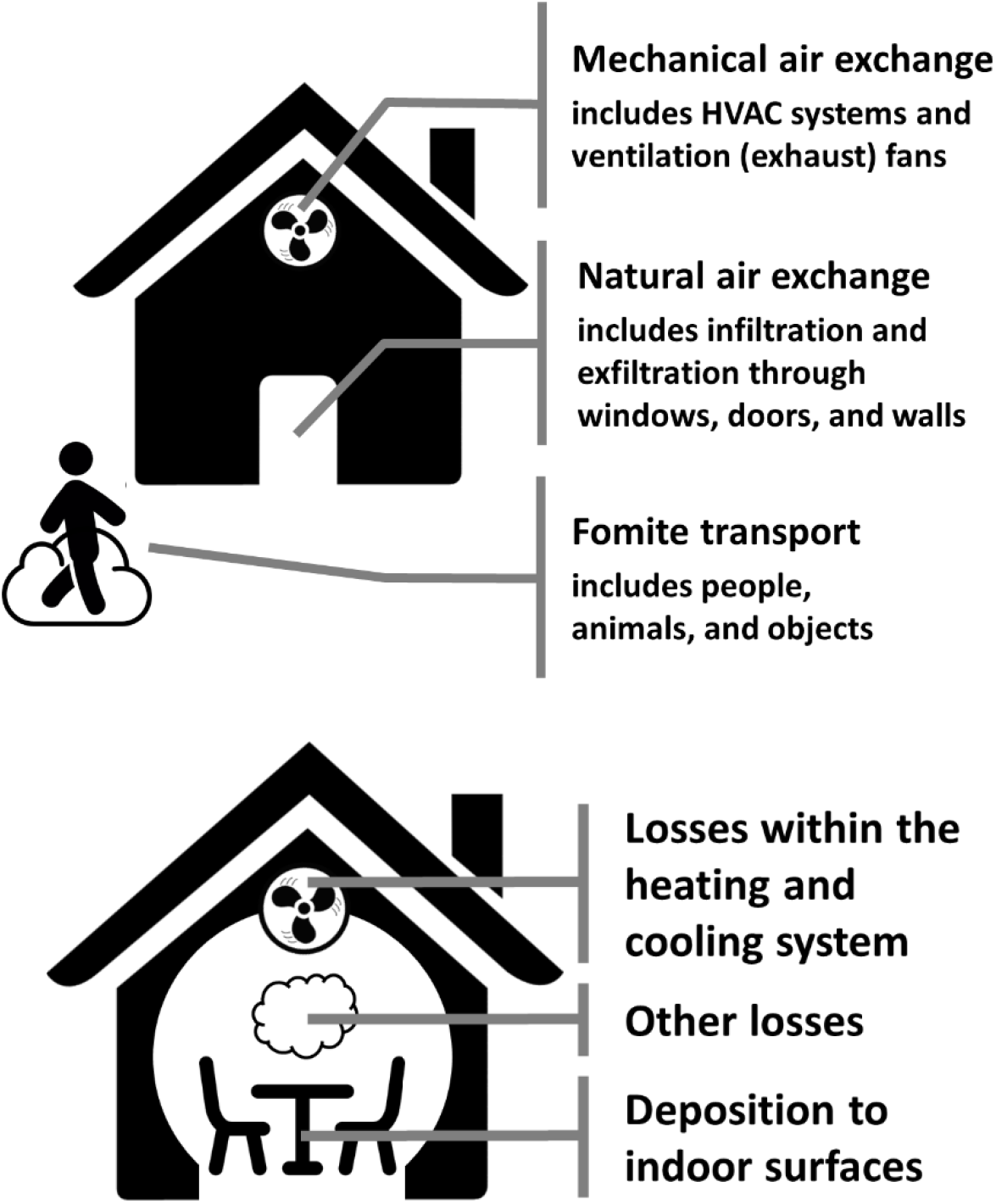
Illustrations of (top) mechanisms by which airborne material and fomites can travel between the outdoor and indoor environments and (bottom) indoor loss mechanisms.

These assumptions are codified in the single box model (**Equation D1a**) which can be used to describe the time evolution of indoor air concentrations after outdoor contaminants have entered a given building. This study includes the additional, commonly used assumption that the transport and loss terms, i.e., the λ parameters, are independent of both time and air concentration on the timescales of interest. **Equation D1a** thus reduces to **Equation D1b**. Derivations provided in the methodology report [31] demonstrate that **Equation D2** provides the building protection factor (the ratio of outdoor to indoor exposure).^40^

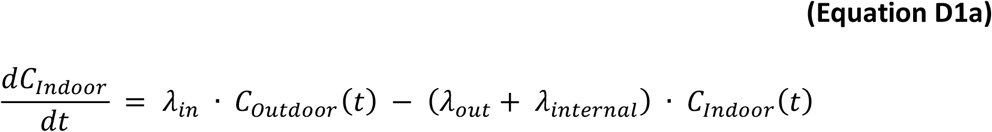

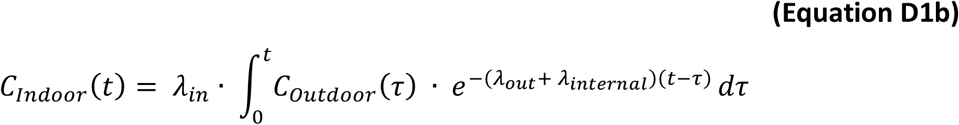

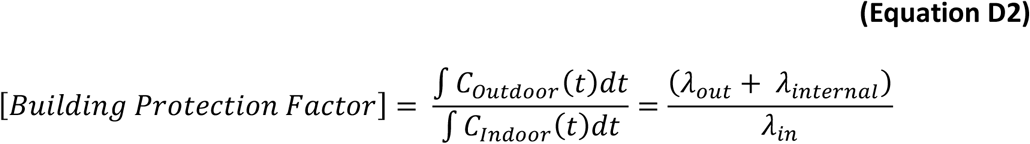

We adapt **Equation D2** to develop two generalized building PF equations to cover the range of building types typically found in the US. The equation terms are grouped to correspond to the *λ*_*in*_, *λ*_*out*_, and *λ*_*internal*_ terms in **Equation D2**. Each PF equation is based on one of two common combinations of (a) the three airflow mechanisms described above and (b) the relevant indoor loss mechanisms. In addition to any filtration that might be present, and already incorporated into the airflow terms, we consider two additional indoor loss mechanisms: (1) deposition of airborne particles to indoor surfaces, *λ*_*dep*_, and (2) a generic first-order airborne loss mechanism, *λ*_*decay*_, which can be used to represent radioactive decay (radiological hazards) or airborne loss of infectivity (biological hazards). Several parameters are particle size dependent. For readability, these dependencies are not shown in the equations themselves, but are noted in the variable list definitions above. See [30] for a detailed discussion of parameters and their common values.

**Equation D3** specifies the protection factor for buildings with filtered recirculation. In the US, these are typically residences. Outdoor airborne material enters the building only through the infiltration pathway. The forced air furnace recirculation air filter, if present, removes indoor airborne particles.

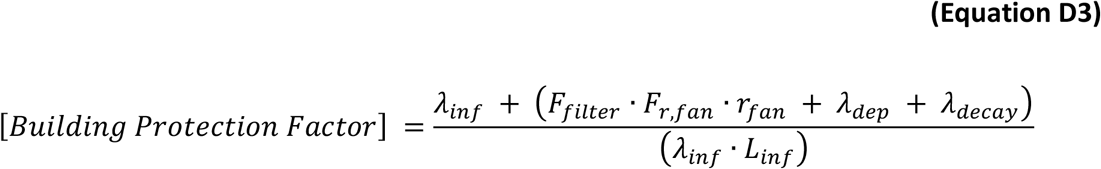

**Equation D4** specifies the protection factor for buildings with an active HVAC system. In the US, these are typically commercial buildings. Outdoor airborne material enters the building through either infiltration or the HVAC system outdoor air intake. The HVAC system air filter removes airborne particles from both the entering and recirculating air. This equation implicitly assumes that the HVAC system is always moving building air although not necessarily heating or cooling it, i.e., the fan duty cycle is 100%.

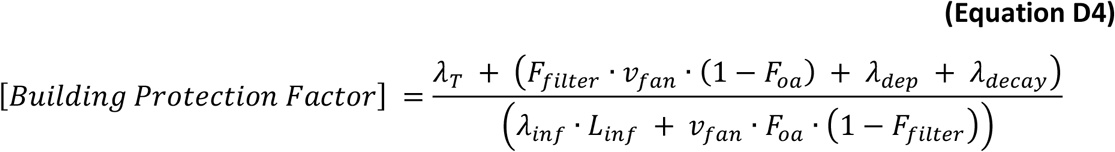

### Normalized Indoor Exposures

With the additional assumptions that outdoor concentrations are zero and all indoor emissions occur at a single time (t = 0), **Equation D1a** simplifies to **Equations D5(a-b)**. When a unit amount of material is released at t = 0, **Equation D5** reduces to **Equation D6**.

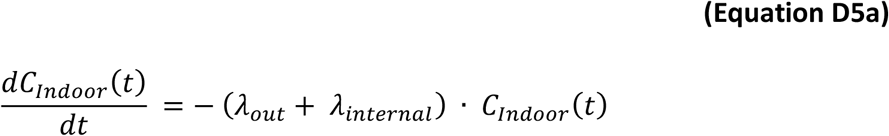

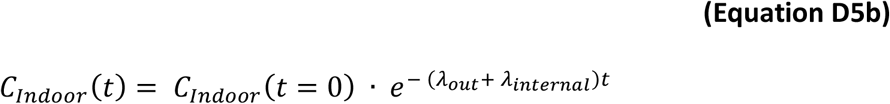

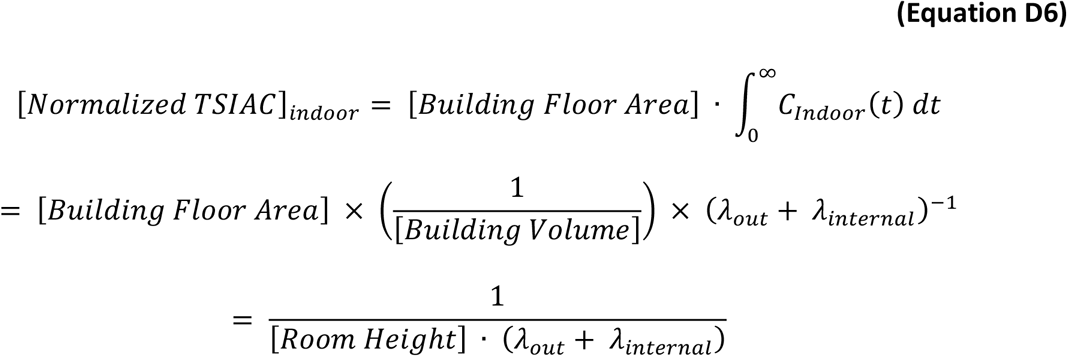

We now adapt the general building protection equations to estimate the [*Normalized TSIAC*]_*indoor*_. The equation terms are grouped to correspond to the *λ*_*out*_ and *λ*_*internal*_ terms in **Equation D6**.

**Equation D7** specifies [*Normalized TSIAC*]_*indoor*_ for buildings with filtered recirculation (US residences).

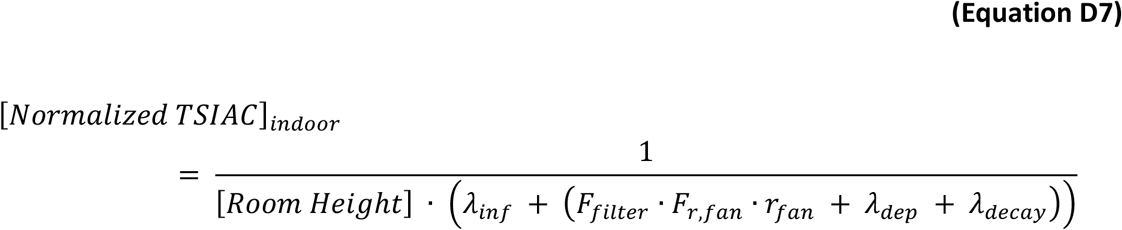

**Equation D8** specifies [*Normalized TSIAC*]_*indoor*_ for buildings with an active HVAC system (US commercial buildings).

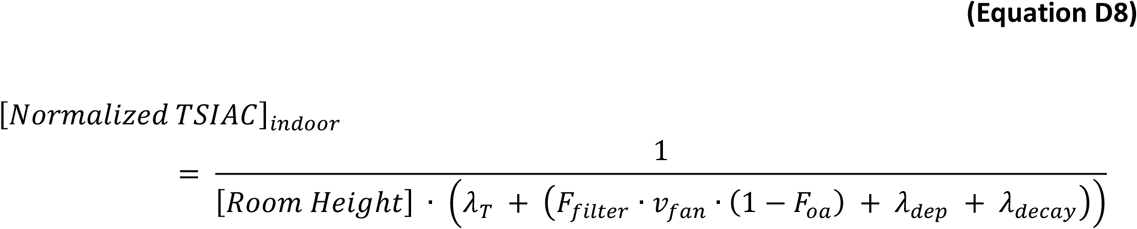

### Fraction of Indoor Airborne Particles Released to the Outdoors

**Equation D9** shows the fraction of indoor airborne material that exits the building and enters the outdoor atmosphere. This equation is based on (a) the (normalized) indoor concentration, (b) the rate at which indoor air (and airborne material) leaves the building, and (c) particulate losses that occur while airborne particles are suspended in indoor air exiting the building.

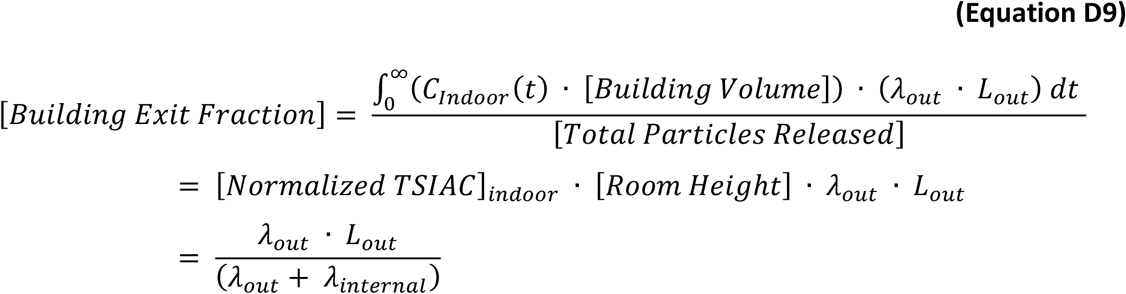

For buildings with filtered recirculation (US residences), outdoor air enters the building only through the infiltration pathway and so *λ*_*out*_ = *λ*_*inf*_ and **Equation D10** shows the fraction of material released indoors that exits the building. The entrance and exit airflow paths may differ. For examples, outdoor air could enter the building through wall cracks while some of the indoor air could exit the building through exhaust fans. Thus, entrance and exit loss fractions, *L*_*in*_ and *L*_*out*_, respectively, can in theory differ. However at a practical level, there is little data on which to estimate the *L*_*out*_ term. In addition, small, e.g., 1 μm aerodynamic diameter, particle losses are minimal. Therefore we assume *L*_*out*_ = *L*_*inf*_ and the [*Exit Fraction*] *is equal to the inverse of the building protection factor as derived in* [30]. For context, 35%, on average, of the airborne, 1 μm aerodynamic diameter, stable (do not lose infectivity in the air) particles released in US single family homes are expected to exit the building (and be released to the outdoor atmosphere) [30].

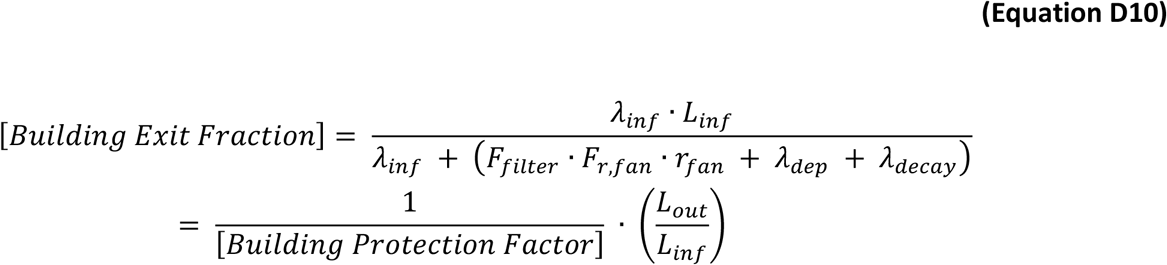

For buildings with an active HVAC system (US commercial buildings), indoor airborne material exits the building through either infiltration or through mechanical means, e.g., the HVAC system exhaust. Again we note that the entrance and exit airflow paths may differ, but as a practical matter assume (a) that they are the same (*λ*_*out*_ = *λ*_*inf*_; *L*_*out*_ = *L*_*inf*_) and (b) no particles are lost while the indoor air is exiting through the exhaust system. ***Equation D11***, *which shows the fraction of material released indoors that exits the building, is not the same as the building protection factor derived in* [30].

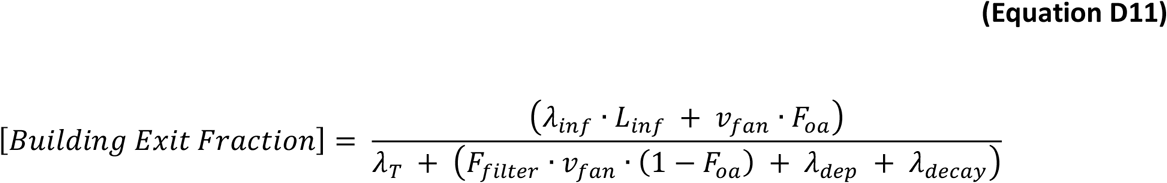

## Supplemental Material E: Outdoor Normalized Time and Space Integrated Air Concentrations

Outdoor normalized time and space integrated air concentration (Normalized TSIAC) values are a key component in our estimation of the single particle atmospheric transmission rates discussed in the main text. In this Supplemental Material, we describe how these values were calculated. For context, we also provide absolute and relative infection probabilities for a wider range of input parameter values than discussed in the main text.

We use the Lawrence Livermore National Laboratory Atmospheric Data Assimilation and Parameterization Techniques (ADAPT) and Lagrangian Operational Dispersion Integrator (LODI) meteorological and atmospheric dispersion models to calculate outdoor normalized time and space integrated air concentrations [242], [243]. The LODI/ADAPT model accuracy and validation are discussed in **Supplemental Material B: Key Atmospheric Transport and Dispersion Modeling Concepts**, *Atmospheric Dispersion Models* section.

We consider a suite of atmospheric conditions, **Table E1**, which were chosen to span a reasonably wide, but not comprehensive, range of common atmospheric conditions. The atmospheric conditions were held constant during the plume passage. Following this table, we provide the methodology and specific modeling assumptions used.

**Tables E2(a-b)** to **E5(a-b)** provide the predicted outdoor Normalized TSIAC as a function of distance from an emission source, atmospheric conditions, and airborne loss rate. The Normalized TSIAC values are specified for both discs and circle arcs at distances ranging from 50 m to 20 km. Each table corresponds to one airborne (infectivity) loss rate (0, 0.1, 1, or 10 h^-1^ loss rates were modeled). Each table column corresponds to 1 of 7 commonly encountered wind speeds and atmospheric stabilities as specified in **Table E1**.

For context, **Figures E1** to **E4** provide downwind, upper bound estimates for two cases: (a) the average per-person, single particle, absolute infection probability and (b) the corresponding urban area infections. **Figures E5** to **E8** provide relative infection probabilities relative to 1 km disc or arc values. Each figure corresponds to a single airborne (infectivity) loss rate.

**Table E1.** Summary of modeled atmospheric conditions

| Qualitative Description | Stability Class<br>(MO Length in m) | Wind Speed at 10 m<br>above ground level<br>in m s<sup>-1</sup> | Boundary Layer<br>Height in m |
| --- | --- | --- | --- |
| Clear, cold night with<br>light winds | F<br>(25) | 1.0 | 300 |
| Clear night<br>with gentle breeze | E<br>(50) | 4.5 | 500 |
| Overcast<br>with light winds | C<br>(-50) | 1.0 | 1,000 |
| Overcast<br>with gentle breeze | D<br>(∞) | 4.5 | 800 |
| Overcast<br>with strong breeze | D<br>(∞) | 10 | 800 |
| Clear day<br>with gentle breeze | B<br>(-25) | 4.5 | 1,200 |
| Clear, hot day<br>with light winds | A<br>(-10) | 1.0 | 1,500 |

We modeled the downwind impacts assuming:

- The release quantity is one (effective) particle.
- All particles have 1 μm aerodynamic diameter.
- The released particle has a constant probability of being emitted from a point within a 1 hr period.
- Gravitational settling is the only deposition mechanism considered. The inclusion of other deposition mechanisms including (but not limited to) Brownian diffusion, rainout, impaction, and interception would all serve to reduce the modeled air concentrations and so our neglect of these mechanisms is consistent with the intent of providing an upper bound on the number of downwind infections.
- 4 different 1^st^ order airborne loss rates (in addition to gravitational settling) are considered: 0, 0.1, 1, and 10 h^-1^. These airborne loss rates are used to characterize the effects of airborne loss of particle infectivity.
- Resuspension (the re-emission of deposited material back into the atmosphere) is neglected.
- Flat terrain with a surface roughness of 0.1 m corresponding to a suburban setting. In forested and downtown regions, the earth’s surface is typically rougher – resulting in lower surface air concentrations during stable and neutral atmospheric conditions.
- The modeled wind blew from the west (270°) to the east with no vertical or horizontal variation in wind direction (modeled wind speeds increase with height above ground).

Downwind concentrations were predicted using 2 modeling grids. The first had a 30 km horizontal extent and 100 m horizontal resolution grid cells. The second had a 1.5 km horizontal extent and 6 m horizontal resolution grid cells

For each case, LODI was run with 1,000,000 marker particles^41^ to calculate the (ensemble) average surface (lowest 20 m) normalized air concentrations integrated over the entire plume passage (i.e., until no marker particles remain in the modeling domain) for each model grid cell. For each atmospheric condition, the following procedure was used to calculate the normalized space and time integrated air concentration provided in **Tables E2(a-b)** to **E5(a-b)**.

> First, we developed an interpolation function for each modeling grid using the MATLAB *griddedInterpolant* function configured to return either the value at the nearest sample grid point to the requested location (if the requested location is within the sample grid) or zero (if the requested location is outside the sample grid).
>
> Second, we calculated the normalized time integrated air concentrations for a fixed set of radial distances by numerically integrating previously developed interpolation functions (using the MATLAB *integral* and *integral2* functions, as appropriate) for (a) a disk and (b) circle arc, both centered on the release. For the 1.5 km extent grid, the distances considered were 50 m to 1,100 m at 50 m increments. For the 30 km extent grid, the distances considered were 2,000 m to 20,000 m at 1,000 m increments.
>
> Third, we developed a second, linear interpolation function using the *griddedInterpolant* function from the distances considered and the normalized time integrated air concentrations calculated in step 2. This function was used for remainder of the analysis.

**Table E2a.** Normalized Time and Space Integrated Particle Air Concentration for a Disc with the specified distance from the release assuming no airborne loss rate (*λ*_*decay*_ = 0 h^-1^).

| Distance<br>Downwind<br>(m) | Clear, cold<br>night with<br>light winds<br>(s m<sup>-1</sup>) | Clear night<br>with<br>gentle<br>breeze<br>(s m<sup>-1</sup>) | Overcast<br>with light<br>winds<br>(s m<sup>-1</sup>) | Overcast<br>with<br>gentle<br>winds<br>(s m<sup>-1</sup>) | Overcast<br>with<br>strong<br>winds<br>(s m<sup>-1</sup>) | Clear day<br>with<br>gentle<br>winds<br>(s m<sup>-1</sup>) | Clear, hot<br>day with<br>light winds<br>(s m<sup>-1</sup>) |
| --- | --- | --- | --- | --- | --- | --- | --- |
| 50 | 2.2E+01 | 4.4E+00 | 1.1E+01 | 3.4E+00 | 1.5E+00 | 2.1E+00 | 7.1E+00 |
| 100 | 4.0E+01 | 8.0E+00 | 1.6E+01 | 5.6E+00 | 2.5E+00 | 3.0E+00 | 9.5E+00 |
| 150 | 5.6E+01 | 1.1E+01 | 1.9E+01 | 7.2E+00 | 3.3E+00 | 3.6E+00 | 1.1E+01 |
| 200 | 7.0E+01 | 1.3E+01 | 2.2E+01 | 8.4E+00 | 3.8E+00 | 3.9E+00 | 1.2E+01 |
| 250 | 8.3E+01 | 1.5E+01 | 2.4E+01 | 9.4E+00 | 4.3E+00 | 4.2E+00 | 1.3E+01 |
| 300 | 9.4E+01 | 1.7E+01 | 2.5E+01 | 1.0E+01 | 4.6E+00 | 4.5E+00 | 1.3E+01 |
| 350 | 1.0E+02 | 1.9E+01 | 2.6E+01 | 1.1E+01 | 5.0E+00 | 4.6E+00 | 1.4E+01 |
| 400 | 1.1E+02 | 2.0E+01 | 2.8E+01 | 1.2E+01 | 5.3E+00 | 4.8E+00 | 1.4E+01 |
| 450 | 1.2E+02 | 2.2E+01 | 2.9E+01 | 1.2E+01 | 5.5E+00 | 5.0E+00 | 1.4E+01 |
| 500 | 1.3E+02 | 2.3E+01 | 2.9E+01 | 1.3E+01 | 5.8E+00 | 5.1E+00 | 1.5E+01 |
| 550 | 1.4E+02 | 2.4E+01 | 3.0E+01 | 1.3E+01 | 6.0E+00 | 5.2E+00 | 1.5E+01 |
| 600 | 1.4E+02 | 2.5E+01 | 3.1E+01 | 1.4E+01 | 6.2E+00 | 5.3E+00 | 1.5E+01 |
| 650 | 1.5E+02 | 2.6E+01 | 3.1E+01 | 1.4E+01 | 6.4E+00 | 5.4E+00 | 1.5E+01 |
| 700 | 1.5E+02 | 2.7E+01 | 3.2E+01 | 1.4E+01 | 6.5E+00 | 5.5E+00 | 1.6E+01 |
| 750 | 1.6E+02 | 2.8E+01 | 3.2E+01 | 1.5E+01 | 6.7E+00 | 5.5E+00 | 1.6E+01 |
| 800 | 1.6E+02 | 2.9E+01 | 3.3E+01 | 1.5E+01 | 6.8E+00 | 5.6E+00 | 1.6E+01 |
| 850 | 1.7E+02 | 2.9E+01 | 3.3E+01 | 1.5E+01 | 7.0E+00 | 5.7E+00 | 1.6E+01 |
| 900 | 1.7E+02 | 3.0E+01 | 3.4E+01 | 1.6E+01 | 7.1E+00 | 5.7E+00 | 1.6E+01 |
| 950 | 1.8E+02 | 3.1E+01 | 3.4E+01 | 1.6E+01 | 7.2E+00 | 5.8E+00 | 1.6E+01 |
| 1,000 | 1.8E+02 | 3.1E+01 | 3.4E+01 | 1.6E+01 | 7.4E+00 | 5.8E+00 | 1.6E+01 |
| 1,050 | 1.8E+02 | 3.2E+01 | 3.5E+01 | 1.6E+01 | 7.5E+00 | 5.9E+00 | 1.7E+01 |
| 1,100 | 1.9E+02 | 3.3E+01 | 3.5E+01 | 1.7E+01 | 7.6E+00 | 5.9E+00 | 1.7E+01 |
| 2,000 | 2.3E+02 | 3.8E+01 | 3.6E+01 | 1.8E+01 | 8.1E+00 | 6.7E+00 | 1.9E+01 |
| 3,000 | 2.6E+02 | 4.2E+01 | 3.8E+01 | 1.9E+01 | 8.7E+00 | 7.1E+00 | 1.9E+01 |
| 4,000 | 2.8E+02 | 4.6E+01 | 4.0E+01 | 2.0E+01 | 9.2E+00 | 7.3E+00 | 2.0E+01 |
| 5,000 | 3.0E+02 | 4.8E+01 | 4.1E+01 | 2.1E+01 | 9.6E+00 | 7.5E+00 | 2.1E+01 |
| 6,000 | 3.1E+02 | 5.1E+01 | 4.2E+01 | 2.2E+01 | 9.9E+00 | 7.7E+00 | 2.1E+01 |
| 7,000 | 3.1E+02 | 5.3E+01 | 4.3E+01 | 2.2E+01 | 1.0E+01 | 7.9E+00 | 2.2E+01 |
| 8,000 | 3.2E+02 | 5.5E+01 | 4.4E+01 | 2.3E+01 | 1.0E+01 | 8.0E+00 | 2.2E+01 |
| 9,000 | 3.3E+02 | 5.7E+01 | 4.5E+01 | 2.3E+01 | 1.1E+01 | 8.2E+00 | 2.2E+01 |
| 10,000 | 3.3E+02 | 5.8E+01 | 4.5E+01 | 2.4E+01 | 1.1E+01 | 8.3E+00 | 2.3E+01 |
| 11,000 | 3.4E+02 | 6.0E+01 | 4.6E+01 | 2.4E+01 | 1.1E+01 | 8.4E+00 | 2.3E+01 |
| 12,000 | 3.4E+02 | 6.1E+01 | 4.7E+01 | 2.4E+01 | 1.1E+01 | 8.5E+00 | 2.4E+01 |
| 13,000 | 3.5E+02 | 6.2E+01 | 4.7E+01 | 2.5E+01 | 1.2E+01 | 8.7E+00 | 2.4E+01 |
| 14,000 | 3.5E+02 | 6.4E+01 | 4.8E+01 | 2.5E+01 | 1.2E+01 | 8.8E+00 | 2.4E+01 |
| 15,000 | 3.5E+02 | 6.5E+01 | 4.9E+01 | 2.6E+01 | 1.2E+01 | 8.9E+00 | 2.5E+01 |
| 16,000 | 3.5E+02 | 6.6E+01 | 4.9E+01 | 2.6E+01 | 1.2E+01 | 9.0E+00 | 2.5E+01 |
| 17,000 | 3.6E+02 | 6.7E+01 | 5.0E+01 | 2.6E+01 | 1.2E+01 | 9.1E+00 | 2.5E+01 |
| 18,000 | 3.6E+02 | 6.8E+01 | 5.0E+01 | 2.6E+01 | 1.2E+01 | 9.2E+00 | 2.6E+01 |
| 19,000 | 3.6E+02 | 6.9E+01 | 5.1E+01 | 2.7E+01 | 1.3E+01 | 9.3E+00 | 2.6E+01 |
| 20,000 | 3.6E+02 | 7.0E+01 | 5.1E+01 | 2.7E+01 | 1.3E+01 | 9.4E+00 | 2.7E+01 |

**Table E2b.** Normalized Time and Space Integrated Particle Air Concentration for a Circle Arc at the specified distance from the release assuming no airborne loss rate (*λ*_*decay*_ = 0 h^-1^).

| Distance<br>Downwind<br>(m) | Clear, cold<br>night with<br>light winds<br>(s m<sup>-2</sup>) | Clear night<br>with<br>gentle<br>breeze<br>(s m<sup>-2</sup>) | Overcast<br>with light<br>winds<br>(s m<sup>-2</sup>) | Overcast<br>with<br>gentle<br>winds<br>(s m<sup>-2</sup>) | Overcast<br>with<br>strong<br>winds<br>(s m<sup>-2</sup>) | Clear day<br>with<br>gentle<br>winds<br>(s m<sup>-2</sup>) | Clear, hot<br>day with<br>light winds<br>(s m<sup>-2</sup>) |
| --- | --- | --- | --- | --- | --- | --- | --- |
| 50 | 4.0E-01 | 8.0E-02 | 1.4E-01 | 5.4E-02 | 2.4E-02 | 2.4E-02 | 6.9E-02 |
| 100 | 3.4E-01 | 6.4E-02 | 8.1E-02 | 3.6E-02 | 1.6E-02 | 1.3E-02 | 3.5E-02 |
| 150 | 3.0E-01 | 5.3E-02 | 5.6E-02 | 2.8E-02 | 1.2E-02 | 8.9E-03 | 2.3E-02 |
| 200 | 2.6E-01 | 4.5E-02 | 4.2E-02 | 2.2E-02 | 9.9E-03 | 6.5E-03 | 1.6E-02 |
| 250 | 2.3E-01 | 3.9E-02 | 3.4E-02 | 1.8E-02 | 8.3E-03 | 5.1E-03 | 1.3E-02 |
| 300 | 2.1E-01 | 3.4E-02 | 2.8E-02 | 1.6E-02 | 7.1E-03 | 4.2E-03 | 1.0E-02 |
| 350 | 1.9E-01 | 3.1E-02 | 2.3E-02 | 1.4E-02 | 6.2E-03 | 3.5E-03 | 8.5E-03 |
| 400 | 1.7E-01 | 2.8E-02 | 2.0E-02 | 1.2E-02 | 5.5E-03 | 3.1E-03 | 7.2E-03 |
| 450 | 1.6E-01 | 2.5E-02 | 1.8E-02 | 1.1E-02 | 5.0E-03 | 2.7E-03 | 6.2E-03 |
| 500 | 1.5E-01 | 2.3E-02 | 1.6E-02 | 9.8E-03 | 4.5E-03 | 2.3E-03 | 5.5E-03 |
| 550 | 1.4E-01 | 2.2E-02 | 1.4E-02 | 9.0E-03 | 4.1E-03 | 2.1E-03 | 4.9E-03 |
| 600 | 1.3E-01 | 2.0E-02 | 1.3E-02 | 8.3E-03 | 3.8E-03 | 1.9E-03 | 4.4E-03 |
| 650 | 1.2E-01 | 1.9E-02 | 1.2E-02 | 7.7E-03 | 3.5E-03 | 1.7E-03 | 3.9E-03 |
| 700 | 1.1E-01 | 1.8E-02 | 1.1E-02 | 7.3E-03 | 3.3E-03 | 1.6E-03 | 3.6E-03 |
| 750 | 1.0E-01 | 1.7E-02 | 9.8E-03 | 6.8E-03 | 3.1E-03 | 1.4E-03 | 3.3E-03 |
| 800 | 9.6E-02 | 1.6E-02 | 9.0E-03 | 6.4E-03 | 2.9E-03 | 1.3E-03 | 3.1E-03 |
| 850 | 9.0E-02 | 1.5E-02 | 8.3E-03 | 6.0E-03 | 2.8E-03 | 1.3E-03 | 2.9E-03 |
| 900 | 8.4E-02 | 1.4E-02 | 7.7E-03 | 5.8E-03 | 2.6E-03 | 1.2E-03 | 2.7E-03 |
| 950 | 7.8E-02 | 1.4E-02 | 7.2E-03 | 5.5E-03 | 2.5E-03 | 1.1E-03 | 2.5E-03 |
| 1,000 | 7.2E-02 | 1.3E-02 | 6.7E-03 | 5.2E-03 | 2.4E-03 | 1.0E-03 | 2.3E-03 |
| 1,050 | 6.7E-02 | 1.2E-02 | 6.1E-03 | 4.9E-03 | 2.3E-03 | 9.7E-04 | 2.2E-03 |
| 1,100 | 5.9E-02 | 1.2E-02 | 5.5E-03 | 4.7E-03 | 2.2E-03 | 9.0E-04 | 2.0E-03 |
| 2,000 | 3.8E-02 | 5.5E-03 | 2.6E-03 | 1.7E-03 | 7.7E-04 | 4.5E-04 | 1.1E-03 |
| 3,000 | 2.3E-02 | 4.0E-03 | 1.7E-03 | 1.2E-03 | 5.3E-04 | 2.9E-04 | 7.3E-04 |
| 4,000 | 1.6E-02 | 3.1E-03 | 1.3E-03 | 8.9E-04 | 4.2E-04 | 2.3E-04 | 5.9E-04 |
| 5,000 | 1.2E-02 | 2.6E-03 | 1.1E-03 | 7.4E-04 | 3.6E-04 | 1.9E-04 | 5.2E-04 |
| 6,000 | 9.5E-03 | 2.3E-03 | 9.6E-04 | 6.4E-04 | 3.2E-04 | 1.7E-04 | 4.6E-04 |
| 7,000 | 7.8E-03 | 2.0E-03 | 8.6E-04 | 5.6E-04 | 2.8E-04 | 1.5E-04 | 4.4E-04 |
| 8,000 | 6.6E-03 | 1.8E-03 | 8.1E-04 | 5.1E-04 | 2.6E-04 | 1.4E-04 | 4.3E-04 |
| 9,000 | 5.7E-03 | 1.7E-03 | 7.6E-04 | 4.7E-04 | 2.4E-04 | 1.3E-04 | 4.0E-04 |
| 10,000 | 4.9E-03 | 1.5E-03 | 7.0E-04 | 4.3E-04 | 2.3E-04 | 1.3E-04 | 3.8E-04 |
| 11,000 | 4.4E-03 | 1.4E-03 | 6.9E-04 | 4.1E-04 | 2.2E-04 | 1.3E-04 | 3.7E-04 |
| 12,000 | 3.9E-03 | 1.4E-03 | 6.7E-04 | 3.9E-04 | 2.1E-04 | 1.2E-04 | 3.7E-04 |
| 13,000 | 3.6E-03 | 1.3E-03 | 6.3E-04 | 3.7E-04 | 2.0E-04 | 1.2E-04 | 3.7E-04 |
| 14,000 | 3.2E-03 | 1.2E-03 | 6.1E-04 | 3.5E-04 | 1.9E-04 | 1.1E-04 | 3.6E-04 |
| 15,000 | 3.0E-03 | 1.2E-03 | 5.9E-04 | 3.3E-04 | 1.8E-04 | 1.1E-04 | 3.6E-04 |
| 16,000 | 2.7E-03 | 1.1E-03 | 5.7E-04 | 3.2E-04 | 1.7E-04 | 1.1E-04 | 3.6E-04 |
| 17,000 | 2.5E-03 | 1.1E-03 | 5.9E-04 | 3.1E-04 | 1.7E-04 | 1.1E-04 | 3.6E-04 |
| 18,000 | 2.4E-03 | 1.1E-03 | 5.7E-04 | 2.9E-04 | 1.7E-04 | 1.0E-04 | 3.5E-04 |
| 19,000 | 2.2E-03 | 1.0E-03 | 5.4E-04 | 2.8E-04 | 1.6E-04 | 1.1E-04 | 3.5E-04 |
| 20,000 | 2.1E-03 | 9.9E-04 | 5.2E-04 | 2.8E-04 | 1.6E-04 | 1.0E-04 | 3.6E-04 |

**Table E3a.** Normalized Time and Space Integrated Particle Air Concentration for a Disc with the specified distance from the release assuming airborne loss rate (*λ*_*decay*_ = 0.1 h^-1^).

| Distance<br>Downwind<br>(m) | Clear, cold<br>night with<br>light winds<br>(s m<sup>-1</sup>) | Clear night<br>with<br>gentle<br>breeze<br>(s m<sup>-1</sup>) | Overcast<br>with light<br>winds<br>(s m<sup>-1</sup>) | Overcast<br>with<br>gentle<br>winds<br>(s m<sup>-1</sup>) | Overcast<br>with<br>strong<br>winds<br>(s m<sup>-1</sup>) | Clear day<br>with<br>gentle<br>winds<br>(s m<sup>-1</sup>) | Clear, hot<br>day with<br>light winds<br>(s m<sup>-1</sup>) |
| --- | --- | --- | --- | --- | --- | --- | --- |
| 50 | 2.2E+01 | 4.4E+00 | 1.1E+01 | 3.4E+00 | 1.5E+00 | 2.1E+00 | 7.1E+00 |
| 100 | 4.0E+01 | 8.0E+00 | 1.6E+01 | 5.6E+00 | 2.5E+00 | 3.0E+00 | 9.5E+00 |
| 150 | 5.6E+01 | 1.1E+01 | 1.9E+01 | 7.2E+00 | 3.3E+00 | 3.6E+00 | 1.1E+01 |
| 200 | 7.0E+01 | 1.3E+01 | 2.2E+01 | 8.4E+00 | 3.8E+00 | 3.9E+00 | 1.2E+01 |
| 250 | 8.2E+01 | 1.5E+01 | 2.4E+01 | 9.4E+00 | 4.3E+00 | 4.2E+00 | 1.3E+01 |
| 300 | 9.3E+01 | 1.7E+01 | 2.5E+01 | 1.0E+01 | 4.6E+00 | 4.5E+00 | 1.3E+01 |
| 350 | 1.0E+02 | 1.9E+01 | 2.6E+01 | 1.1E+01 | 5.0E+00 | 4.6E+00 | 1.4E+01 |
| 400 | 1.1E+02 | 2.0E+01 | 2.7E+01 | 1.2E+01 | 5.3E+00 | 4.8E+00 | 1.4E+01 |
| 450 | 1.2E+02 | 2.2E+01 | 2.8E+01 | 1.2E+01 | 5.5E+00 | 5.0E+00 | 1.4E+01 |
| 500 | 1.3E+02 | 2.3E+01 | 2.9E+01 | 1.3E+01 | 5.8E+00 | 5.1E+00 | 1.5E+01 |
| 550 | 1.3E+02 | 2.4E+01 | 3.0E+01 | 1.3E+01 | 6.0E+00 | 5.2E+00 | 1.5E+01 |
| 600 | 1.4E+02 | 2.5E+01 | 3.1E+01 | 1.4E+01 | 6.2E+00 | 5.3E+00 | 1.5E+01 |
| 650 | 1.5E+02 | 2.6E+01 | 3.1E+01 | 1.4E+01 | 6.3E+00 | 5.4E+00 | 1.5E+01 |
| 700 | 1.5E+02 | 2.7E+01 | 3.2E+01 | 1.4E+01 | 6.5E+00 | 5.5E+00 | 1.5E+01 |
| 750 | 1.6E+02 | 2.8E+01 | 3.2E+01 | 1.5E+01 | 6.7E+00 | 5.5E+00 | 1.6E+01 |
| 800 | 1.6E+02 | 2.8E+01 | 3.3E+01 | 1.5E+01 | 6.8E+00 | 5.6E+00 | 1.6E+01 |
| 850 | 1.7E+02 | 2.9E+01 | 3.3E+01 | 1.5E+01 | 7.0E+00 | 5.7E+00 | 1.6E+01 |
| 900 | 1.7E+02 | 3.0E+01 | 3.3E+01 | 1.6E+01 | 7.1E+00 | 5.7E+00 | 1.6E+01 |
| 950 | 1.7E+02 | 3.1E+01 | 3.4E+01 | 1.6E+01 | 7.2E+00 | 5.8E+00 | 1.6E+01 |
| 1,000 | 1.8E+02 | 3.1E+01 | 3.4E+01 | 1.6E+01 | 7.4E+00 | 5.8E+00 | 1.6E+01 |
| 1,050 | 1.8E+02 | 3.2E+01 | 3.5E+01 | 1.6E+01 | 7.5E+00 | 5.9E+00 | 1.6E+01 |
| 1,100 | 1.8E+02 | 3.2E+01 | 3.5E+01 | 1.7E+01 | 7.6E+00 | 5.9E+00 | 1.7E+01 |
| 2,000 | 2.3E+02 | 3.7E+01 | 3.6E+01 | 1.8E+01 | 8.1E+00 | 6.7E+00 | 1.8E+01 |
| 3,000 | 2.5E+02 | 4.2E+01 | 3.8E+01 | 1.9E+01 | 8.7E+00 | 7.1E+00 | 1.9E+01 |
| 4,000 | 2.7E+02 | 4.5E+01 | 3.9E+01 | 2.0E+01 | 9.1E+00 | 7.3E+00 | 2.0E+01 |
| 5,000 | 2.8E+02 | 4.8E+01 | 4.0E+01 | 2.1E+01 | 9.5E+00 | 7.5E+00 | 2.0E+01 |
| 6,000 | 2.9E+02 | 5.0E+01 | 4.1E+01 | 2.1E+01 | 9.9E+00 | 7.7E+00 | 2.1E+01 |
| 7,000 | 3.0E+02 | 5.2E+01 | 4.2E+01 | 2.2E+01 | 1.0E+01 | 7.8E+00 | 2.1E+01 |
| 8,000 | 3.0E+02 | 5.4E+01 | 4.3E+01 | 2.2E+01 | 1.0E+01 | 8.0E+00 | 2.2E+01 |
| 9,000 | 3.1E+02 | 5.6E+01 | 4.3E+01 | 2.3E+01 | 1.1E+01 | 8.1E+00 | 2.2E+01 |
| 10,000 | 3.1E+02 | 5.7E+01 | 4.4E+01 | 2.3E+01 | 1.1E+01 | 8.2E+00 | 2.2E+01 |
| 11,000 | 3.1E+02 | 5.9E+01 | 4.4E+01 | 2.4E+01 | 1.1E+01 | 8.4E+00 | 2.3E+01 |
| 12,000 | 3.2E+02 | 6.0E+01 | 4.5E+01 | 2.4E+01 | 1.1E+01 | 8.5E+00 | 2.3E+01 |
| 13,000 | 3.2E+02 | 6.1E+01 | 4.5E+01 | 2.4E+01 | 1.1E+01 | 8.6E+00 | 2.3E+01 |
| 14,000 | 3.2E+02 | 6.2E+01 | 4.6E+01 | 2.5E+01 | 1.2E+01 | 8.7E+00 | 2.3E+01 |
| 15,000 | 3.2E+02 | 6.4E+01 | 4.6E+01 | 2.5E+01 | 1.2E+01 | 8.8E+00 | 2.4E+01 |
| 16,000 | 3.3E+02 | 6.5E+01 | 4.7E+01 | 2.5E+01 | 1.2E+01 | 8.9E+00 | 2.4E+01 |
| 17,000 | 3.3E+02 | 6.6E+01 | 4.7E+01 | 2.6E+01 | 1.2E+01 | 9.0E+00 | 2.4E+01 |
| 18,000 | 3.3E+02 | 6.7E+01 | 4.8E+01 | 2.6E+01 | 1.2E+01 | 9.1E+00 | 2.4E+01 |
| 19,000 | 3.3E+02 | 6.8E+01 | 4.8E+01 | 2.6E+01 | 1.3E+01 | 9.2E+00 | 2.5E+01 |
| 20,000 | 3.3E+02 | 6.8E+01 | 4.8E+01 | 2.7E+01 | 1.3E+01 | 9.3E+00 | 2.5E+01 |

**Table E3b.** Normalized Time and Space Integrated Particle Air Concentration for a Circle Arc at the specified distance from the release assuming airborne loss rate (*λ*_*decay*_ = 0.1 h^-1^).

| Distance<br>Downwind<br>(m) | Clear, cold<br>night with<br>light winds<br>(s m<sup>-2</sup>) | Clear night<br>with<br>gentle<br>breeze<br>(s m<sup>-2</sup>) | Overcast<br>with light<br>winds<br>(s m<sup>-2</sup>) | Overcast<br>with<br>gentle<br>winds<br>(s m<sup>-2</sup>) | Overcast<br>with<br>strong<br>winds<br>(s m<sup>-2</sup>) | Clear day<br>with<br>gentle<br>winds<br>(s m<sup>-2</sup>) | Clear, hot<br>day with<br>light winds<br>(s m<sup>-2</sup>) |
| --- | --- | --- | --- | --- | --- | --- | --- |
| 50 | 4.0E-01 | 8.0E-02 | 1.3E-01 | 5.4E-02 | 2.4E-02 | 2.4E-02 | 6.9E-02 |
| 100 | 3.4E-01 | 6.4E-02 | 8.0E-02 | 3.6E-02 | 1.6E-02 | 1.3E-02 | 3.5E-02 |
| 150 | 3.0E-01 | 5.3E-02 | 5.6E-02 | 2.8E-02 | 1.2E-02 | 8.9E-03 | 2.3E-02 |
| 200 | 2.6E-01 | 4.4E-02 | 4.2E-02 | 2.2E-02 | 9.9E-03 | 6.5E-03 | 1.6E-02 |
| 250 | 2.3E-01 | 3.9E-02 | 3.3E-02 | 1.8E-02 | 8.2E-03 | 5.1E-03 | 1.3E-02 |
| 300 | 2.1E-01 | 3.4E-02 | 2.8E-02 | 1.6E-02 | 7.1E-03 | 4.2E-03 | 1.0E-02 |
| 350 | 1.9E-01 | 3.1E-02 | 2.3E-02 | 1.4E-02 | 6.2E-03 | 3.5E-03 | 8.4E-03 |
| 400 | 1.7E-01 | 2.8E-02 | 2.0E-02 | 1.2E-02 | 5.5E-03 | 3.1E-03 | 7.1E-03 |
| 450 | 1.5E-01 | 2.5E-02 | 1.7E-02 | 1.1E-02 | 4.9E-03 | 2.7E-03 | 6.1E-03 |
| 500 | 1.4E-01 | 2.3E-02 | 1.5E-02 | 9.8E-03 | 4.5E-03 | 2.3E-03 | 5.4E-03 |
| 550 | 1.3E-01 | 2.1E-02 | 1.4E-02 | 9.0E-03 | 4.1E-03 | 2.1E-03 | 4.8E-03 |
| 600 | 1.2E-01 | 2.0E-02 | 1.3E-02 | 8.3E-03 | 3.8E-03 | 1.9E-03 | 4.3E-03 |
| 650 | 1.1E-01 | 1.9E-02 | 1.1E-02 | 7.7E-03 | 3.5E-03 | 1.7E-03 | 3.8E-03 |
| 700 | 1.1E-01 | 1.7E-02 | 1.0E-02 | 7.2E-03 | 3.3E-03 | 1.6E-03 | 3.5E-03 |
| 750 | 1.0E-01 | 1.7E-02 | 9.6E-03 | 6.7E-03 | 3.1E-03 | 1.4E-03 | 3.3E-03 |
| 800 | 9.3E-02 | 1.6E-02 | 8.8E-03 | 6.3E-03 | 2.9E-03 | 1.3E-03 | 3.0E-03 |
| 850 | 8.6E-02 | 1.5E-02 | 8.1E-03 | 6.0E-03 | 2.8E-03 | 1.2E-03 | 2.8E-03 |
| 900 | 8.0E-02 | 1.4E-02 | 7.5E-03 | 5.7E-03 | 2.6E-03 | 1.2E-03 | 2.6E-03 |
| 950 | 7.5E-02 | 1.3E-02 | 7.0E-03 | 5.4E-03 | 2.5E-03 | 1.1E-03 | 2.4E-03 |
| 1,000 | 6.9E-02 | 1.3E-02 | 6.5E-03 | 5.1E-03 | 2.4E-03 | 1.0E-03 | 2.3E-03 |
| 1,050 | 6.3E-02 | 1.2E-02 | 6.0E-03 | 4.9E-03 | 2.3E-03 | 9.7E-04 | 2.1E-03 |
| 1,100 | 5.6E-02 | 1.2E-02 | 5.3E-03 | 4.6E-03 | 2.1E-03 | 8.9E-04 | 1.9E-03 |
| 2,000 | 3.5E-02 | 5.4E-03 | 2.4E-03 | 1.6E-03 | 7.6E-04 | 4.4E-04 | 1.0E-03 |
| 3,000 | 2.1E-02 | 3.9E-03 | 1.6E-03 | 1.1E-03 | 5.3E-04 | 2.9E-04 | 6.8E-04 |
| 4,000 | 1.4E-02 | 3.1E-03 | 1.2E-03 | 8.7E-04 | 4.2E-04 | 2.2E-04 | 5.3E-04 |
| 5,000 | 1.0E-02 | 2.5E-03 | 9.6E-04 | 7.2E-04 | 3.5E-04 | 1.9E-04 | 4.6E-04 |
| 6,000 | 7.8E-03 | 2.2E-03 | 8.3E-04 | 6.2E-04 | 3.1E-04 | 1.6E-04 | 4.0E-04 |
| 7,000 | 6.2E-03 | 1.9E-03 | 7.3E-04 | 5.4E-04 | 2.8E-04 | 1.5E-04 | 3.7E-04 |
| 8,000 | 5.2E-03 | 1.7E-03 | 6.7E-04 | 4.8E-04 | 2.6E-04 | 1.3E-04 | 3.6E-04 |
| 9,000 | 4.3E-03 | 1.6E-03 | 6.2E-04 | 4.5E-04 | 2.4E-04 | 1.3E-04 | 3.3E-04 |
| 10,000 | 3.7E-03 | 1.5E-03 | 5.6E-04 | 4.1E-04 | 2.2E-04 | 1.2E-04 | 3.0E-04 |
| 11,000 | 3.2E-03 | 1.4E-03 | 5.4E-04 | 3.8E-04 | 2.1E-04 | 1.2E-04 | 2.9E-04 |
| 12,000 | 2.8E-03 | 1.3E-03 | 5.1E-04 | 3.7E-04 | 2.0E-04 | 1.1E-04 | 2.8E-04 |
| 13,000 | 2.5E-03 | 1.2E-03 | 4.7E-04 | 3.5E-04 | 1.9E-04 | 1.1E-04 | 2.8E-04 |
| 14,000 | 2.2E-03 | 1.1E-03 | 4.4E-04 | 3.3E-04 | 1.9E-04 | 1.1E-04 | 2.6E-04 |
| 15,000 | 2.0E-03 | 1.1E-03 | 4.2E-04 | 3.1E-04 | 1.8E-04 | 1.0E-04 | 2.5E-04 |
| 16,000 | 1.8E-03 | 1.0E-03 | 4.0E-04 | 2.9E-04 | 1.7E-04 | 9.9E-05 | 2.5E-04 |
| 17,000 | 1.7E-03 | 1.0E-03 | 4.0E-04 | 2.8E-04 | 1.7E-04 | 1.0E-04 | 2.4E-04 |
| 18,000 | 1.5E-03 | 9.6E-04 | 3.8E-04 | 2.7E-04 | 1.6E-04 | 9.4E-05 | 2.3E-04 |
| 19,000 | 1.4E-03 | 9.2E-04 | 3.5E-04 | 2.5E-04 | 1.6E-04 | 9.9E-05 | 2.3E-04 |
| 20,000 | 1.3E-03 | 8.9E-04 | 3.3E-04 | 2.5E-04 | 1.5E-04 | 9.4E-05 | 2.3E-04 |

**Table E4a.** Normalized Time and Space Integrated Particle Air Concentration for a Disc with the specified distance from the release assuming airborne loss rate (*λ*_*decay*_ = 1 h^-1^).

| Distance<br>Downwind<br>(m) | Clear, cold<br>night with<br>light winds<br>(s m<sup>-1</sup>) | Clear night<br>with<br>gentle<br>breeze<br>(s m<sup>-1</sup>) | Overcast<br>with light<br>winds<br>(s m<sup>-1</sup>) | Overcast<br>with<br>gentle<br>winds<br>(s m<sup>-1</sup>) | Overcast<br>with<br>strong<br>winds<br>(s m<sup>-1</sup>) | Clear day<br>with<br>gentle<br>winds<br>(s m<sup>-1</sup>) | Clear, hot<br>day with<br>light winds<br>(s m<sup>-1</sup>) |
| --- | --- | --- | --- | --- | --- | --- | --- |
| 50 | 2.1E+01 | 4.4E+00 | 1.1E+01 | 3.4E+00 | 1.5E+00 | 2.1E+00 | 7.0E+00 |
| 100 | 3.9E+01 | 7.9E+00 | 1.6E+01 | 5.6E+00 | 2.5E+00 | 3.0E+00 | 9.4E+00 |
| 150 | 5.3E+01 | 1.1E+01 | 1.9E+01 | 7.2E+00 | 3.2E+00 | 3.5E+00 | 1.1E+01 |
| 200 | 6.6E+01 | 1.3E+01 | 2.1E+01 | 8.4E+00 | 3.8E+00 | 3.9E+00 | 1.2E+01 |
| 250 | 7.6E+01 | 1.5E+01 | 2.3E+01 | 9.3E+00 | 4.2E+00 | 4.2E+00 | 1.2E+01 |
| 300 | 8.6E+01 | 1.7E+01 | 2.4E+01 | 1.0E+01 | 4.6E+00 | 4.4E+00 | 1.3E+01 |
| 350 | 9.4E+01 | 1.9E+01 | 2.5E+01 | 1.1E+01 | 4.9E+00 | 4.6E+00 | 1.3E+01 |
| 400 | 1.0E+02 | 2.0E+01 | 2.6E+01 | 1.1E+01 | 5.2E+00 | 4.8E+00 | 1.4E+01 |
| 450 | 1.1E+02 | 2.1E+01 | 2.7E+01 | 1.2E+01 | 5.5E+00 | 4.9E+00 | 1.4E+01 |
| 500 | 1.1E+02 | 2.2E+01 | 2.8E+01 | 1.3E+01 | 5.7E+00 | 5.0E+00 | 1.4E+01 |
| 550 | 1.2E+02 | 2.3E+01 | 2.9E+01 | 1.3E+01 | 5.9E+00 | 5.1E+00 | 1.4E+01 |
| 600 | 1.2E+02 | 2.4E+01 | 2.9E+01 | 1.3E+01 | 6.1E+00 | 5.2E+00 | 1.5E+01 |
| 650 | 1.3E+02 | 2.5E+01 | 3.0E+01 | 1.4E+01 | 6.3E+00 | 5.3E+00 | 1.5E+01 |
| 700 | 1.3E+02 | 2.6E+01 | 3.0E+01 | 1.4E+01 | 6.5E+00 | 5.4E+00 | 1.5E+01 |
| 750 | 1.4E+02 | 2.7E+01 | 3.1E+01 | 1.4E+01 | 6.6E+00 | 5.5E+00 | 1.5E+01 |
| 800 | 1.4E+02 | 2.8E+01 | 3.1E+01 | 1.5E+01 | 6.8E+00 | 5.5E+00 | 1.5E+01 |
| 850 | 1.4E+02 | 2.8E+01 | 3.1E+01 | 1.5E+01 | 6.9E+00 | 5.6E+00 | 1.5E+01 |
| 900 | 1.5E+02 | 2.9E+01 | 3.2E+01 | 1.5E+01 | 7.0E+00 | 5.7E+00 | 1.5E+01 |
| 950 | 1.5E+02 | 3.0E+01 | 3.2E+01 | 1.6E+01 | 7.2E+00 | 5.7E+00 | 1.6E+01 |
| 1,000 | 1.5E+02 | 3.0E+01 | 3.2E+01 | 1.6E+01 | 7.3E+00 | 5.8E+00 | 1.6E+01 |
| 1,050 | 1.5E+02 | 3.1E+01 | 3.2E+01 | 1.6E+01 | 7.4E+00 | 5.8E+00 | 1.6E+01 |
| 1,100 | 1.5E+02 | 3.1E+01 | 3.3E+01 | 1.6E+01 | 7.5E+00 | 5.9E+00 | 1.6E+01 |
| 2,000 | 1.8E+02 | 3.6E+01 | 3.3E+01 | 1.7E+01 | 7.9E+00 | 6.6E+00 | 1.7E+01 |
| 3,000 | 1.9E+02 | 3.9E+01 | 3.4E+01 | 1.8E+01 | 8.5E+00 | 6.9E+00 | 1.8E+01 |
| 4,000 | 1.9E+02 | 4.2E+01 | 3.5E+01 | 1.9E+01 | 8.9E+00 | 7.1E+00 | 1.8E+01 |
| 5,000 | 2.0E+02 | 4.4E+01 | 3.5E+01 | 2.0E+01 | 9.3E+00 | 7.3E+00 | 1.8E+01 |
| 6,000 | 2.0E+02 | 4.6E+01 | 3.5E+01 | 2.0E+01 | 9.6E+00 | 7.4E+00 | 1.8E+01 |
| 7,000 | 2.0E+02 | 4.7E+01 | 3.6E+01 | 2.1E+01 | 9.8E+00 | 7.5E+00 | 1.8E+01 |
| 8,000 | 2.0E+02 | 4.8E+01 | 3.6E+01 | 2.1E+01 | 1.0E+01 | 7.6E+00 | 1.9E+01 |
| 9,000 | 2.0E+02 | 4.9E+01 | 3.6E+01 | 2.1E+01 | 1.0E+01 | 7.7E+00 | 1.9E+01 |
| 10,000 | 2.0E+02 | 5.0E+01 | 3.6E+01 | 2.1E+01 | 1.0E+01 | 7.8E+00 | 1.9E+01 |
| 11,000 | 2.0E+02 | 5.1E+01 | 3.6E+01 | 2.2E+01 | 1.1E+01 | 7.8E+00 | 1.9E+01 |
| 12,000 | 2.0E+02 | 5.2E+01 | 3.6E+01 | 2.2E+01 | 1.1E+01 | 7.9E+00 | 1.9E+01 |
| 13,000 | 2.0E+02 | 5.2E+01 | 3.6E+01 | 2.2E+01 | 1.1E+01 | 8.0E+00 | 1.9E+01 |
| 14,000 | 2.0E+02 | 5.3E+01 | 3.6E+01 | 2.2E+01 | 1.1E+01 | 8.0E+00 | 1.9E+01 |
| 15,000 | 2.0E+02 | 5.4E+01 | 3.6E+01 | 2.2E+01 | 1.1E+01 | 8.1E+00 | 1.9E+01 |
| 16,000 | 2.0E+02 | 5.4E+01 | 3.6E+01 | 2.3E+01 | 1.1E+01 | 8.1E+00 | 1.9E+01 |
| 17,000 | 2.0E+02 | 5.5E+01 | 3.6E+01 | 2.3E+01 | 1.1E+01 | 8.2E+00 | 1.9E+01 |
| 18,000 | 2.0E+02 | 5.5E+01 | 3.6E+01 | 2.3E+01 | 1.2E+01 | 8.2E+00 | 1.9E+01 |
| 19,000 | 2.0E+02 | 5.5E+01 | 3.6E+01 | 2.3E+01 | 1.2E+01 | 8.3E+00 | 1.9E+01 |
| 20,000 | 2.0E+02 | 5.6E+01 | 3.6E+01 | 2.3E+01 | 1.2E+01 | 8.3E+00 | 1.9E+01 |

**Table E4b.** Normalized Time and Space Integrated Particle Air Concentration for a Circle Arc at the specified distance from the release assuming airborne loss rate (*λ*_*decay*_ = 1 h^-1^).

| Distance<br>Downwind<br>(m) | Clear, cold<br>night with<br>light winds<br>(s m<sup>-2</sup>) | Clear night<br>with<br>gentle<br>breeze<br>(s m<sup>-2</sup>) | Overcast<br>with light<br>winds<br>(s m<sup>-2</sup>) | Overcast<br>with<br>gentle<br>winds<br>(s m<sup>-2</sup>) | Overcast<br>with<br>strong<br>winds<br>(s m<sup>-2</sup>) | Clear day<br>with<br>gentle<br>winds<br>(s m<sup>-2</sup>) | Clear, hot<br>day with<br>light winds<br>(s m<sup>-2</sup>) |
| --- | --- | --- | --- | --- | --- | --- | --- |
| 50 | 3.8E-01 | 8.0E-02 | 1.3E-01 | 5.4E-02 | 2.4E-02 | 2.4E-02 | 6.8E-02 |
| 100 | 3.2E-01 | 6.3E-02 | 7.8E-02 | 3.6E-02 | 1.6E-02 | 1.3E-02 | 3.4E-02 |
| 150 | 2.7E-01 | 5.2E-02 | 5.3E-02 | 2.7E-02 | 1.2E-02 | 8.8E-03 | 2.2E-02 |
| 200 | 2.3E-01 | 4.3E-02 | 3.9E-02 | 2.1E-02 | 9.8E-03 | 6.4E-03 | 1.5E-02 |
| 250 | 2.0E-01 | 3.8E-02 | 3.1E-02 | 1.8E-02 | 8.2E-03 | 5.0E-03 | 1.2E-02 |
| 300 | 1.8E-01 | 3.3E-02 | 2.5E-02 | 1.5E-02 | 7.0E-03 | 4.1E-03 | 9.4E-03 |
| 350 | 1.6E-01 | 2.9E-02 | 2.1E-02 | 1.3E-02 | 6.1E-03 | 3.4E-03 | 7.6E-03 |
| 400 | 1.4E-01 | 2.7E-02 | 1.8E-02 | 1.2E-02 | 5.4E-03 | 3.0E-03 | 6.4E-03 |
| 450 | 1.2E-01 | 2.4E-02 | 1.5E-02 | 1.0E-02 | 4.9E-03 | 2.6E-03 | 5.5E-03 |
| 500 | 1.1E-01 | 2.2E-02 | 1.3E-02 | 9.4E-03 | 4.4E-03 | 2.3E-03 | 4.8E-03 |
| 550 | 1.0E-01 | 2.0E-02 | 1.2E-02 | 8.6E-03 | 4.0E-03 | 2.0E-03 | 4.2E-03 |
| 600 | 9.3E-02 | 1.9E-02 | 1.1E-02 | 7.9E-03 | 3.7E-03 | 1.8E-03 | 3.7E-03 |
| 650 | 8.5E-02 | 1.8E-02 | 9.5E-03 | 7.3E-03 | 3.5E-03 | 1.7E-03 | 3.3E-03 |
| 700 | 7.8E-02 | 1.6E-02 | 8.6E-03 | 6.9E-03 | 3.2E-03 | 1.5E-03 | 3.0E-03 |
| 750 | 7.2E-02 | 1.5E-02 | 7.8E-03 | 6.4E-03 | 3.0E-03 | 1.4E-03 | 2.7E-03 |
| 800 | 6.5E-02 | 1.4E-02 | 7.1E-03 | 6.0E-03 | 2.9E-03 | 1.3E-03 | 2.5E-03 |
| 850 | 6.0E-02 | 1.4E-02 | 6.5E-03 | 5.6E-03 | 2.7E-03 | 1.2E-03 | 2.3E-03 |
| 900 | 5.5E-02 | 1.3E-02 | 6.0E-03 | 5.4E-03 | 2.6E-03 | 1.1E-03 | 2.1E-03 |
| 950 | 5.0E-02 | 1.2E-02 | 5.5E-03 | 5.1E-03 | 2.4E-03 | 1.0E-03 | 2.0E-03 |
| 1,000 | 4.6E-02 | 1.2E-02 | 5.0E-03 | 4.8E-03 | 2.3E-03 | 9.7E-04 | 1.8E-03 |
| 1,050 | 4.1E-02 | 1.1E-02 | 4.6E-03 | 4.6E-03 | 2.2E-03 | 9.1E-04 | 1.7E-03 |
| 1,100 | 3.6E-02 | 1.1E-02 | 4.1E-03 | 4.3E-03 | 2.1E-03 | 8.4E-04 | 1.5E-03 |
| 2,000 | 1.7E-02 | 4.7E-03 | 1.5E-03 | 1.4E-03 | 7.2E-04 | 4.0E-04 | 6.4E-04 |
| 3,000 | 7.5E-03 | 3.1E-03 | 8.0E-04 | 9.5E-04 | 4.9E-04 | 2.5E-04 | 3.5E-04 |
| 4,000 | 3.9E-03 | 2.3E-03 | 4.9E-04 | 6.9E-04 | 3.8E-04 | 1.8E-04 | 2.2E-04 |
| 5,000 | 2.3E-03 | 1.9E-03 | 3.3E-04 | 5.5E-04 | 3.1E-04 | 1.5E-04 | 1.6E-04 |
| 6,000 | 1.4E-03 | 1.5E-03 | 2.3E-04 | 4.5E-04 | 2.7E-04 | 1.2E-04 | 1.1E-04 |
| 7,000 | 9.3E-04 | 1.3E-03 | 1.7E-04 | 3.8E-04 | 2.3E-04 | 1.1E-04 | 8.4E-05 |
| 8,000 | 6.4E-04 | 1.1E-03 | 1.3E-04 | 3.2E-04 | 2.1E-04 | 9.1E-05 | 6.6E-05 |
| 9,000 | 4.5E-04 | 9.7E-04 | 1.0E-04 | 2.9E-04 | 1.9E-04 | 8.4E-05 | 5.0E-05 |
| 10,000 | 3.2E-04 | 8.6E-04 | 7.4E-05 | 2.5E-04 | 1.8E-04 | 7.7E-05 | 3.8E-05 |
| 11,000 | 2.3E-04 | 7.7E-04 | 5.9E-05 | 2.3E-04 | 1.7E-04 | 7.2E-05 | 2.9E-05 |
| 12,000 | 1.7E-04 | 6.9E-04 | 4.7E-05 | 2.1E-04 | 1.6E-04 | 6.5E-05 | 2.3E-05 |
| 13,000 | 1.3E-04 | 6.3E-04 | 3.6E-05 | 1.9E-04 | 1.5E-04 | 6.0E-05 | 1.9E-05 |
| 14,000 | 9.7E-05 | 5.8E-04 | 2.8E-05 | 1.7E-04 | 1.4E-04 | 5.6E-05 | 1.4E-05 |
| 15,000 | 7.4E-05 | 5.3E-04 | 2.2E-05 | 1.5E-04 | 1.3E-04 | 5.1E-05 | 1.1E-05 |
| 16,000 | 5.7E-05 | 4.8E-04 | 1.7E-05 | 1.4E-04 | 1.2E-04 | 4.8E-05 | 9.2E-06 |
| 17,000 | 4.4E-05 | 4.5E-04 | 1.4E-05 | 1.3E-04 | 1.2E-04 | 4.7E-05 | 7.3E-06 |
| 18,000 | 3.4E-05 | 4.2E-04 | 1.1E-05 | 1.2E-04 | 1.1E-04 | 4.2E-05 | 5.8E-06 |
| 19,000 | 2.7E-05 | 3.8E-04 | 8.7E-06 | 1.1E-04 | 1.1E-04 | 4.2E-05 | 4.6E-06 |
| 20,000 | 2.1E-05 | 3.6E-04 | 6.9E-06 | 1.0E-04 | 1.0E-04 | 3.8E-05 | 3.8E-06 |

**Table E5a.** Normalized Time and Space Integrated Particle Air Concentration for a Disc with the specified distance from the release assuming airborne loss rate (*λ*_*decay*_ = 10 h^-1^).

| Distance<br>Downwind | Clear, cold<br>night with<br>light winds | Clear night<br>with<br>gentle<br>breeze | Overcast<br>with light<br>winds | Overcast<br>with<br>gentle<br>winds | Overcast<br>with<br>strong<br>winds | Clear day<br>with<br>gentle<br>winds | Clear, hot<br>day with<br>light winds |
| --- | --- | --- | --- | --- | --- | --- | --- |
| (m) | (s m<sup>-1</sup>) | (s m<sup>-1</sup>) | (s m<sup>-1</sup>) | (s m<sup>-1</sup>) | (s m<sup>-1</sup>) | (s m<sup>-1</sup>) | (s m<sup>-1</sup>) |
| 50 | 1.7E+01 | 4.3E+00 | 9.7E+00 | 3.3E+00 | 1.5E+00 | 2.1E+00 | 6.5E+00 |
| 100 | 2.8E+01 | 7.5E+00 | 1.4E+01 | 5.4E+00 | 2.5E+00 | 2.9E+00 | 8.5E+00 |
| 150 | 3.6E+01 | 1.0E+01 | 1.6E+01 | 6.8E+00 | 3.2E+00 | 3.4E+00 | 9.4E+00 |
| 200 | 4.1E+01 | 1.2E+01 | 1.7E+01 | 7.8E+00 | 3.7E+00 | 3.7E+00 | 1.0E+01 |
| 250 | 4.4E+01 | 1.4E+01 | 1.8E+01 | 8.6E+00 | 4.1E+00 | 4.0E+00 | 1.0E+01 |
| 300 | 4.7E+01 | 1.5E+01 | 1.8E+01 | 9.2E+00 | 4.4E+00 | 4.2E+00 | 1.1E+01 |
| 350 | 4.9E+01 | 1.6E+01 | 1.9E+01 | 9.8E+00 | 4.7E+00 | 4.3E+00 | 1.1E+01 |
| 400 | 5.1E+01 | 1.7E+01 | 1.9E+01 | 1.0E+01 | 5.0E+00 | 4.4E+00 | 1.1E+01 |
| 450 | 5.2E+01 | 1.8E+01 | 2.0E+01 | 1.1E+01 | 5.2E+00 | 4.6E+00 | 1.1E+01 |
| 500 | 5.3E+01 | 1.8E+01 | 2.0E+01 | 1.1E+01 | 5.4E+00 | 4.6E+00 | 1.1E+01 |
| 550 | 5.3E+01 | 1.9E+01 | 2.0E+01 | 1.1E+01 | 5.5E+00 | 4.7E+00 | 1.1E+01 |
| 600 | 5.4E+01 | 2.0E+01 | 2.0E+01 | 1.2E+01 | 5.7E+00 | 4.8E+00 | 1.1E+01 |
| 650 | 5.4E+01 | 2.0E+01 | 2.0E+01 | 1.2E+01 | 5.8E+00 | 4.8E+00 | 1.1E+01 |
| 700 | 5.5E+01 | 2.1E+01 | 2.0E+01 | 1.2E+01 | 6.0E+00 | 4.9E+00 | 1.1E+01 |
| 750 | 5.5E+01 | 2.1E+01 | 2.0E+01 | 1.2E+01 | 6.1E+00 | 4.9E+00 | 1.1E+01 |
| 800 | 5.5E+01 | 2.1E+01 | 2.0E+01 | 1.2E+01 | 6.2E+00 | 5.0E+00 | 1.1E+01 |
| 850 | 5.5E+01 | 2.2E+01 | 2.0E+01 | 1.3E+01 | 6.3E+00 | 5.0E+00 | 1.1E+01 |
| 900 | 5.5E+01 | 2.2E+01 | 2.0E+01 | 1.3E+01 | 6.4E+00 | 5.1E+00 | 1.1E+01 |
| 950 | 5.5E+01 | 2.2E+01 | 2.0E+01 | 1.3E+01 | 6.5E+00 | 5.1E+00 | 1.1E+01 |
| 1,000 | 5.5E+01 | 2.3E+01 | 2.0E+01 | 1.3E+01 | 6.6E+00 | 5.1E+00 | 1.1E+01 |
| 1,050 | 5.6E+01 | 2.3E+01 | 2.0E+01 | 1.3E+01 | 6.7E+00 | 5.2E+00 | 1.1E+01 |
| 1,100 | 5.6E+01 | 2.3E+01 | 2.1E+01 | 1.3E+01 | 6.8E+00 | 5.2E+00 | 1.1E+01 |
| 2,000 | 6.1E+01 | 2.5E+01 | 2.1E+01 | 1.3E+01 | 7.0E+00 | 5.6E+00 | 1.2E+01 |
| 3,000 | 6.1E+01 | 2.5E+01 | 2.1E+01 | 1.4E+01 | 7.3E+00 | 5.7E+00 | 1.2E+01 |
| 4,000 | 6.1E+01 | 2.6E+01 | 2.1E+01 | 1.4E+01 | 7.4E+00 | 5.7E+00 | 1.2E+01 |
| 5,000 | 6.1E+01 | 2.6E+01 | 2.1E+01 | 1.4E+01 | 7.6E+00 | 5.8E+00 | 1.2E+01 |
| 6,000 | 6.1E+01 | 2.6E+01 | 2.1E+01 | 1.4E+01 | 7.6E+00 | 5.8E+00 | 1.2E+01 |
| 7,000 | 6.1E+01 | 2.6E+01 | 2.1E+01 | 1.4E+01 | 7.7E+00 | 5.8E+00 | 1.2E+01 |
| 8,000 | 6.1E+01 | 2.6E+01 | 2.1E+01 | 1.4E+01 | 7.7E+00 | 5.8E+00 | 1.2E+01 |
| 9,000 | 6.1E+01 | 2.6E+01 | 2.1E+01 | 1.4E+01 | 7.8E+00 | 5.8E+00 | 1.2E+01 |
| 10,000 | 6.1E+01 | 2.6E+01 | 2.1E+01 | 1.4E+01 | 7.8E+00 | 5.8E+00 | 1.2E+01 |
| 11,000 | 6.1E+01 | 2.6E+01 | 2.1E+01 | 1.4E+01 | 7.8E+00 | 5.8E+00 | 1.2E+01 |
| 12,000 | 6.1E+01 | 2.6E+01 | 2.1E+01 | 1.4E+01 | 7.8E+00 | 5.8E+00 | 1.2E+01 |
| 13,000 | 6.1E+01 | 2.6E+01 | 2.1E+01 | 1.4E+01 | 7.8E+00 | 5.8E+00 | 1.2E+01 |
| 14,000 | 6.1E+01 | 2.6E+01 | 2.1E+01 | 1.4E+01 | 7.8E+00 | 5.8E+00 | 1.2E+01 |
| 15,000 | 6.1E+01 | 2.6E+01 | 2.1E+01 | 1.4E+01 | 7.8E+00 | 5.8E+00 | 1.2E+01 |
| 16,000 | 6.1E+01 | 2.6E+01 | 2.1E+01 | 1.4E+01 | 7.8E+00 | 5.8E+00 | 1.2E+01 |
| 17,000 | 6.1E+01 | 2.6E+01 | 2.1E+01 | 1.4E+01 | 7.9E+00 | 5.8E+00 | 1.2E+01 |
| 18,000 | 6.1E+01 | 2.6E+01 | 2.1E+01 | 1.4E+01 | 7.9E+00 | 5.8E+00 | 1.2E+01 |
| 19,000 | 6.1E+01 | 2.6E+01 | 2.1E+01 | 1.4E+01 | 7.9E+00 | 5.8E+00 | 1.2E+01 |
| 20,000 | 6.1E+01 | 2.6E+01 | 2.1E+01 | 1.4E+01 | 7.9E+00 | 5.8E+00 | 1.2E+01 |

**Table E5b.** Normalized Time and Space Integrated Particle Air Concentration for a Circle Arc at the specified distance from the release assuming airborne loss rate (*λ*_*decay*_ = 10 h^-1^).

| Distance<br>Downwind<br>(m) | Clear, cold<br>night with<br>light winds<br>(s m<sup>-2</sup>) | Clear night<br>with<br>gentle<br>breeze<br>(s m<sup>-2</sup>) | Overcast<br>with light<br>winds<br>(s m<sup>-2</sup>) | Overcast<br>with<br>gentle<br>winds<br>(s m<sup>-2</sup>) | Overcast<br>with<br>strong<br>winds<br>(s m<sup>-2</sup>) | Clear day<br>with<br>gentle<br>winds<br>(s m<sup>-2</sup>) | Clear, hot<br>day with<br>light winds<br>(s m<sup>-2</sup>) |
| --- | --- | --- | --- | --- | --- | --- | --- |
| 50 | 2.7E-01 | 7.5E-02 | 1.1E-01 | 5.1E-02 | 2.4E-02 | 2.3E-02 | 5.7E-02 |
| 100 | 1.8E-01 | 5.6E-02 | 5.5E-02 | 3.3E-02 | 1.6E-02 | 1.2E-02 | 2.5E-02 |
| 150 | 1.2E-01 | 4.4E-02 | 3.3E-02 | 2.4E-02 | 1.2E-02 | 7.9E-03 | 1.4E-02 |
| 200 | 8.6E-02 | 3.5E-02 | 2.1E-02 | 1.8E-02 | 9.0E-03 | 5.6E-03 | 9.0E-03 |
| 250 | 6.3E-02 | 2.9E-02 | 1.5E-02 | 1.4E-02 | 7.4E-03 | 4.2E-03 | 6.2E-03 |
| 300 | 4.6E-02 | 2.4E-02 | 1.1E-02 | 1.2E-02 | 6.2E-03 | 3.4E-03 | 4.4E-03 |
| 350 | 3.4E-02 | 2.1E-02 | 7.9E-03 | 9.8E-03 | 5.3E-03 | 2.8E-03 | 3.2E-03 |
| 400 | 2.6E-02 | 1.8E-02 | 6.0E-03 | 8.4E-03 | 4.7E-03 | 2.3E-03 | 2.4E-03 |
| 450 | 2.0E-02 | 1.6E-02 | 4.6E-03 | 7.3E-03 | 4.1E-03 | 2.0E-03 | 1.9E-03 |
| 500 | 1.5E-02 | 1.4E-02 | 3.6E-03 | 6.4E-03 | 3.7E-03 | 1.7E-03 | 1.5E-03 |
| 550 | 1.2E-02 | 1.2E-02 | 2.9E-03 | 5.6E-03 | 3.3E-03 | 1.5E-03 | 1.2E-03 |
| 600 | 9.3E-03 | 1.1E-02 | 2.3E-03 | 5.0E-03 | 3.0E-03 | 1.3E-03 | 9.4E-04 |
| 650 | 7.3E-03 | 9.6E-03 | 1.9E-03 | 4.5E-03 | 2.8E-03 | 1.1E-03 | 7.5E-04 |
| 700 | 5.7E-03 | 8.6E-03 | 1.5E-03 | 4.1E-03 | 2.5E-03 | 1.0E-03 | 6.1E-04 |
| 750 | 4.6E-03 | 7.8E-03 | 1.3E-03 | 3.7E-03 | 2.4E-03 | 9.0E-04 | 5.1E-04 |
| 800 | 3.6E-03 | 7.0E-03 | 1.0E-03 | 3.4E-03 | 2.2E-03 | 8.1E-04 | 4.3E-04 |
| 850 | 2.9E-03 | 6.5E-03 | 8.6E-04 | 3.1E-03 | 2.1E-03 | 7.4E-04 | 3.6E-04 |
| 900 | 2.3E-03 | 5.9E-03 | 7.2E-04 | 2.8E-03 | 1.9E-03 | 6.7E-04 | 3.0E-04 |
| 950 | 1.8E-03 | 5.4E-03 | 6.0E-04 | 2.6E-03 | 1.8E-03 | 6.0E-04 | 2.5E-04 |
| 1,000 | 1.5E-03 | 5.0E-03 | 5.0E-04 | 2.4E-03 | 1.7E-03 | 5.6E-04 | 2.1E-04 |
| 1,050 | 1.2E-03 | 4.6E-03 | 4.2E-04 | 2.2E-03 | 1.6E-03 | 5.2E-04 | 1.8E-04 |
| 1,100 | 9.2E-04 | 4.2E-03 | 3.4E-04 | 2.0E-03 | 1.5E-03 | 4.7E-04 | 1.5E-04 |
| 2,000 | 4.9E-05 | 1.1E-03 | 2.3E-05 | 4.1E-04 | 4.0E-04 | 1.4E-04 | 1.2E-05 |
| 3,000 | 2.6E-06 | 4.1E-04 | 2.1E-06 | 1.7E-04 | 2.2E-04 | 5.5E-05 | 1.0E-06 |
| 4,000 | 2.0E-07 | 1.9E-04 | 2.5E-07 | 7.6E-05 | 1.4E-04 | 2.6E-05 | 1.1E-07 |
| 5,000 | 2.1E-08 | 9.7E-05 | 3.3E-08 | 3.9E-05 | 9.3E-05 | 1.4E-05 | 1.4E-08 |
| 6,000 | 2.2E-09 | 5.2E-05 | 4.4E-09 | 2.1E-05 | 6.6E-05 | 7.4E-06 | 1.7E-09 |
| 7,000 | 2.2E-10 | 2.9E-05 | 6.0E-10 | 1.1E-05 | 4.7E-05 | 4.2E-06 | 1.9E-10 |
| 8,000 | 1.3E-11 | 1.7E-05 | 6.4E-11 | 6.5E-06 | 3.5E-05 | 2.4E-06 | 1.7E-11 |
| 9,000 | 3.7E-13 | 1.0E-05 | 3.2E-12 | 3.8E-06 | 2.7E-05 | 1.4E-06 | 5.5E-13 |
| 10,000 | 0.0E+00 | 6.3E-06 | 9.3E-14 | 2.2E-06 | 2.0E-05 | 8.5E-07 | 0.0E+00 |
| 11,000 | 0.0E+00 | 3.9E-06 | 0.0E+00 | 1.3E-06 | 1.6E-05 | 5.3E-07 | 0.0E+00 |
| 12,000 | 0.0E+00 | 2.5E-06 | 0.0E+00 | 8.1E-07 | 1.2E-05 | 3.1E-07 | 0.0E+00 |
| 13,000 | 0.0E+00 | 1.6E-06 | 0.0E+00 | 5.0E-07 | 9.6E-06 | 1.9E-07 | 0.0E+00 |
| 14,000 | 0.0E+00 | 1.0E-06 | 0.0E+00 | 3.0E-07 | 7.6E-06 | 1.1E-07 | 0.0E+00 |
| 15,000 | 0.0E+00 | 6.8E-07 | 0.0E+00 | 1.8E-07 | 5.8E-06 | 6.8E-08 | 0.0E+00 |
| 16,000 | 0.0E+00 | 4.4E-07 | 0.0E+00 | 1.1E-07 | 4.6E-06 | 4.3E-08 | 0.0E+00 |
| 17,000 | 0.0E+00 | 3.0E-07 | 0.0E+00 | 6.9E-08 | 3.7E-06 | 2.7E-08 | 0.0E+00 |
| 18,000 | 0.0E+00 | 1.9E-07 | 0.0E+00 | 4.2E-08 | 3.0E-06 | 1.6E-08 | 0.0E+00 |
| 19,000 | 0.0E+00 | 1.3E-07 | 0.0E+00 | 2.6E-08 | 2.3E-06 | 1.0E-08 | 0.0E+00 |
| 20,000 | 0.0E+00 | 8.7E-08 | 0.0E+00 | 1.7E-08 | 1.9E-06 | 6.2E-09 | 0.0E+00 |

**Figure E1.**
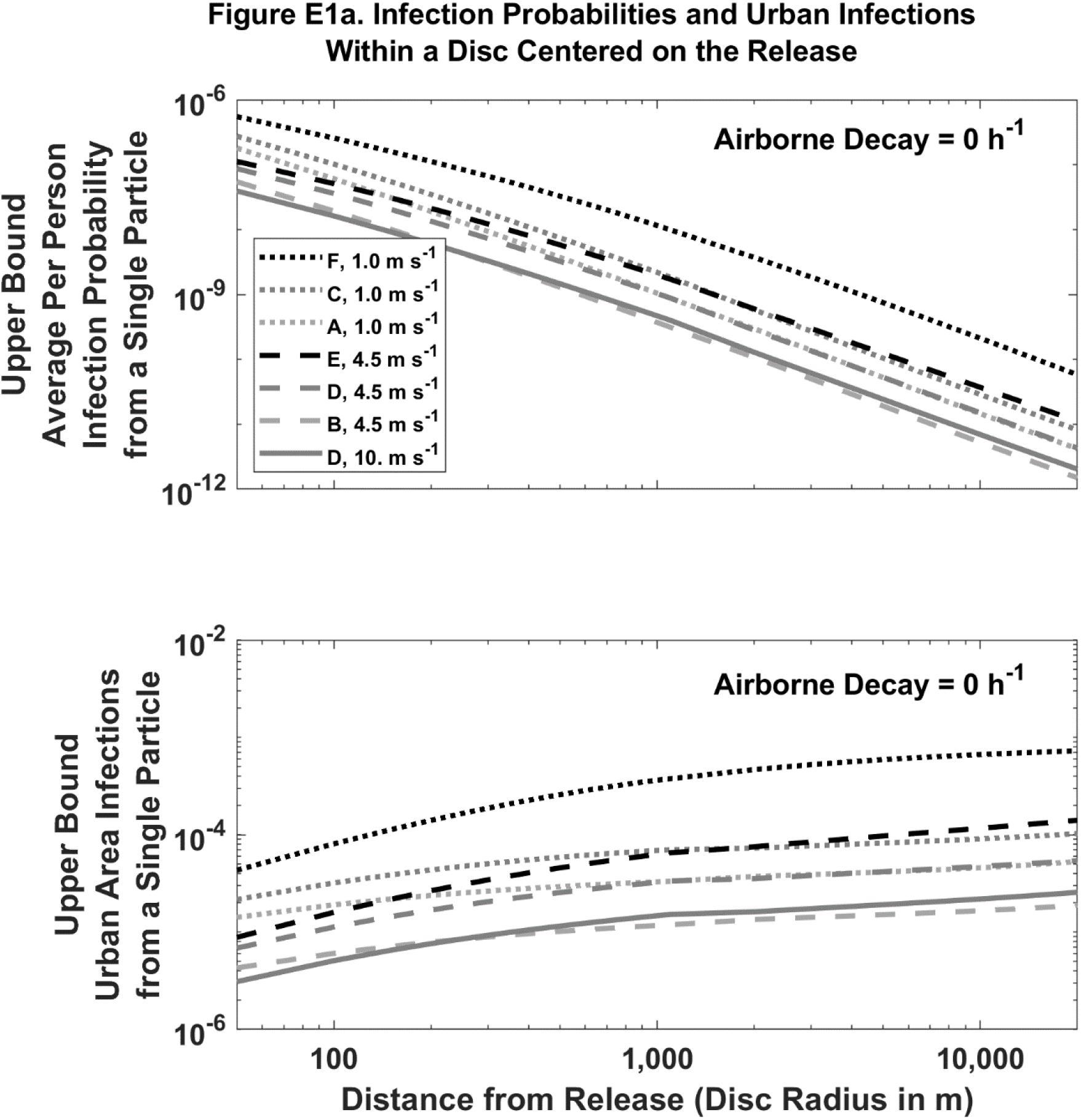
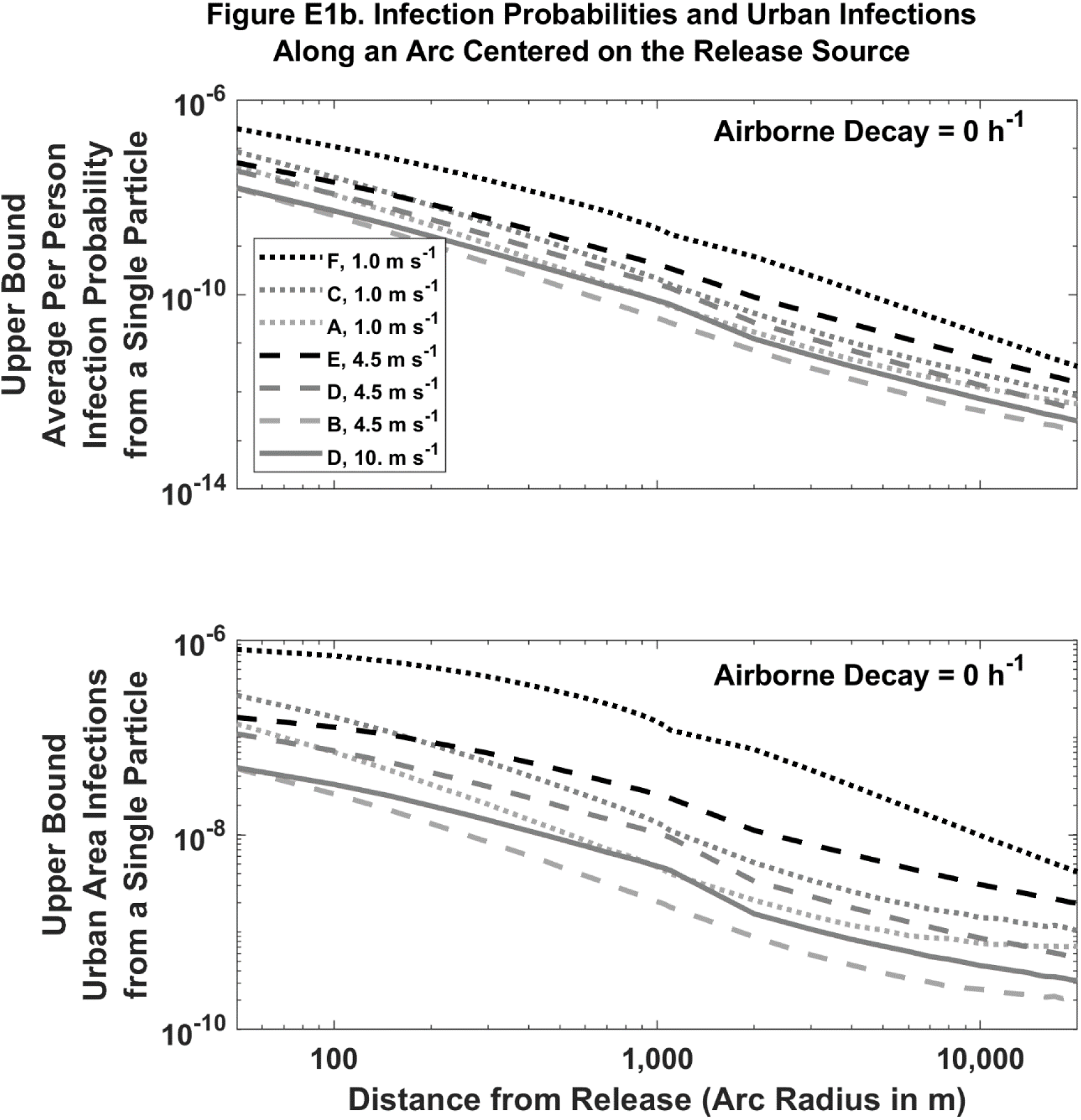
Predicted absolute infection probabilities and infections by distance, wind speed and atmospheric stability for a single airborne particle with 0 h^-1^ airborne loss of infectivity. Legend indicates Pasquill-Gifford-Turner atmospheric stability class (A to F) and the 10 m agl wind speed. Individual person infection probability (top panels) is dimensionless. Urban area infections (bottom panels) assumes a uniform population density of 0.01 people m^-2^ and has dimensions of people (disc) or people m^-1^ (arc).

**Figure E2.**
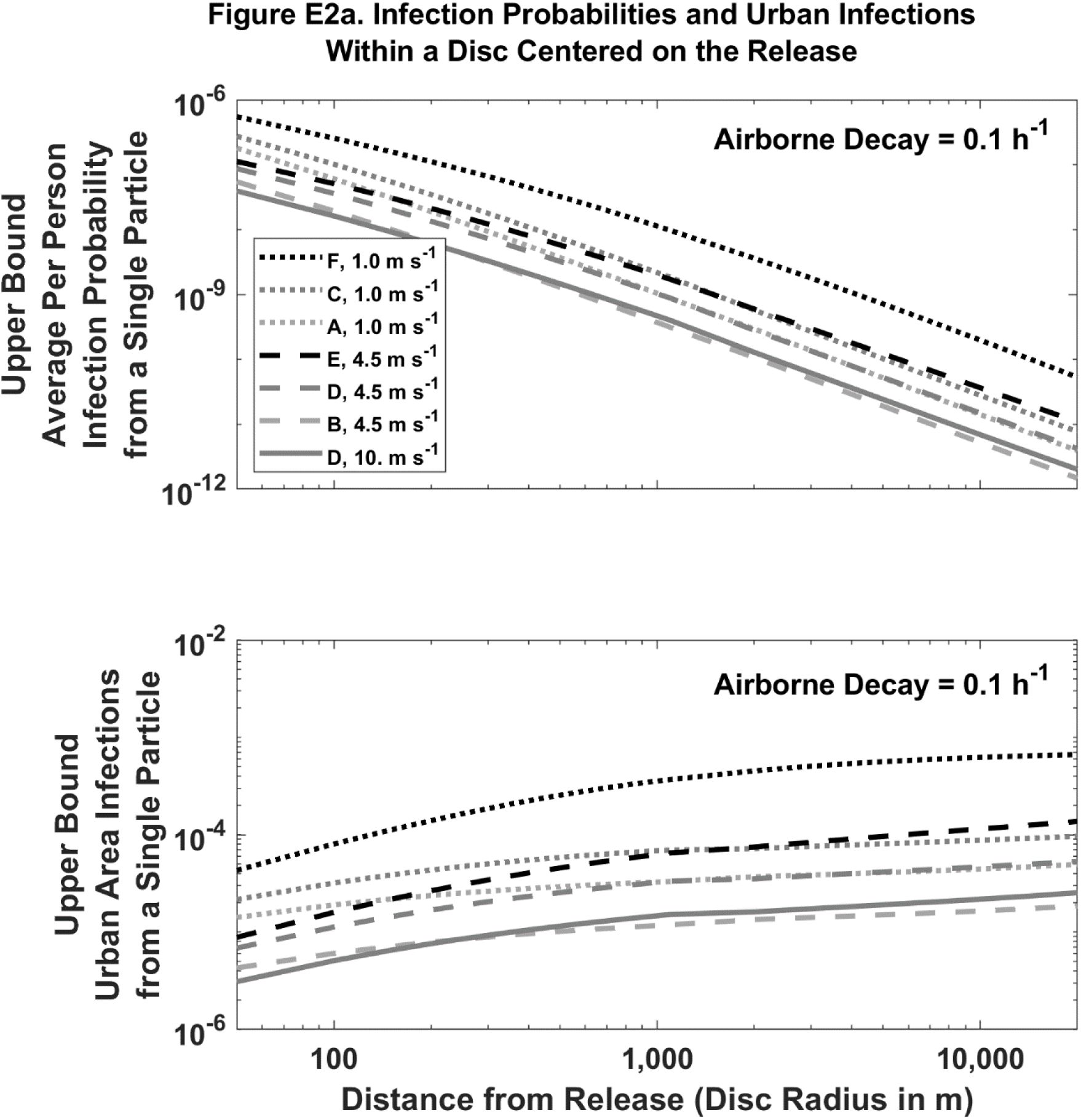
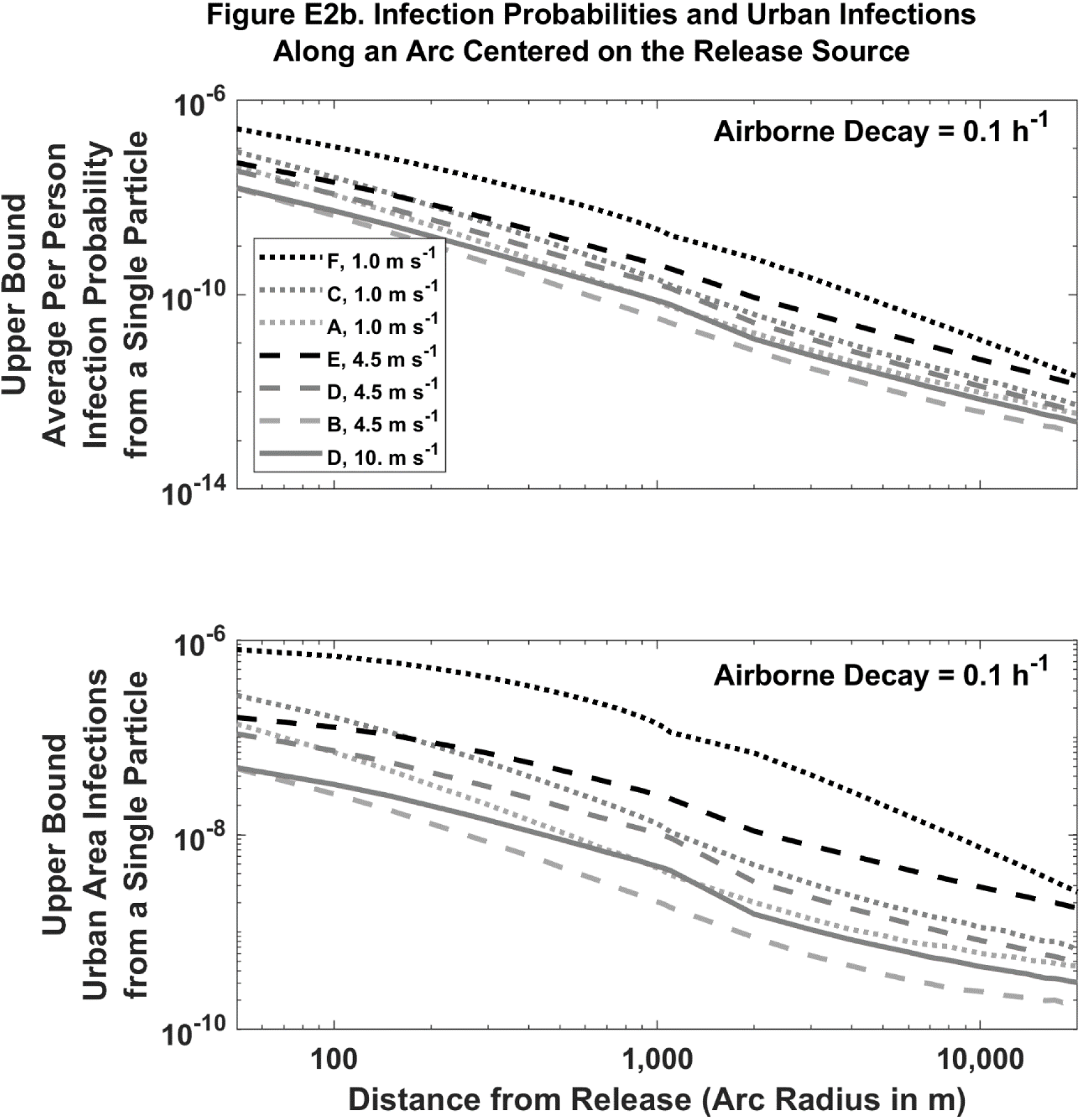
Predicted absolute infection probabilities and infections by distance, wind speed and atmospheric stability for a single airborne particle with 0.1 h^-1^ airborne loss of infectivity. Legend indicates Pasquill-Gifford-Turner atmospheric stability class (A to F) and the 10 m agl wind speed. Individual person infection probability (top panels) is dimensionless. Urban area infections (bottom panels) assumes a uniform population density of 0.01 people m^-2^ and has dimensions of people (disc) or people m^-1^ (arc).

**Figure E3.**
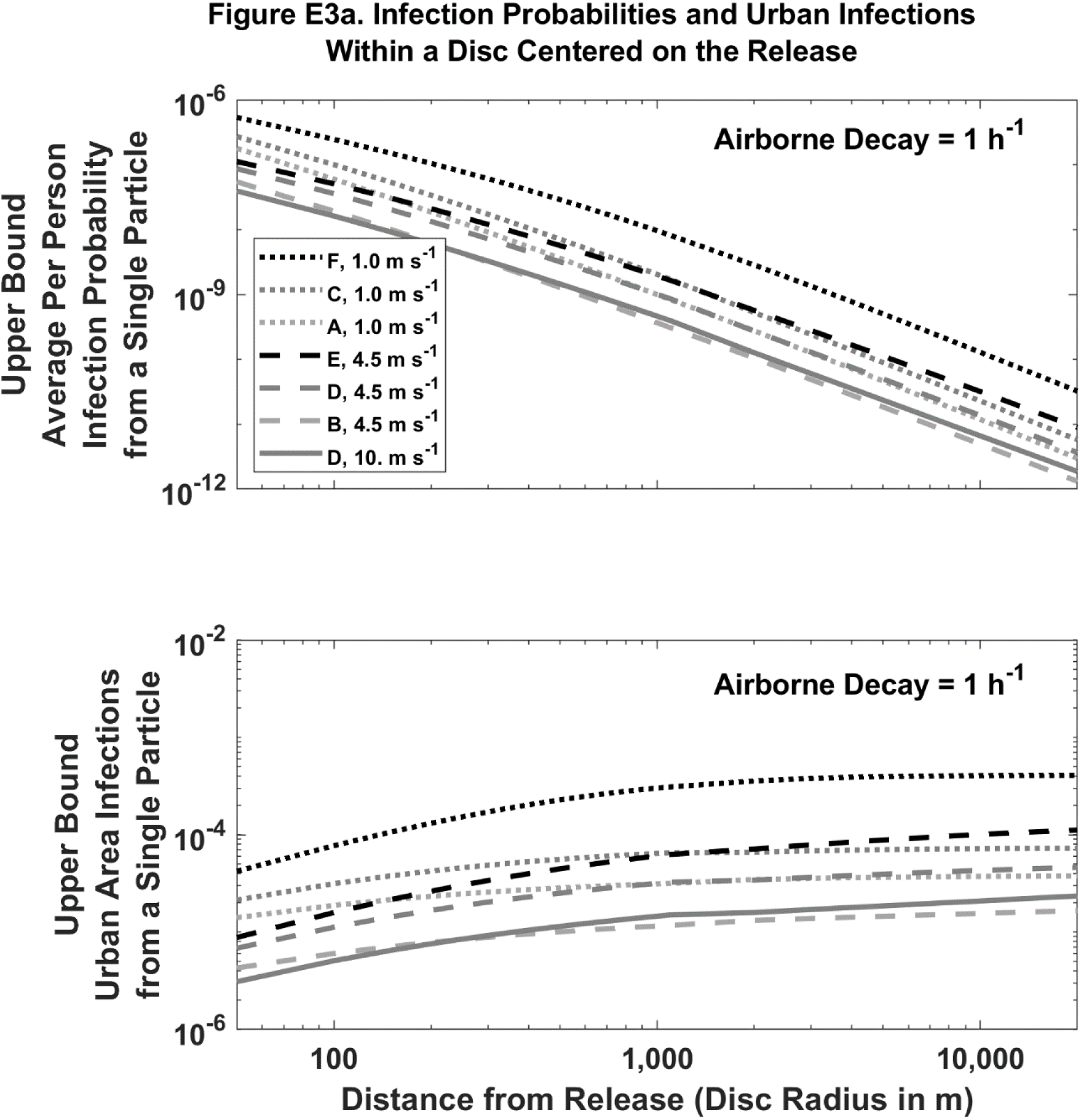
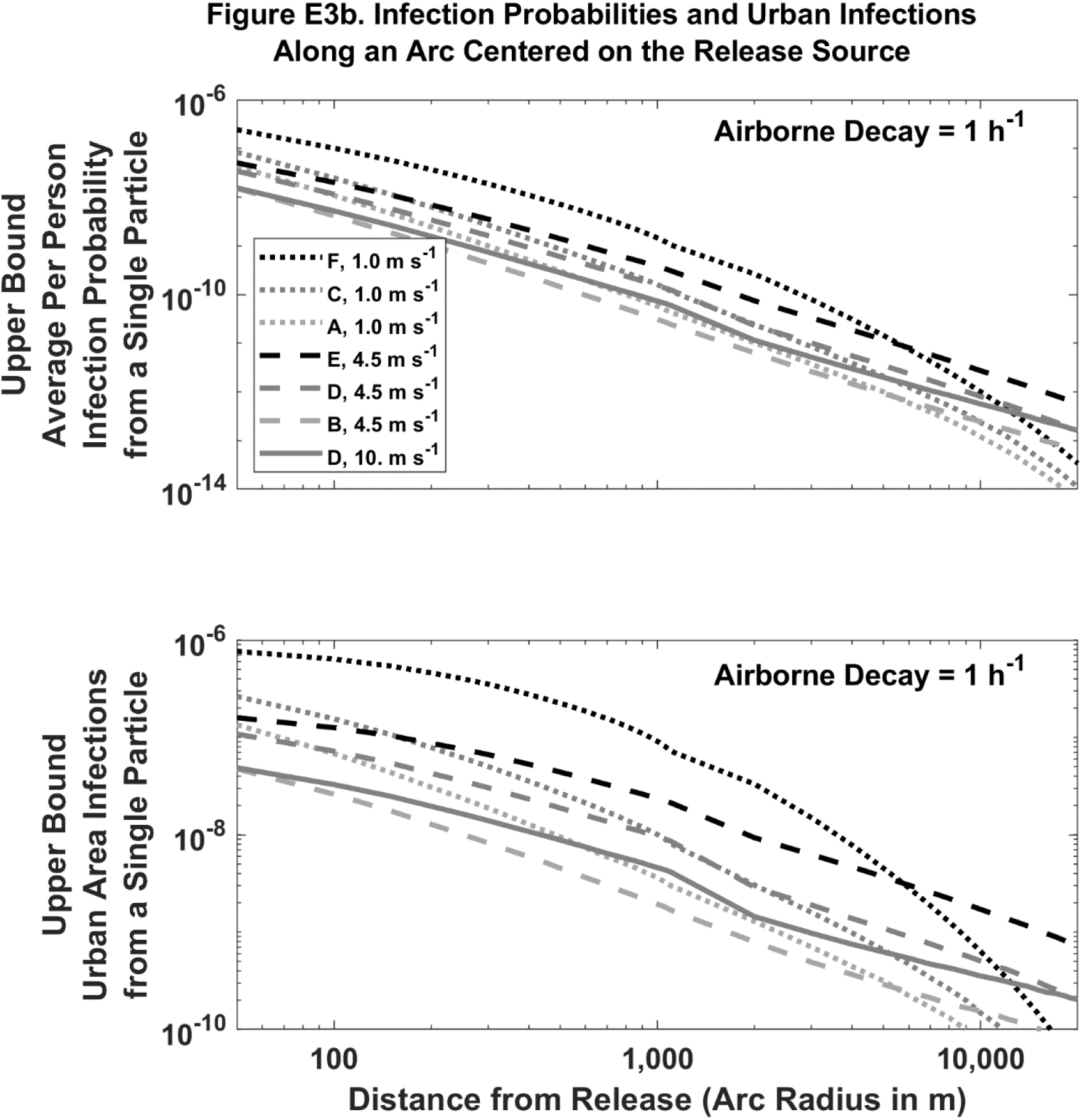
Predicted absolute infection probabilities and infections by distance, wind speed and atmospheric stability for a single airborne particle with 1 h^-1^ airborne loss of infectivity. Legend indicates Pasquill-Gifford-Turner atmospheric stability class (A to F) and the 10 m agl wind speed. Individual person infection probability (top panels) is dimensionless. Urban area infections (bottom panels) assumes a uniform population density of 0.01 people m^-2^ and has dimensions of people (disc) or people m^-1^ (arc).

**Figure E4.**
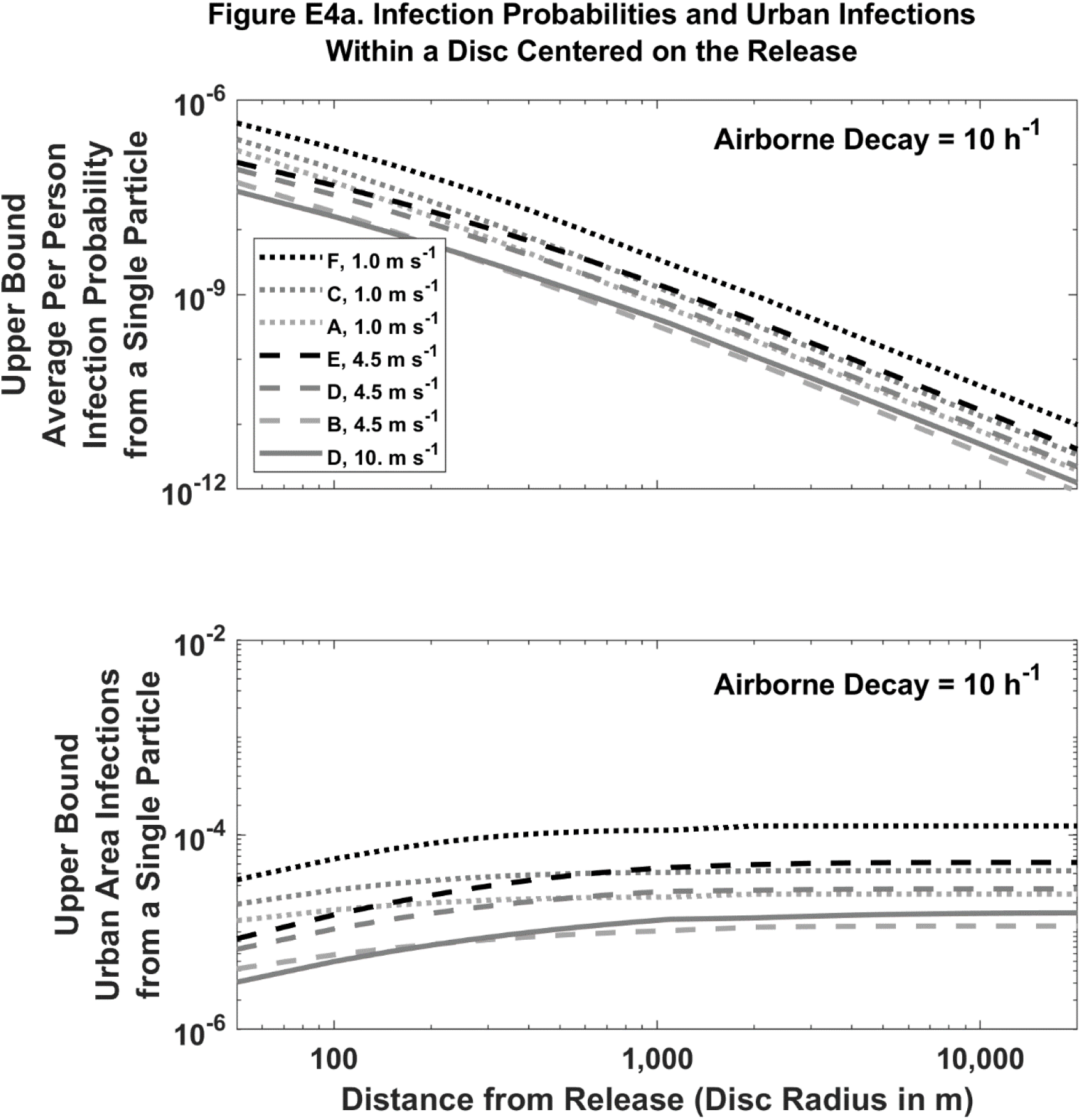
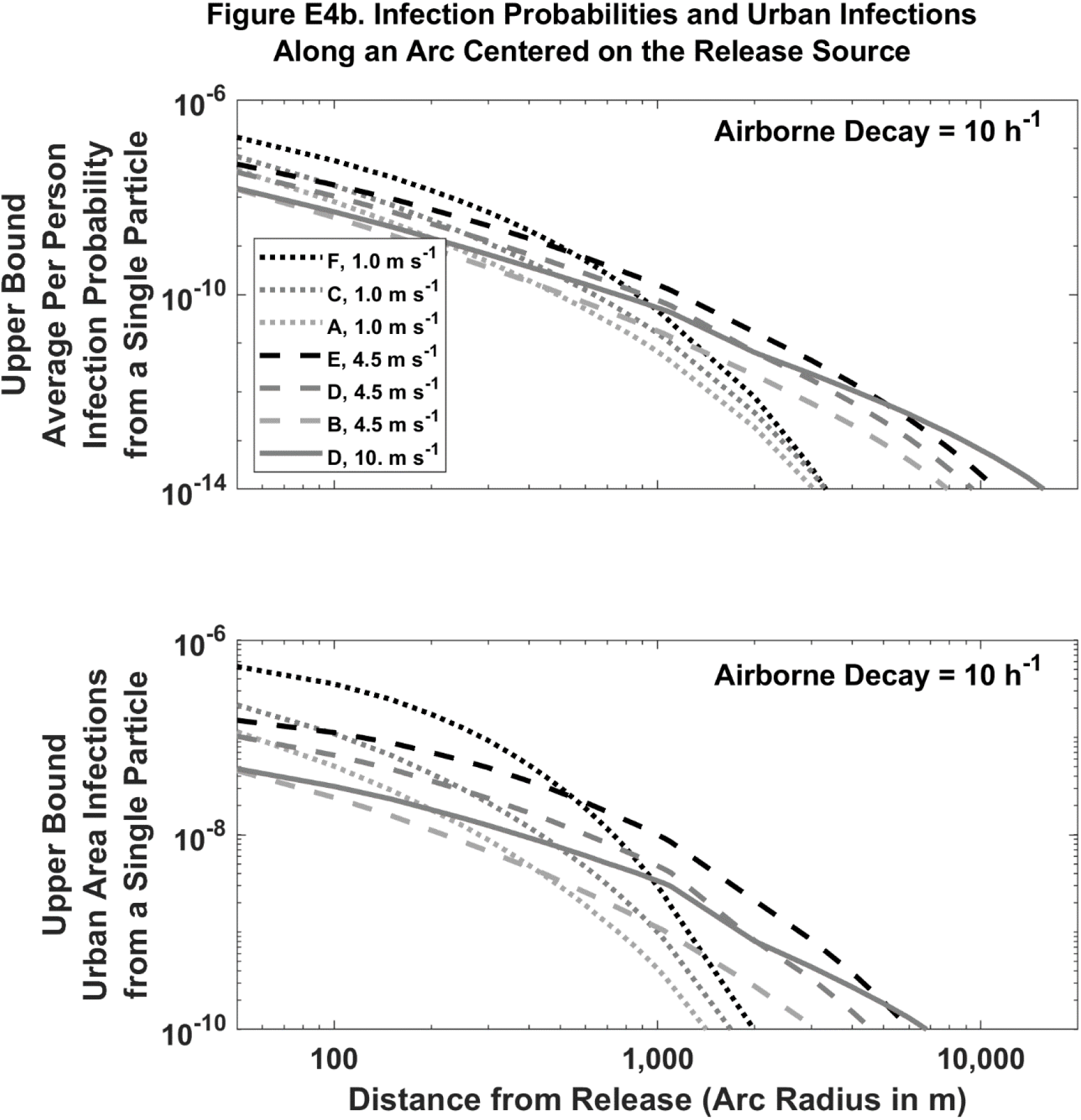
Predicted absolute infection probabilities and infections by distance, wind speed and atmospheric stability for a single airborne particle with 10 h^-1^ airborne loss of infectivity. Legend indicates Pasquill-Gifford-Turner atmospheric stability class (A to F) and the 10 m agl wind speed. Individual person infection probability (top panels) is dimensionless. Urban area infections (bottom panels) a uniform population density of 0.01 people m^-2^ and has dimensions of people (disc) or people m^-1^ (arc).

**Figure E5.**
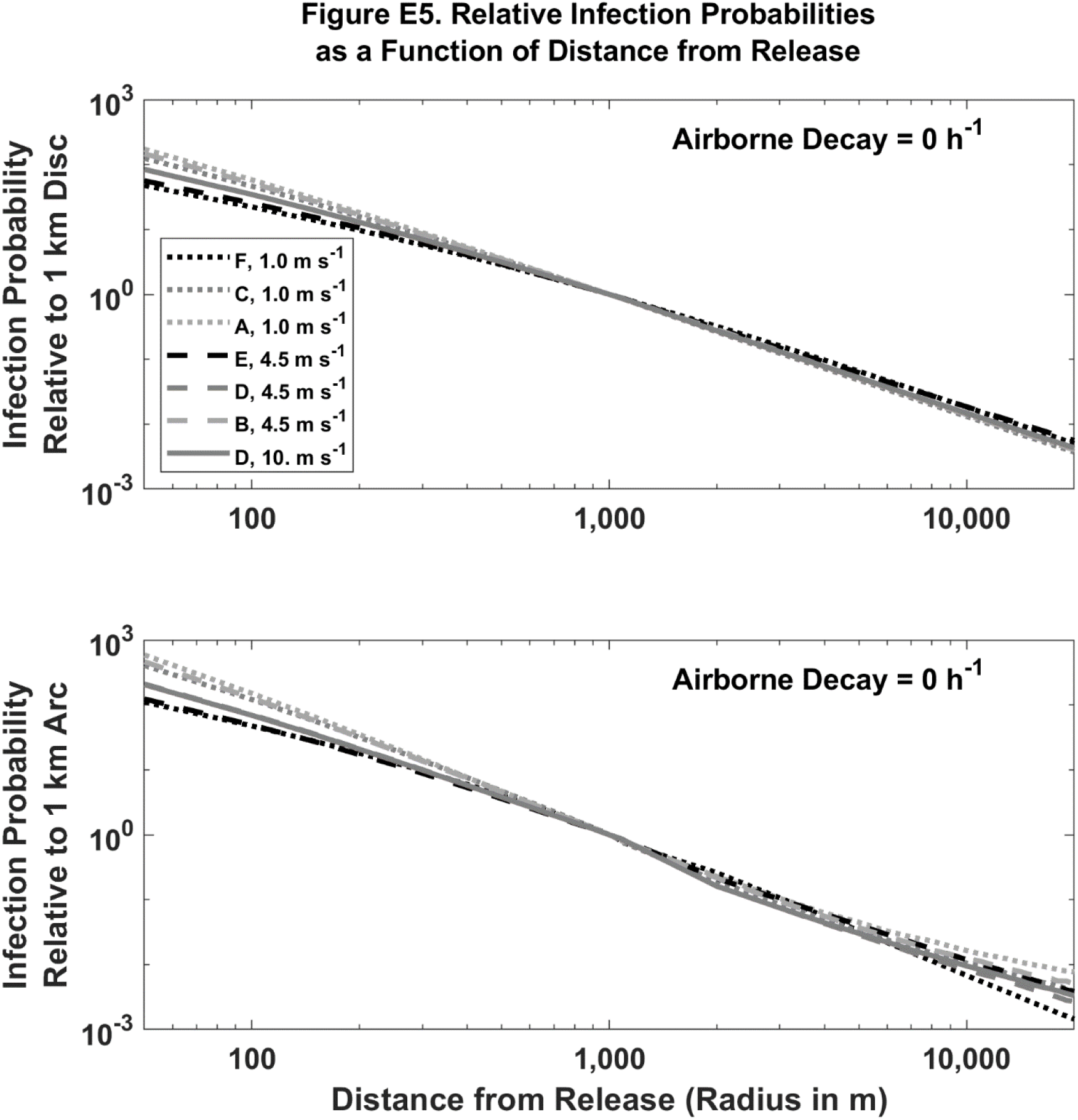
Predicted relative infection probabilities by distance, wind speed and atmospheric stability for a single airborne particle with 0 h^-1^ airborne loss of infectivity. Legend indicates Pasquill-Gifford-Turner atmospheric stability class (A to F) and the 10 m agl wind speed. Relative infection probability is dimensionless.

**Figure E6.**
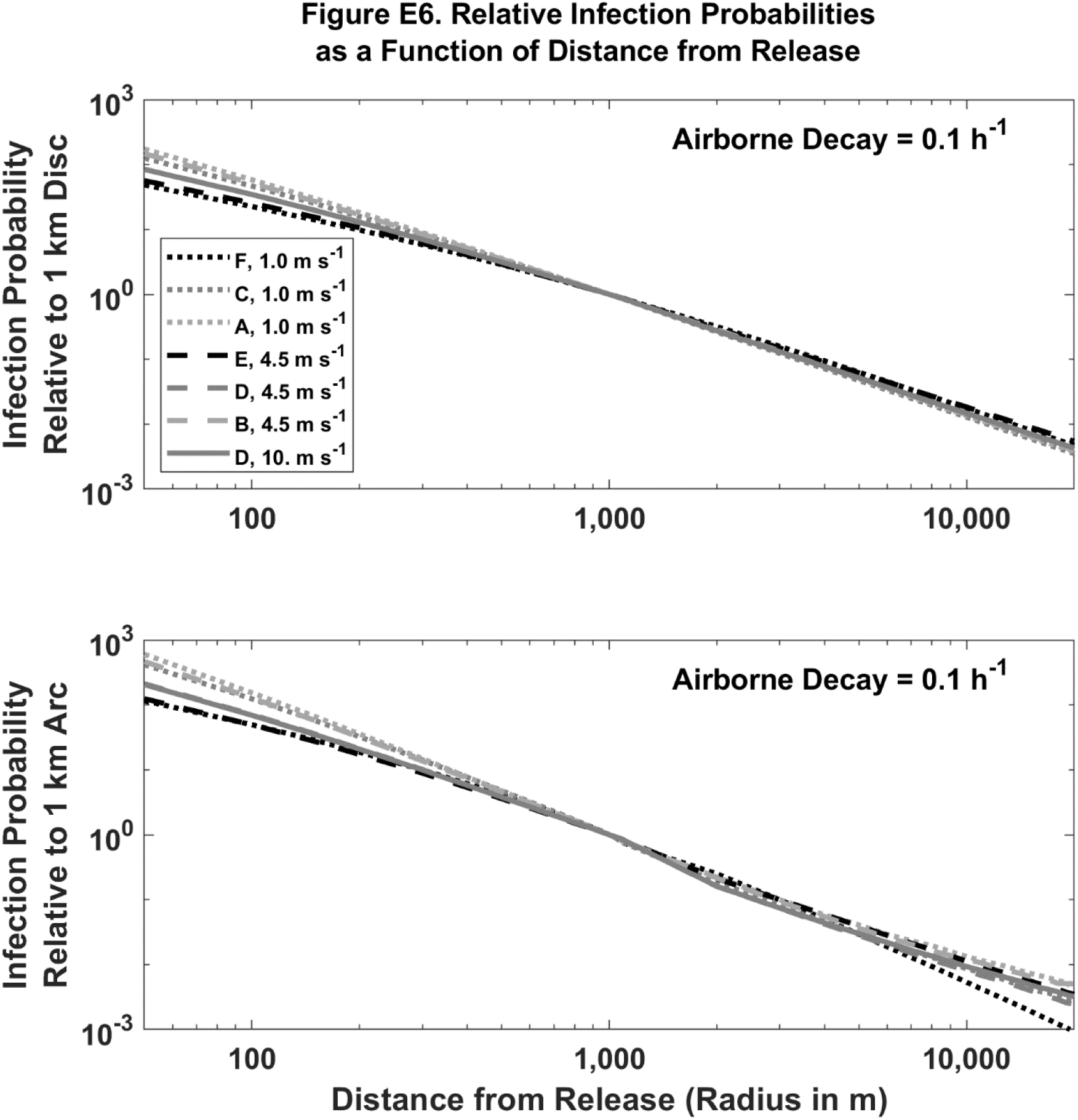
Predicted relative infection probabilities by distance, wind speed and atmospheric stability for a single airborne particle with 0.1 h^-1^ airborne loss of infectivity. Legend indicates Pasquill-Gifford-Turner atmospheric stability class (A to F) and the 10 m agl wind speed. Relative infection probability is dimensionless.

**Figure E7.**
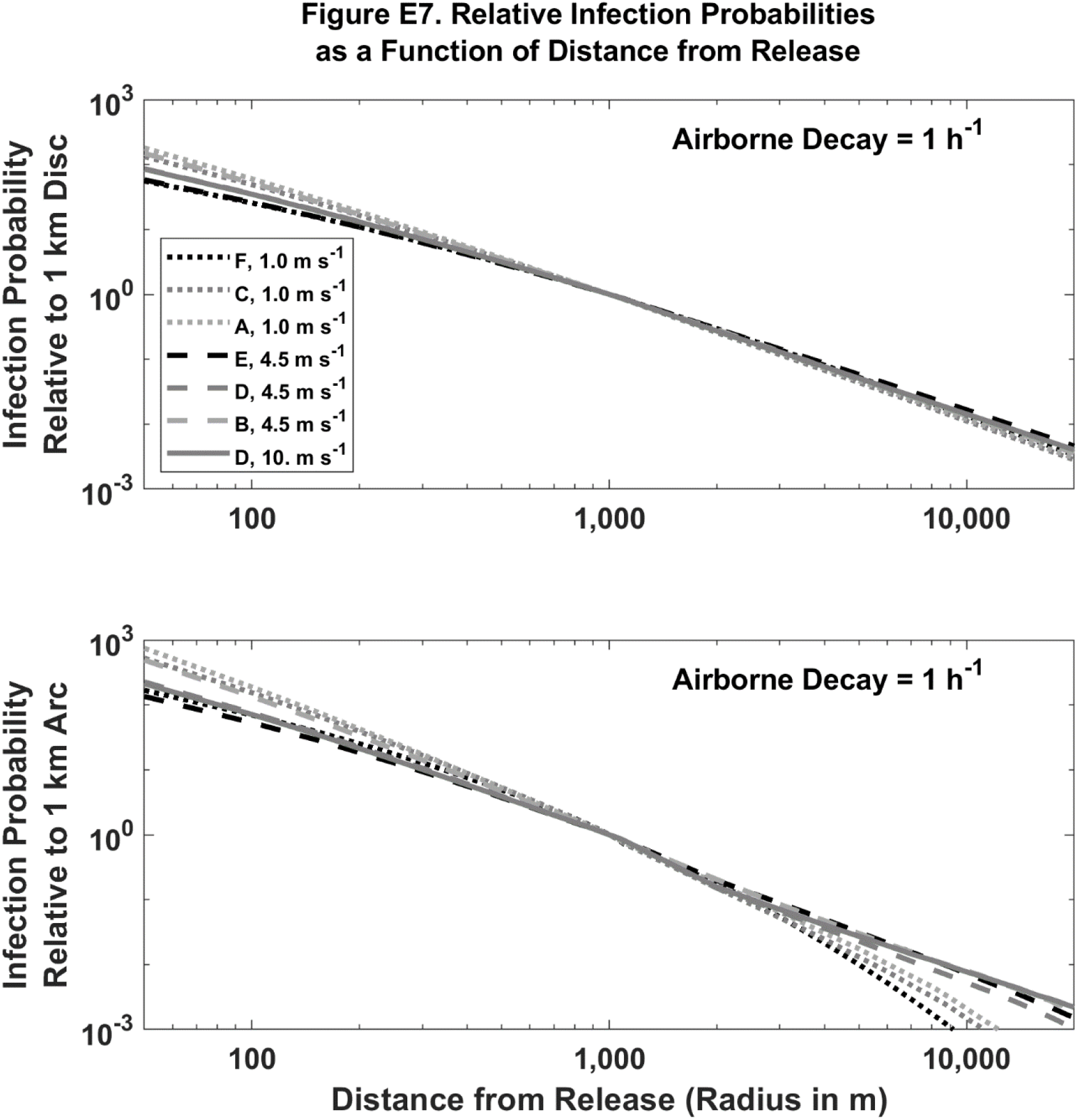
Predicted relative infection probabilities by distance, wind speed and atmospheric stability for a single airborne particle with 1 h^-1^ airborne loss of infectivity. Legend indicates Pasquill-Gifford-Turner atmospheric stability class (A to F) and the 10 m agl wind speed. Relative infection probability is dimensionless.

**Figure E8.**
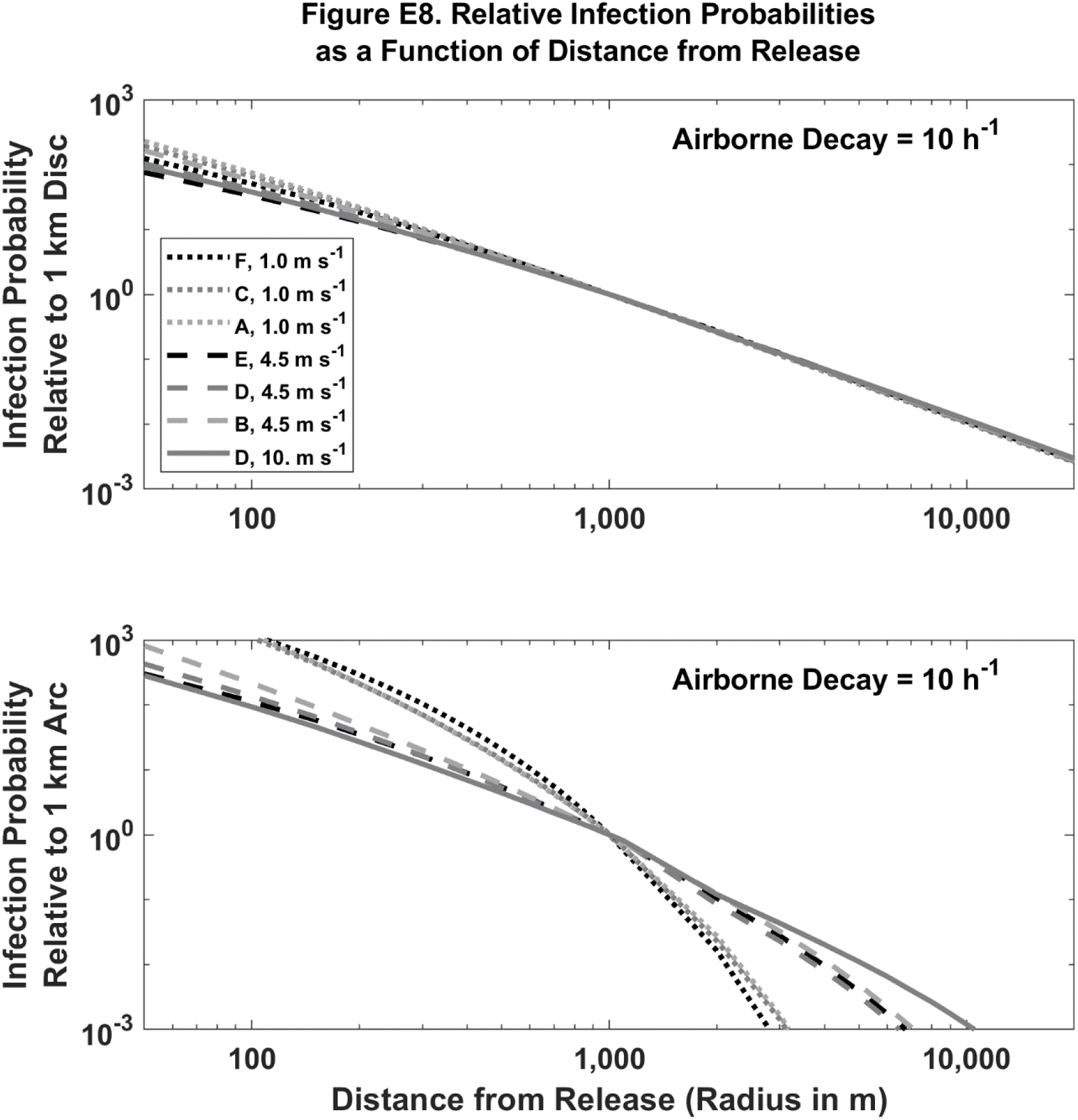
Predicted relative infection probabilities by distance, wind speed and atmospheric stability for a single airborne particle with 10 h^-1^ airborne loss of infectivity. Legend indicates Pasquill-Gifford-Turner atmospheric stability class (A to F) and the 10 m agl wind speed. Relative infection probability is dimensionless.

## Supplemental Material F: Outbreak Model-Measurement Comparison

see companion excel spreadsheet

## Supplemental Material G: Infection Estimates

### Section Variables Definitions

*b* = a specific particle type

*r* = a specific geographic region

[*Expected Number of Infections in Region*](*r, b*) =number of infections in region *r* resulting from exposure to *b*-type particles (people)

[*Population Density*](*r*) = population density in region *r* (people m^-2^)

[*Total Particles Refeased*] = total number of infectious, airborne particles released into the air (particles)

### Key Equation

**Equation G1** is adapted from **Equations 1** and **2** in the main text.

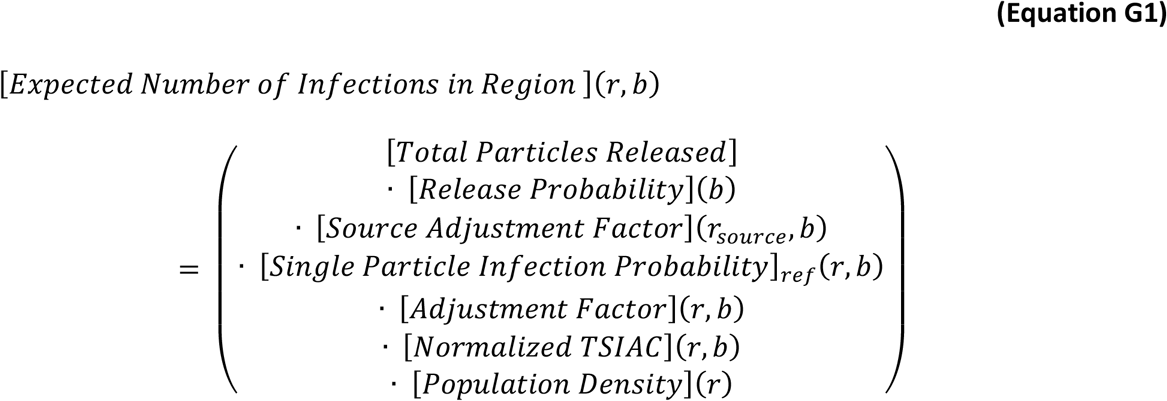

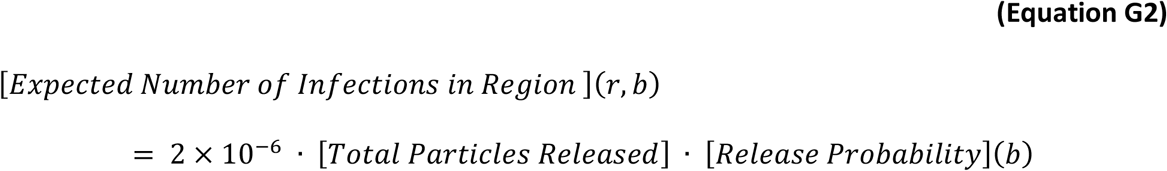

### Downwind Infections

*Assumptions*

1. There is an approximately 20% chance of particles emitted indoors exiting the house assuming the 1 μm diameter particles lose infectivity at a rate of 1 hr^-1^:

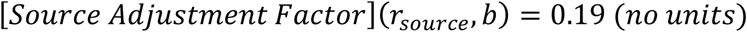
2. Each particle will cause infection if inhaled and each individual has a breathing rate of 10^−4^ m^3^ s^-1^:

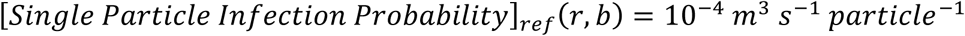
3. Downwind individuals are fully susceptible, but are physically protected to same degree as typical US person is protected from an airborne, outdoor, 1 μm diameter particle that losses infectivity at a rate of 1 hr^-1^:

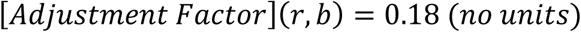
4. Exposures occur in an urban area:

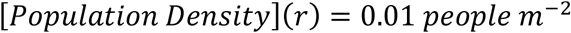
5. Downwind individuals are located between 50 m and 20 km from the infected person, the meteorology corresponds to clear night with a gentle breeze (stable atmospheric conditions and a 10 m agl wind speed of 4.5 m s^-1^), each particle has a 1 μm aerodynamic diameter and the particle loses infectivity in the atmosphere at a rate of 1 hr^-1^ (see **Table E4a**):

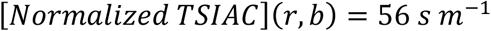

### Within Building Assumptions

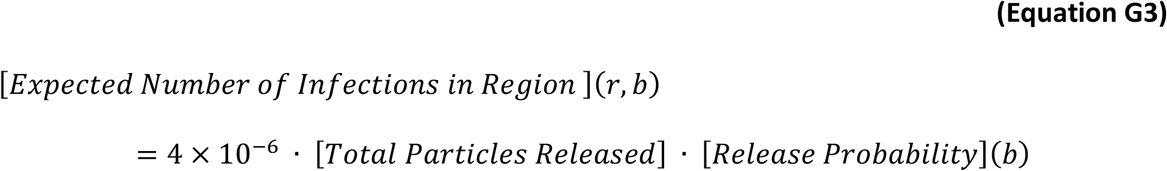

*Assumptions*

1. Airborne particles are released indoors:

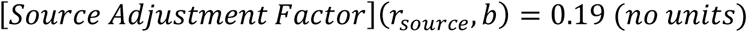
2. Each particle will cause infection if inhaled and each individual has a breathing rate of 10^−4^ m^3^ s^-1^:
3. [*Single Particle Infection Probability*]_*ref*_(*r, b*) = 10^−4^ *m*^3^ *s*^−1^ *particle*^−1^
4. Indoor individuals are fully susceptible and are not protected:

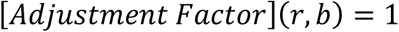
5. There is one susceptible individual in a typical US home. For context, a typical US single family home has 200 m^2^ of heated floor area and 2 total residents (Tables HC10.14 and HC9.1, respectively, per [244]):

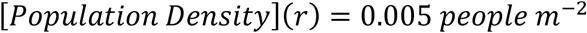
6. Exposures are determined by **Equation D7** using typical US home values [30] and an assumed room height of 3 m. Each particle has a 1 μm aerodynamic diameter and the particle loses infectivity at a rate of 1 hr^-1^.

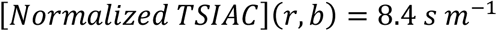

## Supplemental Material H: U.S. Coxiella burnetti Infection and Disease Estimates

Q Fever is a veterinary disease of livestock that can be transmitted to humans by inhalation of infectious aerosols [61] and has caused significant community and regional level disease outbreaks, see **Supplemental Material A: Airborne Disease Transmission Literature Review**. Control measures exist for animal and human Q Fever and vaccines are available in some countries [245]–[248].

### Background

In the 1930’s, it was first recognized that exposure to infected livestock could cause human disease. During a 1935 disease outbreak, Q Fever was recognized in Australia. Initially the cause was unknown and so investigators called it “Query” (of questionable cause) Fever. The bacterium Coxiella burnetti, which causes Q Fever, was named after the two researchers who first identified it (Herald Cox and McFarlane Burnet). Since this time, Coxiella burnetti infection in animals has been shown to be common in almost all countries, including the United States [249] and causes significant disease in commercial livestock including, but not limited to, cows, sheep, and goats [250]. A recently summary indicates high US infection rates in goats (41.6%), sheep (16.5%), and cattle (3.4%) [250].

In the US soils, an environmental survey demonstrates widespread contamination, with 23.8% of samples testing positive for Coxiella DNA [251]. US state-level positive sample rates range from 6 to 44%. A subset of these samples was tested for viability and Coxiella was culturable in the PCR positive specimens. As expected, the Coxiella DNA was detected in locations with livestock; however, it was also often found in locations associated with human activity, such as post offices, stores, and schools.

### Human Infection and Disease Characteristics

Q Fever in humans is characteristically caused by inhalation of airborne bacteria, although direct contact and ticks are also known infection pathways [61], [249]. The infectious aerosols may be emitted directly from infected animals or the result of aerosolization of contaminated particles or soil. Coxiella is highly infectious since exposure to only a few bacteria can cause infection^42^ and while there is some variation between research studies and also variation by infectious dosage, in general it is thought that 40% to 50% of humans infected for the first time become ill with Acute Q Fever. The proportion of infected persons who will develop Chronic Q Fever is still uncertain as it can be much more difficult to diagnose [61], [249], [252]–[254].

Acute Q Fever often exhibits as a short influenza-like illness, but the disease can be severe with pneumonia, liver disease, or encephalitis requiring hospitalization. Acute Q Fever can lead to fatalities, with a death rate of 1 to 2% [61]. Pregnant women are considered to face the same risks as infected farm animals, namely increased risk abortion, stillbirth, and premature delivery [249]. High titers of subtype Phase II Q Fever antibody (IgG IFA ≥1:128) are considered supportive laboratory evidence for Acute Q Fever infection and are often used as a surrogate surveillance measure for Q Fever in Public Health surveillance [61], [249], [255], [256]. In our prevalence analysis presented in the next subsection, we use a more selective definition for laboratory-supportive Acute Q Fever infection to increase the likelihood of identifying true Acute Q Fever associated infections. Specifically, we define Acute Q Fever infection as a case with (a) Phase II IgG IFA ≥1:128, (b) Phase II antibodies ≥ 4x Phase I antibodies, and (c) Phase I IgG IFA < 1:800).^43^

Chronic Q Fever is challenging to diagnose and also to treat with existing antibiotics. It causes significant heart disease and high mortality rates. Notably, Chronic Q Fever can develop in infected individuals who have never had Acute Q Fever and so it may go unnoticed for years after the initial infection. It is usually fatal if left untreated [69], [249]. For example a 2018 follow up of a recent 2008 to 2010 outbreak showed that among the 519 chronic Q Fever cases identified, 86 patients had died [69]. A persistently high Phase I antibody level (IgG IFA ≥1:800) that persists after Acute Q Fever infection is a risk factor for Chronic Q Fever but is not diagnostic [61].

### Human Infection and Disease Prevalence

It is uncertain how many human Coxiella infections and cases of Q Fever there are as it is widely acknowledged that many, if not most, cases of Q Fever are unrecognized and do not come to medical attention [61], [249]. Existing Q Fever disease notification systems are “passive” surveillance systems in which disease prevalence is determined by the number of cases diagnosed and then reported by medical providers [252]. ^44^ “Passive” surveillance systems, while useful for trends analysis and disease outbreak identification, undercount the disease prevalence as undiagnosed cases and persons without access to health care are not included. In contrast, “active” surveillance provides less biased estimates. “Active” surveillance consists of actively surveying the general population with a scientifically based sampling strategy and collecting data and blood samples to determine each person’s infection or disease status.

While no country has national estimates of Q Fever prevalence, we provide approximate estimates for US prevalence here. The US National Health and Nutrition Examination Survey (NHANES) used active surveillance to provide national representative Coxiella burnetti serum antibody levels in the 2003 to 2004 survey [62], [257].^45^ Q Fever seroprevalence was determined by screening with an enzyme-linked immunosorbent assay and confirmed by using standard immunofluorescent antibody (IFA) testing [61]. The overall US prevalence of any positive antibody (any Phase I or Phase II IgG IFA ≥1:16) in US adults 20+ years of age was 3.1% of the population (95%CI 2.1% to 4.3%) [62].^46,47^ This suggests that 6.1 million persons (95% CI: 4.2 to 8.5 million) have some type of Q Fever infection, including acute infections, recovery phase of acute or chronic infections. We note that while a recent analysis of the NHANES data demonstrated a clear association with agricultural work, 80% of positive Coxiella NHANES test results were in persons working in non-agricultural sectors [258]. Similarly, 60% of reported Q Fever cases had no association with farming or animal processing based on a review of cases in the US Centers for Disease Control and Prevention’s (CDC) National Notifiable Disease Surveillance System (NNDSS) and state and local US Public Health Departments [259].

### Acute Q Fever

We further analyzed the NHANES 2003 to 2004 Coxiella dataset to estimate the US seroprevalence Acute Coxiella infection rates. Using the standard criterion (Phase II Q Fever antibody IgG IFA ≥1:128) [255], [256], laboratory supportive evidence of Acute Q Fever infection is 1.5% of the general US adult population (95% CI: 1.0% to 2.0%). This corresponds to 20% of all positive tested samples, an estimated 2.5 million persons. Using our stricter definition, which has a higher likelihood of reflecting Acute Coxiella infection, the US population prevalence was 0.9% of the general US adult population (95% CI: 0.4% to 1.4%), which corresponds to 15% of all positive tested samples and an estimated 1.9 million persons, see **Table H1**.

Assuming 40% of persons acutely infected with Coxiella will develop Q Fever, the US incidence of Acute Q Fever cases is estimated at 0.4% of the general US adult population (95% CI: 0.2% to 0.6%), which corresponds to 820,000 adults (95% CI: 410,000 to 1.2 million adults). Assuming a 1.5% fatality rate, this corresponds to 12,000 deaths (95% CI: 6,000 to 18,000).

As previously mentioned, 70 Q Fever cases were reported (passive surveillance) nationally in 2003 and 2004 and 153 Acute Q Fever cases were reported in 2017 [64]–[66].

**Table H1.** NHANES 2003 to 2004 Coxiella burnetti IFA Serology Prevalence Analysis

|  | N<br>(people) | Cases<br>(people) | Fraction<br>US<br>Adults<br>(%) | 95% CI<br>(%) | CPS<br>Total<br>(million<br>people) | CPS<br>95% CI<br>(million<br>people) |
| --- | --- | --- | --- | --- | --- | --- |
| *General Serology Estimates* |  |  |  |  |  |  |
| Any Positive QF Serology ≥ 1:16 | 4437 | 180 | 3.1 | 2.1 to 4.3 | 6.1 | 4.2 to 8.5 |
| All QF Phase II Serologies ≥<br>1:128 | 4437 | 91 | 1.5 | 1.0 to 2.0 | 3.1 | 2.1 to 4.1 |
| *More Selective Serology Estimates* |  |  |  |  |  |  |
| Acute Q Fever Supportive | 4437 | 50 | 0.9 | 0.4 to 1.4 | 1.9 | 0.8 to 2.9 |
| All Other QF Positive Serologies | 4437 | 130 | 2.2 | 1.3 to 3.1 | 4.5 | 2.7 to 6.4 |

Abbreviations: N = total sample; % = prevalence; 95%CI = 95th% confidence interval; CPS = US Census Current Population Survey (US adult population); QF = Q Fever; IFA = antibody Immunofluorescence Assay; Phase II = initial acute disease QF IgG antibodies.
Notes: Phase II QF antibodies ≥ 1:128 is US Centers for Disease Control & Prevention definition of laboratory supportive evidence of acute Q Fever infection; Selective Acute QF Supportive infection definition is Coxiella (Phase II antibody ≥ 1:128; Phase II IgG IFA ≥ 4x Phase I IgG IFA; and Phase I IgG IFA $<$ 1:800). Total 2003 to 2004 US adult population is 205,284,670 people.

### Comparison to Prior Work

Prior studies suggest that the Q Fever prevalence estimates derived above may be plausible. First, the overall US population Coxiella infection rate of 3.1% is similar to that reported in major European outbreak studies that used IFA testing. Second, a small-scale CDC methodology study examined the 2000 to 2011 Q Fever mortality case underreporting using Capture- Recapture Analysis. Reported Q Fever cases in the US National Death Index (n = 25) were compared to the CDC reported Q Fever case fatalities (n = 9). While strongly limited by small sample sizes, this study estimated a total of 129 fatal Q Fever cases (95% CI: 62 to 1,250 cases) occurred nationally over this time period [259]. Third, the CDC also performed a separate Coxiella seroprevalence study using data from US commercial medical laboratories from tests ordered by medical providers to evaluate symptomatic patients. While this latter survey was not nationally representative, 16% (2,039) of the 12,821 specimens tested were positive for Coxiella [255]. Finally, serological analysis of stored sera from population-based epidemiology studies and medical diagnostic laboratory testing shows Coxiella burnetti infection prevalence rates in other countries is similar to those seen in the United States (Netherlands 2.4%; Australia 5.6%) [260], [261].

1 For readability, much of the focus of this paper is on ≤5 μm AD infectious particles. However, we note that the research presented in this paper is also relevant to larger particles.

2 The infectious particle can, in theory, carry one or more infectious pathogens and be physically larger than the pathogen itself, see the *5*.*3*.*2*.*Single Particle Infectivity* section.

3 R_0_ is the basic reproduction number and is defined as the expected number of secondary cases produced by a single (typical) infection in a totally susceptible population (dimensionless). If R_0_ < 1, then an exposed person can become ill but typically does not infect others. If R_0_ >1, then further person-to-person infection spread may occur.

4 The number of particles required to change the behavior of the local atmosphere depends on the (a) atmospheric volume of interest, (b) particle size and density, (c) release duration, and (d) ambient wind speed [9]. For context, assuming a light wind (1 m s^-1^) and monodisperse particles as dense as water (1000 kg m^-3^); a 1 m^3^ release volume can contain at least 10^16^ 0.1 µm diameter particles, 10^13^ 1 µm diameter particles, or 10^10^ 10 µm diameter particles without violating this assumption.

5 Standard wind measurements are typically only accurate to within 5 degrees [10]–[14]. Thus plume predictions based on such a single measurement may be offset by as much as a 1 km by the time the plume is 10 km downwind.

6 “Less than one” exposures occur when in a mathematical model the expectation value of the individual exposure is less than one infectious agent.

7 These equations assume that the air within the building is well mixed. As the mixing process takes time and depends on the room/building/population details, exposures (and infection probability) may be higher in the room occupied by an infected individual. We note that within room spread is an area of active research, e.g., [32], but the well-mixed assumption has also been often, but not always, been employed by prior modeling studies that examined indoor bioaerosol dynamics and disease transmission, e.g., [6], [7], [33], [34], and references therein. For context, the mixing time constants for both buoyant and mechanical flow conditions in laboratory studies of room mixing are of order 10 min to < 1 h [35], [36].

8 This is an assumption used to simplify the analysis presented in this report. Methodology exists for modeling mobile populations that is compatible with RSA analysis, see the *5*.*4 Potential Future Efforts* section and reference [31].

9 1 μm AD particles are chosen here for theoretical modeling purposes. However, this particle size is characteristic of many infectious bacterial cells. Furthermore, infectious agents, including viruses, may be carried on, or in, larger particles comprised of other material such as respiratory fluids, see the 5.*3*.*2. Single Particle Infectivity* section.

10 This transport timescale calculation is intended for illustrative purposes as it does not consider the effects of atmospheric turbulence (e.g., plume dispersion) nor the known increase of wind speed with height above the earth’s surface.

11 For context, respirable size (1 to 5 μm AD) particle deposition losses are often limited (approximately 0.1 h^-1^ or less) on these scales. Deposition rates depends on particle size, atmospheric conditions, and surface characteristics. For this illustrative calculation, we have assumed (a) a 1 cm s^-1^ deposition velocity (the speed at which the particle travels to a surface and is lost) [38], (b) a 300 m boundary layer height (the height of the air near the surface that is well mixed – this value corresponds to stable, nighttime conditions), and (c) that the atmospheric boundary layer is a well- mixed system and so the deposition loss rate = deposition velocity / boundary layer height [39].

12 The equations shown in **Figure 7** are based on the mean of the individual slope and intercept for each weather case (r^2^ > 0.99 for all cases). The individual values varied less than 15% from the mean values provided.

13 This case is also applicable to initial stage of epidemic that is later propagated by secondary transmission.

14 A linear regression (not shown) of the data presented in **Figure 8** yields log_10_(Modeled Relative Disease Probability) = 1.05 (± 0.08) * log_10_(Observed Relative Disease Incidence) – 0.10 (± 0.05). A linear regression with all available data reduces the correlation, r^2^ = 0.53, but yields a similar line, log_10_(Modeled Relative Disease Probability) = 0.85 (± 0.13) * log_10_(Observed Relative Disease Incidence) - 0.02 (± 0.09). The provided parameter uncertainties are ±1σ.

15 As a practical matter, the population within the region of interest may be low and so the corresponding disease incidence rates may be unreliable.

16 We speculate, but have not proven, that this result is related to the well-known fact that away from the source, airborne plume dispersion is proportional to (distance)^1/2^, see **Supplemental Material B: Key Atmospheric Transport and Dispersion Modeling Concepts**. If true, then we would expect that the slope of log_10_(distance from source) vs. log_10_(relative infection rate) would be closer to −1 for impacts that take place close to the source, where dispersion eddies are well-correlated and the airborne plume dispersion is proportional to distance.

17 (a) This statement is true in the mathematical limit that the “base” and “reference” distance in the prior study equations are large relative to the constant infectivity distance, (b) The modeling presented in this study is limited to less than 20 km from the source.

18 This fraction is expected to vary with particle size, airborne infectivity loss rate, and building type.

19 Direct contact and vectors (ticks) are also known to transmit Coxiella infections and so these estimates may not be solely due to airborne transmission [61], [63].

20 A notable exception is focused vaccine safety studies which perform population-based assessments of background prevalence of rare diseases as a baseline to compare against rare adverse vaccine events, e.g., [68].

21 **Figure 5a** suggests that the outdoor release of 10^3^ to 10^5^ particles would infect a downwind outdoor individual. A factor of 10x reduction is added to account for both (a) the fraction of particles released indoors that exit the building and enter the outdoor atmosphere and (b) protection that downwind buildings protect their occupancy, see **Supplemental Material G: Infection Estimates**.

22 The dose required to infect 50% of the human subjects is 1.18 bacteria (95% CI: 0.76 to 40.2).

23 Bacteria have the capacity to bind to a wide variety of surfaces and particles and depending on the vehicle, multiple bacteria can become airborne, including on the surface of human skin cells, e.g., [84]. Furthermore, viruses and bacteria such as Tuberculosis, Coxiella burnetti, Legionella Pneumophilia, Chlamydia pneumoniae and others are intracellular pathogens, i.e., they infect host hosts by penetrating within cells where they reside and multiply [85]. Thus, there is the potential for multiple pathogens to be present in a single particle when the infected cells are exhaled (or exfoliated) as airborne particles [86]. We are unaware of a field experiment demonstrating multiple infectious agents present on a single airborne particle, no doubt due to the technical challenges involved [87]. However, this characteristic has been demonstrated in the laboratory, e.g., [88].

24 (1) This 0^th^ order estimate is based on volume considerations and assumes a spherical geometry. (2) the number of agents actually present in a particle will also depend on other factors and can be much smaller.

25 Also, bacteria have the ability to repair injuries induced by environmental damage. With regard to UV radiation, [99] reviews of the effects of UV on viruses and makes the point that much of the existing published literature relates to the more potent UV (UV-C) radiation spectrum used in commercial UV germicidal irradiation devices. As UV-C does not reach the earth’s surface; natural ground-level solar UV wavelengths fall in the less viricidal UV-A and B spectrum above 290 nm.

26 As an illustrative example, moderate winds, 4.5 m s^-1^, would transport an outdoor airborne particle 1 km downwind in about 4 min. This example is intended for illustrative purposes as it does not consider the effects of atmospheric turbulence (e.g., plume dispersion) nor the known increase of wind speed with height above the earth’s surface. Furthermore, this calculation focuses on outdoor emissions and exposures. Particles emitted indoors will take time to move outdoors and, similarly, outdoor particles will take additional time to travel indoors to expose indoor people.

27 We note that the effects of known variations in the building stock, population demographic distributions, and daily population mobility patterns could be assessed prior to the outbreak event and used to in real time to provide higher fidelity quantitative results [30], [31].

28 Many viruses and microorganisms are opportunistic and so may have multiple, simultaneous disease transmission pathways.

29 The airborne infection probability is predicted to be directly proportional to population density and so reducing the number of people at risk in high density regions, such as through zoning changes or preventative evacuation, could reduce the overall disease spread rate.

30 airborne microbial threats to animal livestock or resulting from agricultural activities

31 The airborne disease transmission pathway can contribute to overall disease transmission, even when other – droplet (> 5 μm AD particles) and contact – pathways are important, e.g., [6], [7].

32 The physics of outdoor-origin particles penetrating buildings and exposing indoor individuals has been studied, e.g., [30] and is demonstrated by (a) indoor measurements of outdoor-origin PM_2.5_ particles [34], [119] and (b) airborne Bacillus thuringiensis bacteria infiltration into buildings [23].

33 The number of particles required to change the behavior of the atmospheric depends on the (a) atmospheric volume of interest, (b) particle size and density, (c) release duration, and (d) ambient wind speed [9]. For context, assuming a light wind (1 m s^-1^) and monodisperse particles as dense as water (1000 kg m^-3^); a 1 m^3^ release volume can contain at least 10^16^ 0.1 µm diameter particles, 10^13^ 1 µm diameter particles, or 10^10^ 10 µm diameter particles without violating this assumption.

34 These computational marker particles are a large sample from the possible particles emitted and are used in estimating downwind concentrations along the prescribed contaminant emission amount. These particles do NOT directly represent individual physical aerosols.

35 Respiratory minute volume is a traditional unit in pulmonary medicine – this study uses respiratory second volume.

36 For context, an airborne exposure of 10^4^ particles s m^-3^ would result in a typical individual inhaling a single particle (assuming a breathing rate of 10^−4^ m^3^ s^-1^ [235]).

37 One example is a group of people in a room where the air is well mixed.

38 One example is a neighborhood where people are in different buildings that provide varying degrees of protection from an outdoor plume of airborne, infectious particles.

39 We note that the contaminants of concern here are from the outdoors which, due to the air change mechanisms, can result in a relatively uniform indoor spatial contaminant distribution. For example in modeling indoor concentrations of outdoor ozone, *Hayes* [241] demonstrated that the indoor/outdoor ozone concentration ratio was insensitive to the degree of indoor mixing in both single- and multicompartment models for residences and office buildings. For context, the mixing time constants for both buoyant and mechanical flow conditions in laboratory studies of room mixing are short, 10 min to < 1 h, compared to the exposure times of interest [35], [36].

40 These equations assume that (a) initial (t = 0) indoor air concentrations are zero and (b) material removed from the indoor air is not reintroduced at a later time (e.g., resuspension of deposited particles).

41 These computational marker particles are a large sample from the possible particles emitted and are used in estimating downwind concentrations along the prescribed contaminant emission amount. These particles do NOT directly represent individual physical aerosols.

42 The dose required to infect 50% of the human subjects is 1.18 bacteria (95% CI: 0.76 to 40.2) [15], [83].

43 Coxiella antibody “Phases” are a laboratory methods notation where Phase 1 antibodies occur first in embryonated egg cultures and Phase II antibodies later. In human Acute Q Fever, Phase II antibodies peak first at high levels (≥ 1:128) 1 to 2 weeks after the start of symptoms when Phase I antibody levels are low. Phase I antibodies peak some weeks after. Our selective acute infection criteria was chosen to reflect these dynamics and relevant human levels [61], [249].

44 In the US there is mandatory reporting of Q Fever to the US Centers for Disease Control and Prevention national disease reporting system, see https://www.cdc.gov/qfever/info/index.html.

45 The NHANES survey provides nationally representative estimates using a demographically-based complex, multistage survey design to minimize bias [62], [257].

46 Antibody prevalence in men and woman was 3.8% and 2.5%, respectively.

47 NHANES 2003 to 2004 demographics and Coxiella serology data were downloaded from the NHANES website. Data assembly and analysis used SAS™ (Release 9.4, SAS Institute, Inc., Cary, NC). Survey design variables (strata and primary sampling units) and survey sample weights were used to account for differential probabilities of participant selection in the complex multistage survey design. Sample weights provide nationally representative estimates and adjust estimates for nonresponse and noncoverage. Statistical estimates are age-adjusted to the US population. Results with relative standard errors >30%, with <12 degrees of freedom, or low sample sizes are not presented.

